# Epidemiological characteristics and vaccination impact scenario modelling of concurrent Clade I mpox outbreaks in the Democratic Republic of the Congo and Burundi

**DOI:** 10.64898/2026.02.24.26346883

**Authors:** Ruth McCabe, Edward Knock, Alba Halliday, Victoria M Cox, Daniela Olivera Mesa, Kieran Chopra, Brian Ajong, Jean-Claude Bizimana, Thierry Kalonji, Olivier Kamatari, Trystan Leng, Rosie Maddren, Hypolite Muhindo Mavoko, Placide Mbala, Guillaume Morel, Liliane Nkengurutse, Odette Nsavyimana, Joseph Nyandwi, Kanchan Parchani, Anh Pham, Thomas Rawson, Alexander Shaw, Charles Whittaker, Azra C Ghani, Neil Ferguson, David Niyukuri, Lilith K Whittles

## Abstract

In 2024, mpox cases surged in the Democratic Republic of the Congo (DRC) with cross-border spread to Burundi. We developed a transmission-dynamic model calibrated against surveillance data to understand drivers in enzootic (Clade Ia) and non-enzootic (Clade Ib) areas, and the potential impact of vaccination. In non-enzootic areas we estimated that 58-84% of transmission occurred within sexual networks. *MVA-BN* vaccination of sex workers could have averted 91% (95% CrI 81%–98%) of infections in Sud Kivu (DRC) but only 35% (95% CrI 26%–47%) in Bujumbura (Burundi), due to later outbreak detection. In historically enzootic Equateur (DRC), ongoing zoonotic spillover best explained sustained incidence. There, pledged *Lc18m8* vaccines could have averted 42% (95% CrI 40%–46%) of infections; prioritising children improved impact. Across all settings, doubling vaccine coverage by using a single dose of *MVA-BN* outperformed two-dose strategies. Timely detection and tailored vaccination strategies are critical to reducing mpox burden.

## Introduction

Mpox, the disease caused by monkeypox virus (MPXV), has rapidly escalated into a global health priority, triggering two Public Health Emergency of International Concern (PHEIC) declarations by the World Health Organisation (WHO) within the last three years (1). Historically, the two MPXV clades (Clades I and II) were enzootic in Central and West Africa respectively, where mpox circulated in rodent reservoirs. Historical transmission in humans was characterised by sporadic spillover from rodent reservoirs, followed by short chains of human-to-human spread via close personal contact (2). However, the 2022–2023 global outbreak, driven by novel sub-clade IIb, marked a turning point. Rapid international spread through sexual networks, primarily among men-who-have-sex-with-men (MSM), resulted in over 97,000 reported cases and 200 deaths worldwide (3), prompting WHO to declare a PHEIC in July 2022 (6). The PHEIC was lifted in May 2023 after case numbers declined; however, Clade IIb continued to be detected globally throughout the year, while cases of Clade I in enzootic settings rose steadily (1,4–6).

The Democratic Republic of the Congo (DRC) continues to report the highest burden of Clade I mpox (3,7). This is particularly evident in enzootic provinces such as Equateur, where Clade Ia circulates in zoonotic reservoirs, with spillover transmission primarily affecting children and their households (8,9). In 2024 however, Clade I cases rose sharply and spread beyond historically affected regions (10). In Sud Kivu, a non-enzootic province, a novel sub-clade (Ib) was detected in late 2023, estimated to have emerged in September of that year (11). Early case patterns suggested Clade Ib was spreading primarily through sexual contact, initially within commercial (hetero-)sexual networks, before extending into the broader population (12,13). In addition to differences in transmission, crude estimates of mortality suggested Clade Ib may be milder than Clade Ia has been historically in enzootic areas. However this apparent difference may be confounded by case age demographics, healthcare access, transmission routes, and underlying comorbidities (14). These factors mean that the extent of any true phenotypic differences between Clades Ia and Ib, or indeed Clade IIb, remain uncertain.

By July 2024, DRC had reported over 15,600 suspected cases (more than double the number reported in 2023) and 535 deaths (almost 20% increase compared to 2023), with 85% of deaths occurring in children (15,16). Cases of Clade Ib were subsequently detected in neighbouring countries, including Burundi, Rwanda, and Uganda (the first recorded cases of mpox in these countries), raising further concerns about unmitigated spread of mpox throughout the region (17). On 14 August 2024, the WHO re-declared mpox as a PHEIC based on these epidemiological signals (18). By the end of 2024, DRC had reported approximately 45,000 suspected cases and over 1,100 deaths (19), with WHO noting that only one third of suspected cases and very few deaths were tested due to a shortage of resources (3). Burundi reported the second highest number of cases in the region in 2024, with almost 3,000 confirmed cases but just one death (20). Mpox cases continued to be reported throughout 2025, although at a slower rate than the previous year (3). On 5 September 2025, WHO lifted the PHEIC declaration but acknowledged that it remained an emergency, with the African Centre for Disease Control also lifting its emergency declaration in January 2026 (21,22).

Two vaccines, originally developed for smallpox, are available for use against mpox: *MVA-BN* and *LC16m8* (4,23–25). *MVA-BN* is a non-replicating vaccine requiring two 0.5mL doses to achieve maximum effectiveness, but which offers partial protection after one dose (23). In supply-constrained outbreak situations, WHO recommends its off-label use for all ages in either a single 0.5mL subcutaneous dose or 0.1mL fractional dose administered intradermally (26,27). *LC16m8* is a minimally-replicating single-dose vaccine which is licenced without age-restriction and is listed by WHO under the Emergency Use Listing mechanism for use from one year of age; however, it is contraindicated for immunocompromised individuals and pregnant women, raising concerns about its use among sex workers (SWs), who may have higher rates of undiagnosed HIV (23,28,29).

Mass vaccination against mpox is unlikely to be feasible in the region, given limited vaccine availability relative to populations at risk (30–32), operational constraints on uptake and delivery (33) and insecurity areas of eastern DRC (34); accordingly, it is not recommended by WHO in outbreak settings (23). In practice, vaccination in DRC has prioritised children and populations at higher risk, including sex workers and case contacts, as well as geographical hotspots, with approximately 775,000 *LC16m8* doses and 690,000 *MVA-BN* doses (of the 3- and 2.3-million pledged to the country, respectively) administered since October 2024 (31). The mpox response in Burundi has relied primarily on clinical management and enhanced surveillance rather than vaccination (35). However, uncertainties remain as to the most effective vaccination strategies to maximise the impact of limited stocks in regions with different patterns of transmission.

We used a mathematical model of mpox transmission, disease progression and vaccination to investigate the epidemiology of Clades Ia and Ib and evaluate the potential impact of population-level vaccination targeting children and sex workers. We fit our model to data from: (1) Equateur province, an enzootic region in DRC where Clade Ia circulates; (2) Sud Kivu, a non-enzootic DRC province where Clade Ib first emerged; and (3) Bujumbura, the most-affected region in non-enzootic Burundi, which was the second most-affected country in 2024 after DRC. We assess region-specific differences in the transmissibility, severity, and reporting of cases in these three settings, then model retrospective vaccination scenarios to examine the differential impact of targeting strategies and rollout speed on the numbers of cases and deaths that could have been averted under a range of dose supply constraints.

## Methods

We developed a population-level stochastic susceptible-exposed-infected-recovered (SEIR) mathematical model of mpox transmission, stratified by age-group and vaccination status. We used this model to describe the transmission dynamics and disease progression of mpox outbreaks in both the community and commercial sexual networks of Equateur, Sud Kivu, and Bujumbura. We provide a high-level summary of the model here with full details given in the Appendix.

## Modelling transmission and disease progression

In our model, the total population in each region is stratified into 18 groups: 16 representing age groups and two representing key populations, namely: SWs and people who buy sex (PBS). In each region, susceptible individuals can be exposed to mpox infection via different routes: (i) non-sexual contact (ii) sexual contact, and in Equateur where Clade Ia is enzootic, (iii) via zoonotic spillover. After an incubation period, exposed individuals become infectious before subsequently recovering or dying, with the proportion determined by the age-stratified case fatality ratio (CFR). Deaths were not explicitly modelled in Burundi, due to only one death being observed nationally as of January 2026 (3). The model was written in R (v4.5.0) (36) using the *odin2* package (v0.3.24) (37) and is available in the package *mpoxseir* (v0.2.28) (38).

## Model fitting and parameterisation

We calibrated our model to surveillance data in each of the three regions within a Bayesian framework. A comprehensive overview of the data sources used for model calibration is presented in the Appendix (a summary is provided in Table S6). For DRC regions, we fitted our model to time series data on weekly cases and deaths (age-disaggregated where available), the proportion of cases among SWs, and the crude CFR reported in each age group. In Bujumbura, we fitted to weekly case counts and the age-distribution of cases that was observed in Burundi as a whole. We fixed model parameters such as demographics, contact patterns, and disease progression to values obtained from the literature, where available. Only parameters with substantial epidemiological relevance or high prior uncertainty were inferred from the data (full details documented in the Appendix). We estimated either six (Bujumbura) or nine (Equateur and Sud Kivu) parameters during calibration. Prior distributions for these parameters were informed by published literature, where available, with weakly informative priors restricting the parameters to plausible bounds otherwise (Table S7). To sample from the posterior distribution of the model parameters and obtain fitted model trajectories compatible with the observed data, we used particle Markov chain Monte Carlo (pMCMC) methods (39).

## Modelling vaccination

We extended the model to incorporate vaccination at the population-group level. Each population group was further stratified by vaccination status: (1) previously vaccinated against smallpox, (2) unvaccinated, (3) received one mpox vaccine dose of either vaccine, or (4) received two doses (for *MVA-BN* only). Vaccines were assumed to provide protection against infection 14 days after administration (40). We did not model waning of protection due to the short analysis timeframe.

We evaluated how cases and deaths in each region might have changed under different vaccination scenarios (see: *Vaccine type scenarios* and *Prioritisation scenarios*). Impact was assessed over a two-year period (Jan 2024 – Jan 2026) with vaccination starting five weeks after first case detection.

### Vaccine type scenarios

We simulated retrospective vaccination strategies using the two vaccines pledged to DRC: *MVA-BN* and *LC16m8*. We considered scenarios dispensing: (1) *Lc16m8* only; (2) a mixed strategy where both *Lc16m8* and single-dose *MVA-BN* are deployed in parallel; and *MVA-BN* only, delivered under either (3) a single-dose, or (4) a two-dose regimen, where the latter strategy vaccinates half as many people, but offers higher individual-level protection.

The *LC16m8* vaccine, administered via a single dose, has estimated efficacy of around 95% based on seroconversion rates (41). *MVA-BN* requires two doses for full effectiveness of 82% against infection, however a single dose provides partial protection (74%) against infection (6). For the scenarios with two doses, we modelled a 28-day interval between doses, per manufacturer guidance (42).

### Eligibility for vaccination

The population groups eligible for each vaccine reflect evolving licensing guidance – *MVA-BN* was granted emergency approval for those over one year of age, following earlier approval for those over 18, then over 12 (26,43). We assumed *LC16m8* would not be used for SWs due to contraindications in immunocompromised individuals. UNAIDS data indicate that globally, SWs face nearly nine times the risk of acquiring HIV compared with the general population, suggesting a significantly elevated likelihood of undiagnosed or uncontrolled infection within this group (29,44). In scenarios using both vaccines, *LC16m8* was given to those under 12, and *MVA-BN* to older age groups and key populations. Smallpox-vaccinated individuals were assumed ineligible for mpox vaccination.

### Prioritisation scenarios

We modelled each vaccination strategy under different prioritisation approaches: no prioritisation, and two approaches targeting specific risk groups. As mpox infection is typically more severe at younger ages (45), we considered strategies where children under 12 year-olds are vaccinated to target levels before vaccination begins in the general population. The final prioritisation approach targets SWs first instead of children (*MVA-BN* only).

In total, we modelled nine different scenarios combining vaccine types, dosing strategies, and prioritisation approaches as shown in Table 1. Scenarios were explored sequentially: initial results from the no-prioritisation analysis indicated that a single dose of *MVA-BN* was consistently more effective at population-level than the two-dose regimen, so subsequent prioritisation scenarios (children-first or SWs-first) only considered the single-dose regimen.

**Table 1:** Overview of vaccine scenario impact modelling. **A)** the range of dose numbers and rollout speeds considered per region. In Equateur and Sud Kivu (DRC), dose numbers are derived from pledged mpox doses, while in Bujumbura (Burundi) in the absence of this information, plausible dose numbers were based on COVID-19 vaccine rollout. Rollout speeds were selected to reflect a broad range of response capacities. **B)** summary of the vaccine type scenarios and prioritisation strategies modelled. Two doses of *MVA-BN* are only modelled for under no prioritisation due to WHO guidance of only using a single dose of *MVA-BN* in outbreak settings. *LC16m8* is not modelled prioritising SWs as this vaccine is contraindicated for immunocompromised individuals.

**A) Modelled dose numbers and rollout speed (by region)**
|  | Total doses | Rollout speed<br>(doses per day) |
| --- | --- | --- |
| Equateur | 2,600 - 520,000 | 250 - 20,000 |
| Sud Kivu | 10,000 - 2,000,000 | 250 - 20,000 |
| Bujumbura | 500 - 50,000 | 25 - 20,000 |

**B) Strategies considered (all regions)**
|  | No prioritisation | Prioritise <12<br>year-olds | Prioritise<br>sex workers |
| --- | --- | --- | --- |
| *LC16m8* only | ✓ | ✓ | ✗ |
| *LC16m8* and *MVA-BN* | ✓ | ✓ | ✓ |
| *MVA-BN* single dose | ✓ | ✓ | ✓ |
| *MVA-BN* two doses | ✓ | ✗ | ✗ |

### Vaccine supply and rollout capacity

Fragmented reporting limited our ability to determine the exact number of *MVA-BN* and *LC16m8* vaccines received in the region in addition to how quickly they were rolled out in DRC. In Burundi, outbreak response was not centred on pharmaceutical measures. In our main analysis, we therefore explored plausible ranges for dose numbers and rollout speed in each region under each of the nine modelled scenarios (Table 1).

In DRC, we treated the 2m *MVA-BN* vaccines pledged to the country as an upper bound on the number of doses that could be delivered in Sud Kivu (population ∼ 5.8m). We then derived a population-scaled upper bound for Equateur (population ∼1.6m) of 520,000 doses. In Burundi, in the absence of mpox vaccine pledges, we used the number of vaccines delivered during COVID-19 as a proxy (upper bound 50,000 doses). For each region, we explored progressively lower supply scenarios by reducing dose availability down to 0.5% of these upper bounds, corresponding to 10,000 doses in Sud Kivu and 2,600 doses in Equateur respectively. In scenarios in which both vaccine types were modelled, we assumed a 2:3 allocation between *MVA-BN* and *LC16m8* doses, in line with the size of the respective totals pledged to DRC.

Daily vaccination capacity was assumed to be constant throughout the rollout period. We modelled seven different rollout speeds reflecting the number of doses that could be given per day, with a lower bound of 250 and an upper bound of 20,000 for both Equateur and Sud Kivu. Based on expert in-country guidance, we reduced the rollout speeds used in DRC by a factor of 10 for Bujumbura. In all regions, for lower dose numbers and high rollout speeds, the rollout speed was adjusted to ensure it took at least one week to give out the vaccines. We adopted 5,000 doses per day as the central value for Equateur and Sud Kivu, and 500 as the equivalent value for Bujumbura.

### Vaccine uptake and target coverage

Studies of vaccine hesitancy across the region indicate that not everyone who is offered mpox vaccination would accept it (33,46). We used the reported proportion of people willing to accept vaccination to inform the maximum vaccine coverage achievable within each population group (Table S8).

### Evaluation of vaccine strategies

We compared each simulated vaccine scenario with our reconstruction of the observed epidemic to estimate the cases and deaths that could have been averted under each strategy, both in total and per dose administered. Throughout, results are presented as the mean and 95% credible intervals.

## Results

### Mpox epidemiology

Figure 1 shows modelled infections and cases over time, alongside data on reported cases in each region, and estimates of the proportion of infections that are recorded as cases (case ascertainment).

**Figure 1:**
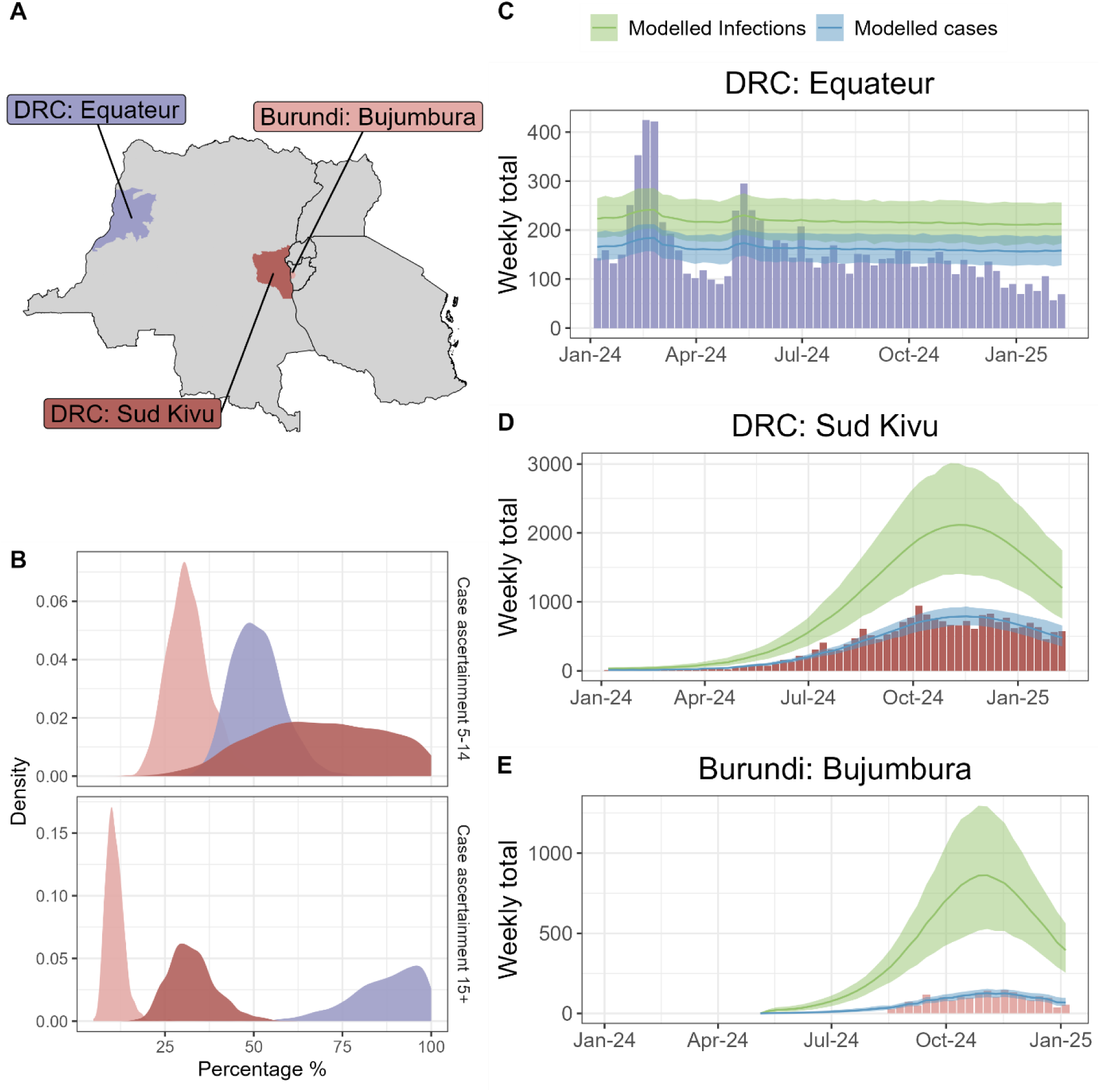
Overview of trends in cases and underreporting by region. (A) Map showing the locations of the three modelled regions of Equateur (DRC), Sud Kivu (DRC) and Bujumbura (Burundi). (B) Posterior distributions of estimated case ascertainment among 5 – 14-year-olds (top) and 15+ year-olds (bottom) in each region (colours as in A). (C) – (E) Reported cases (bars, coloured by regions as in (A) and (B)), modelled cases (blue) and modelled infections (green) for each of the three regions: (C) Equateur; (D) Sud Kivu; (E) Bujumbura. Lines represent posterior means, and shaded areas represent 95% credible intervals.

In Equateur, modelled infections and cases remained relatively constant over time (Figure 1C): the model explained patterns in the province-level case data by short-chain transmission driven by ongoing zoonotic spillover, rather than distinct epidemic waves, although the model does not fully capture fluctuations in the case time series. We estimated a relatively high level of case ascertainment overall (75%, 95% CrI 66% – 83%). In almost all posterior parameter sets (99.7%) the model inferred a higher proportion of infections were detected as cases among those aged over 15 years (87%, 95% CrI 64% – 99%) compared with 5 – 14-year-olds (51%, 95% CrI 39% – 66%) (Figure 1B, Table S11). The inferred age pattern of case ascertainment is driven by how the observed age distribution of cases and deaths compare with model predictions, which are determined by assumptions around age-specific contact patterns, and age differences in zoonotic spillover and case fatality ratios derived from previous Clade I outbreaks. No additional age variation in prior exposure to mpox was assumed beyond age-stratified historic smallpox vaccination coverage (47).

The epidemics in Sud Kivu and Bujumbura peaked in November 2024 (Figure 1D – E). The model reproduces the overall epidemic trajectory effectively, closely matching observed trends. The model reproduces the observed peak and subsequent decline in cases by allowing for a larger, under-reported, epidemic. As recovery confers protection against reinfection, depletion of the susceptible population reduces the number of people at risk, leading to a decline in transmission. We estimate that at the epidemic peak, 33% (95% CrI 8% – 95%) of SWs and 18% (95% CrI 8% – 28%) of PBS had been infected in Sud Kivu, along with corresponding figures of 27% (95% CrI 21% – 33%) and 22% (95% CrI 13% – 32%) in Bujumbura. This scenario is consistent with overall case ascertainment of 38% (95% CrI 27% – 53%) in Sud Kivu and just 14% (95% CrI 9% – 20%) in Bujumbura (Table S11). In contrast to Equateur, in the regions where Clade Ib predominates we estimate that case ascertainment was lower among 15+ year-olds than those aged 5 – 14-years-old: 33% (95% CrI 22% - 48%) vs. 69% (95% CrI 35% - 98%) in Sud Kivu and 11% (95% CrI 7% - 16%) vs. 32% (95% CrI 21% - 45%) in Bujumbura, potentially reflecting reduced care-seeking among key populations due to stigma or other structural barriers (Figure 1B, Table S11). Additionally, we determined that the epidemic in Bujumbura was detected at a relatively later stage than in Sud Kivu: by the time the first cases were reported in each location, we estimate ∼1 in 850 people in Bujumbura had been infected with mpox versus ∼1 in 35,650 in Sud Kivu.

### Transmissibility

Our estimates of the overall basic reproduction number, *R*_0_, varied by region, reflecting underlying differences in the dominant transmission pathways (Figure 2A). Transmission was below the epidemic threshold of *R*_0_ = 1 in Equateur (*R*_0_ = 0.51, 95% CrI 0.36 – 0.76), however *R*_0_ exceeded 1 in both Sud Kivu (1.58, 95% CrI 1.47 – 1.69) and Bujumbura (2.01, 95% CrI 1.77 – 2.24).

**Figure 2:**
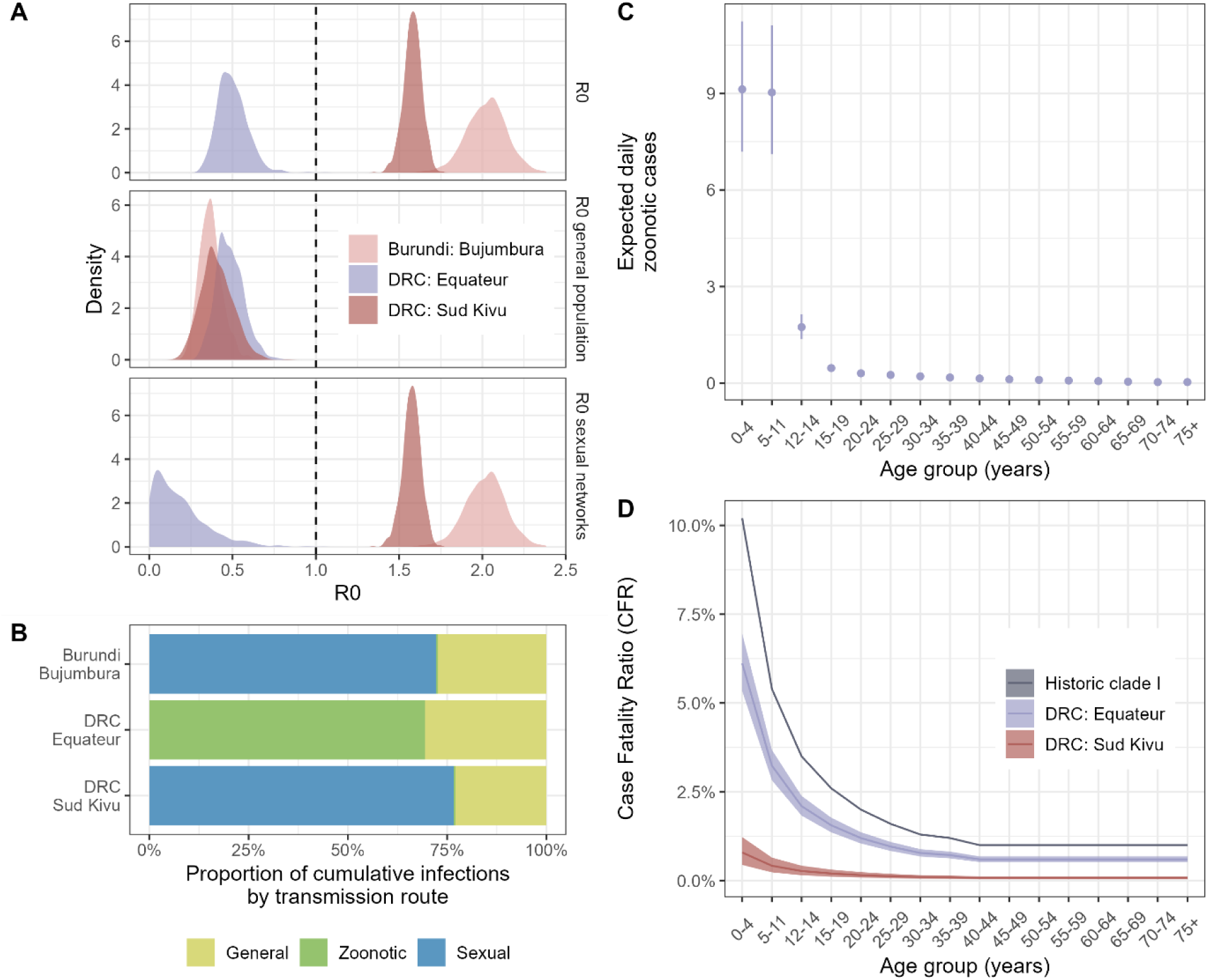
Transmissibility and severity by each modelled region (Equateur (DRC), Sud Kivu (DRC) and Bujumbura (Burundi)). (A) Posterior estimates of *R*_0_ overall (top), within the general population (middle) and in sexual networks (bottom). (B) Posterior mean of the relative contribution to transmission of the three modelled routes: general population, zoonotic and sexual. Uncertainty on means are provided in Table S11. (C) Mean daily zoonotic spillovers and 95% credible intervals by age group in Equateur. (D) Modelled case fatality ratio (CFR) for Equateur and Sud Kivu in respect to historical estimates of Clade Ia from (45). Lines represent posterior means, and shaded areas represent 95% credible intervals.

In Equateur, where Clade Ia predominates, observed case patterns are consistent with infections being primarily driven by zoonotic spillover – estimated at 69% (95% CrI 56% – 80%) (Figure 2B; Table S11). We estimated 22 (95% CrI 17 – 27) cases of zoonotic infection per day in the province, with the majority of these spillover events occurring in the two youngest age groups (9 (95% CrI 7 – 11) for under 5 year-olds and 5–11-year-olds) (Figure 2C; Table S13).

In contrast, in the two regions with Clade Ib outbreaks, the observed epidemic curves are consistent with sustained transmission being driven by sexual networks. We estimate that this route of transmission accounted for 72% (95% CrI 58% – 84%) of total transmission in Sud Kivu and 76% (95% CrI 67% – 84%) in Bujumbura (Figure 2B), based on overall SW population sizes of 20,700 (95% CrI 7,500 – 43,300) and of 4,700 (95% CrI 1,700 – 9,100), respectively. This is consistent with the average number of onward infections in sexual networks with 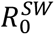 being 1.57 (95% CrI 1.46 – 1.68) in Sud Kivu and 2.00 (95% CrI 1.76 – 2.23) in Bujumbura (Figure 2A; Table S11).

The inferred contribution of general population transmission to the overall transmission rate was broadly similar across regions, with 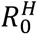 below 1 in each setting - Equateur: 0.48 (0.34 – 0.66); Sud Kivu: 0.41 (0.25 – 0.63); and Bujumbura: 0.39 (0.27 – 0.54) (Figure 2A; Table S11). We estimate that transmission within the general population comprised 31% (95% CrI 20% – 44%) of all transmission in Equateur, 28% (95% CrI 16% – 41%) in Sud Kivu, and 24% (95% CrI 16% – 33%) in Bujumbura (Figure 2B). This suggests that while non-sexual transmission has contributed substantially to cases numbers in all three regions, it is not sufficient on its own to sustain an epidemic.

### Severity

In both regions that reported mpox-related deaths, the crude reported CFR was lower than historical estimates for Clade I mpox: 0.2% in Sud Kivu; 4.5% in Equateur, compared with 8.7% (95% CI 7.0% – 10.8%) historically (2). While part of this difference may reflect the older age profile of cases in the current epidemic, particularly in Sud Kivu where transmission was primarily sexual, we find that age alone does not fully explain the observed differences in severity. Both regions experienced lower mortality rates than would be expected based on the age-specific CFR estimated from historic clade I outbreaks (45); modelled estimates were 40% (95% CrI 32% – 48%) lower overall in Equateur, and 93% (95% CrI 89% – 96%) lower in Sud Kivu. Accounting for the age-pattern of cases, the CFR was consistently higher in the Equateur Clade Ia outbreak – ranging from 6% (95% CrI 5% – 7%) in under 5-year-olds to 0.6% (95% CrI 0.5% – 0.7%) in adults – compared with the Sud Kivu Clade Ib outbreak, where the CFR ranged from 0.8% (95% CrI 0.4% – 1.2%) in under 5-year-olds to 0.1% (95% CrI 0.% – 0.1%) in adults (Figure 2D; Table S14).

#### Impact of vaccination

Figures 3 – 5 present the estimated number of infections that could have been averted through vaccination for each region considered. Analogous plots for deaths in Equateur and Sud Kivu can be found in the Appendix (Figures S20 and S21, respectively).

**Figure 3:**
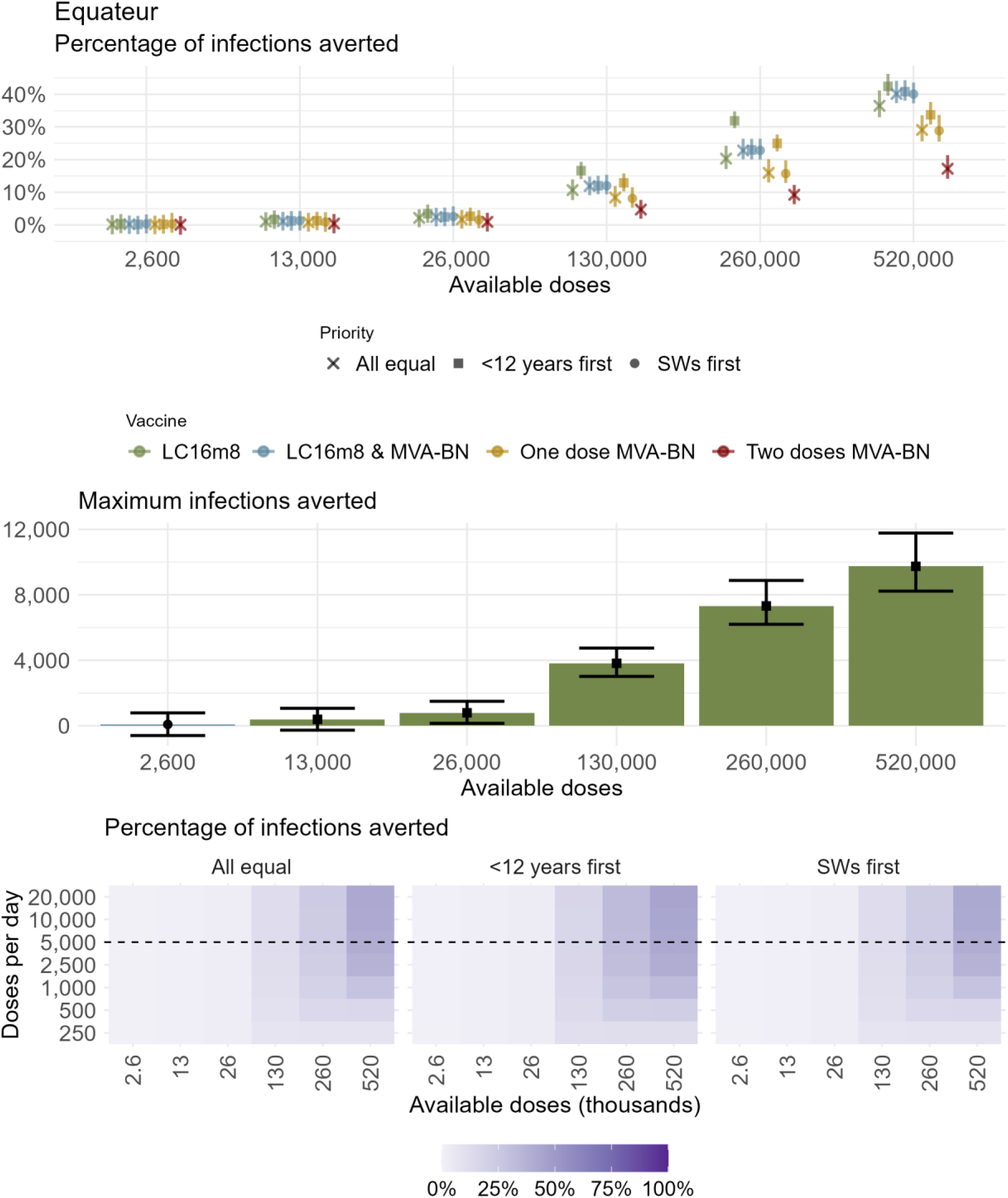
Vaccination scenarios in Equateur. Infections that could potentially have been averted over the period January 2024 – January 2026 compared with the baseline scenario under the nine modelled vaccination strategies (Table 1) and 6 different dosing levels. (A) Percentage of infections averted for the central rollout speed of 5,000 doses per day; (B) Maximum number of infections averted given the best performing vaccine and prioritisation scenario under each dosing level and for the central rollout speed; Colours indicate the four vaccine types while shapes indicate the three prioritisation strategies. Points and bars are posterior means and 95% credible intervals, respectively. (C) Posterior mean percentage of infections averted for each rollout speed, total available doses and prioritisation strategy. For the corresponding dosing level and rollout speed each grid point represents the maximum number of infections averted given the best performing vaccine and prioritisation scenario. Horizontal dashed line indicates the central rollout speed presented in A-B.

**Figure 4:**
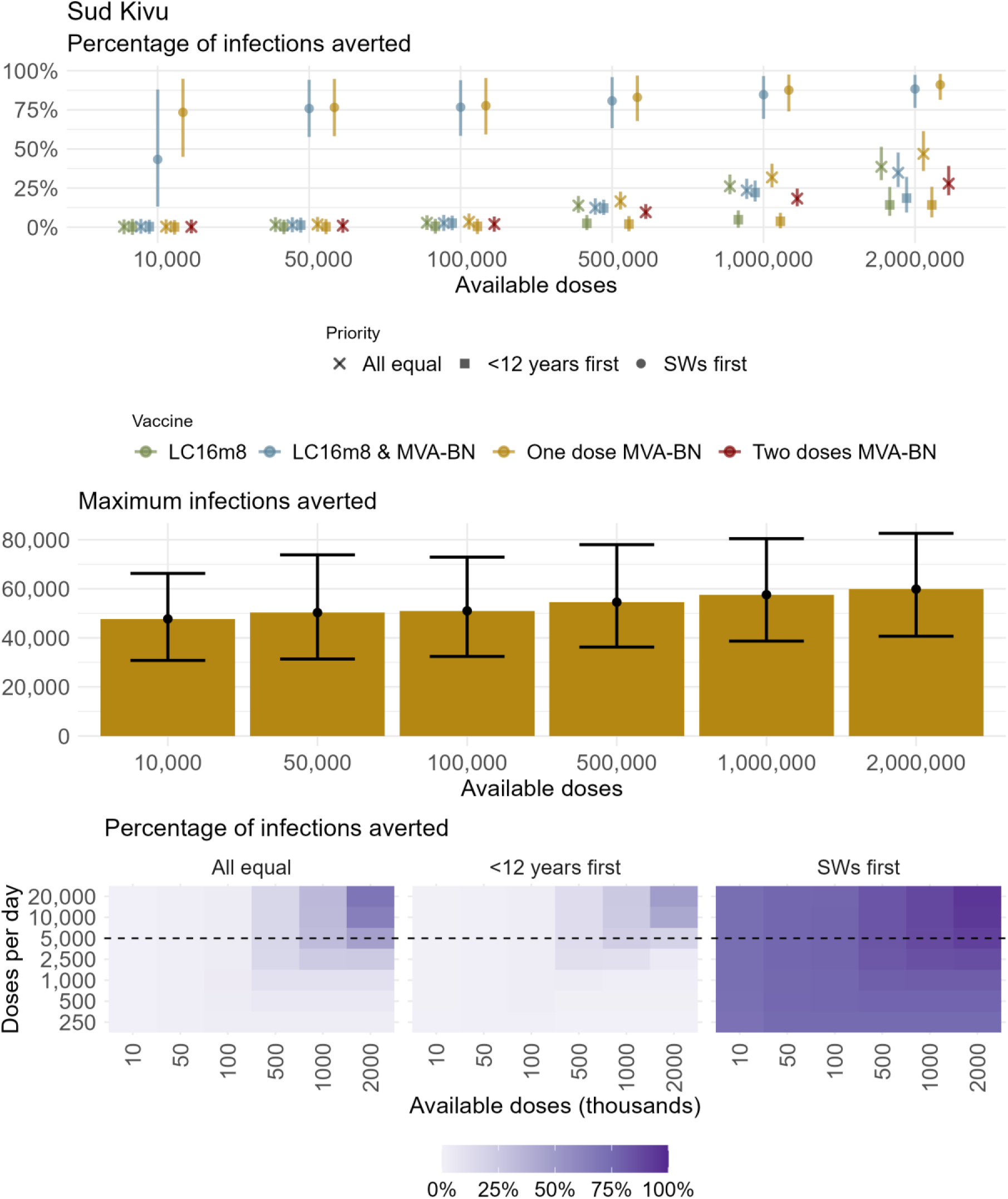
Vaccination scenarios in Sud Kivu. Infections that could potentially have been averted over the period January 2024 – January 2026 compared with the baseline scenario under the nine modelled vaccination strategies (Table 1) and 6 different dosing levels. (A) Percentage of infections averted for the central rollout speed of 5,000 doses per day; (B) Maximum number of infections averted given the best performing vaccine and prioritisation scenario under each dosing level and for the central rollout speed; Colours indicate the four vaccine types while shapes indicate the three prioritisation strategies. Points and bars are posterior means and 95% credible intervals, respectively. (C) Posterior mean percentage of infections averted for each rollout speed, total available doses and prioritisation strategy. For the corresponding dosing level and rollout speed each grid point represents the maximum number of infections averted given the best performing vaccine and prioritisation scenario. Horizontal dashed line indicates the central rollout speed presented in A-B.

**Figure 5:**
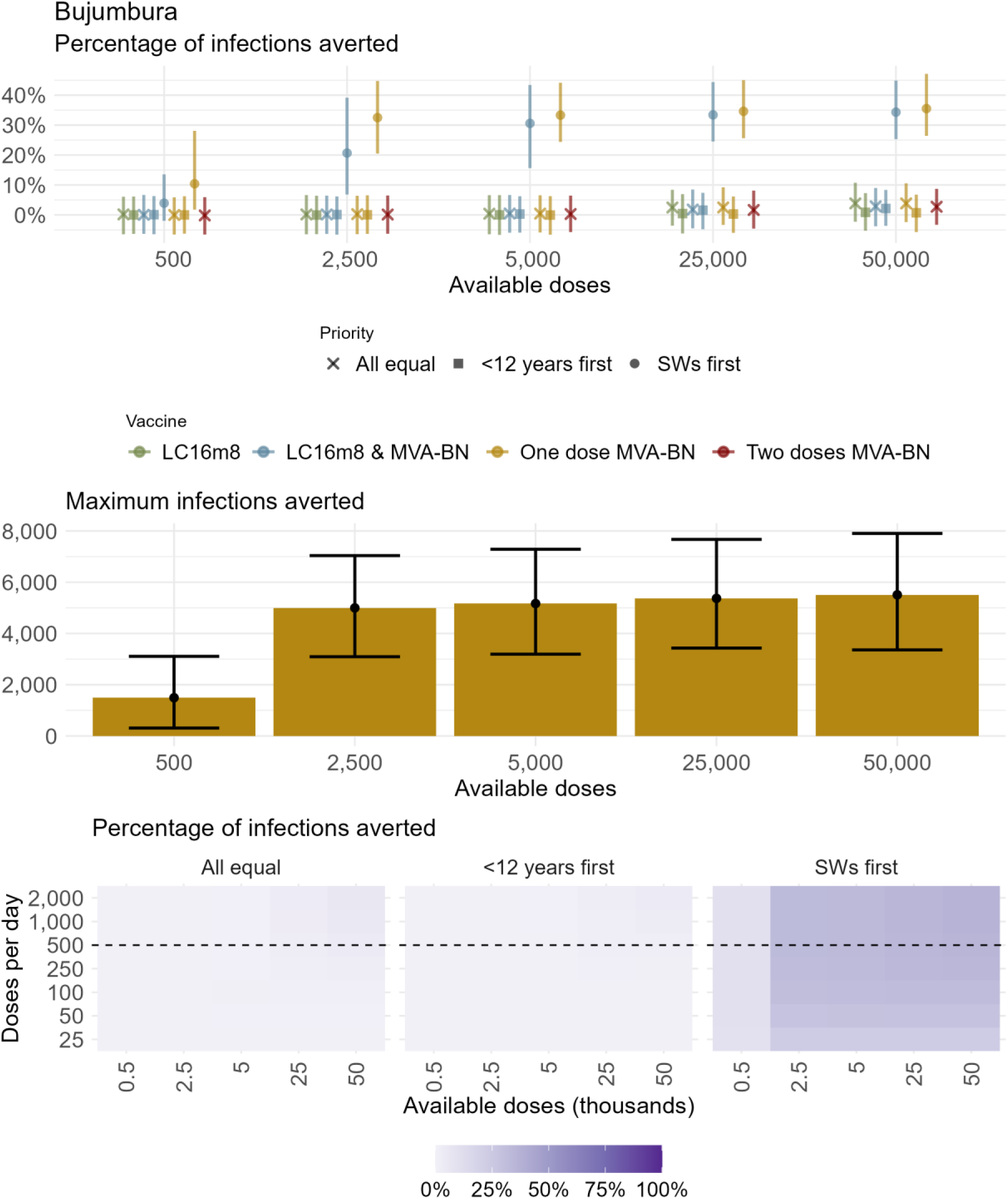
Vaccination scenarios in Bujumbura. Infections that could potentially have been averted over the period January 2024 – January 2026 compared with the baseline scenario under the nine modelled vaccination strategies (Table 1) and 5 different dosing levels. (A) Percentage of infections averted for the central rollout speed of 500 doses per day; (B) Maximum number of infections averted given the best performing vaccine and prioritisation scenario under each dosing level and for the central rollout speed; Colours indicate the four vaccine types while shapes indicate the three prioritisation strategies. Points and bars are posterior means and 95% credible intervals, respectively. (C) Posterior mean percentage of infections averted for each rollout speed, total available doses and prioritisation strategy. For the corresponding dosing level and rollout speed each grid point represents the maximum number of infections averted given the best performing vaccine and prioritisation scenario. Horizontal dashed line indicates the central rollout speed presented in A-B.

### One vs. two doses of MVA-BN

Across all settings, administering one dose of *MVA-BN* to a larger number of people was a more effective use of limited vaccine supply than giving fewer people two doses, despite the higher individual-level protection associated with two-doses. This finding was consistent across rollout speed. In Equateur, the difference was substantial. Using 520,000 doses, delivered at 5,000 doses per day, the two-dose strategy averted approximately 17% (95% CrI 14% – 21%) of cases, compared with 29% (95% CrI 26% – 34%) under a one-dose strategy (Figure 3). Similarly, in Sud Kivu with 2 million doses, an estimated 28% (95% CrI 20% – 39%) of infections could have been averted under a two-dose strategy, compared with 47% (95% CrI 36% – 61%) under a one-dose regime (Figure 4). In Bujumbura, the effect could not be clearly distinguished in our main analysis in Bujumbura because of the relatively low numbers of available doses modelled (Figure 5). however supplementary analyses using higher dose numbers showed the same qualitative result (Figure S27).

### Equateur

For each of the nine vaccination strategies modelled, increasing the number of doses given corresponded to a roughly linear increase in the percentage of infections averted. Regardless of doses available, the greatest impact was achieved by prioritising children with *LC16m8* (Figure 3A-B). Dispensing 520,000 doses in this scenario (the maximum modelled) reduced average weekly infections from around 200 to around 100 (Figure S24), resulting in up to 42% (95% CrI 40% – 46%) of infections averted and 41% (95% CrI 34% – 48%) of deaths over the study period (Figure 3A-B; Figure S20A-B), equivalent to 1.9 (95% CrI 1.6 – 2.3) infections averted per 100 doses (Figure S22A). Increasing doses available did not improve efficiency, with infections and deaths averted per dose roughly constant across vaccination strategies (Figure S22A, S23A). This pattern results from the modelled inference that transmission in Equateur is being maintained by ongoing zoonotic spillover, such that each additional vaccine dose produces a similar marginal impact.

At higher dose levels, differences between vaccine products were more important than targeting strategy in determining overall impact. Scenarios including the higher-efficacy *LC16m8* averted more cases than those using *MVA-BN* only, regardless of prioritisation, (Figure 3A; Figure S24). Prioritisation strategy was of greater importance at intermediate dose levels. For example, under 260,000 *LC16m8* doses, there were approximately 16% fewer infections and deaths when children were prioritised ahead of the general population compared to vaccinating without prioritisation (Figure 3A; Figure S20). Across all scenarios, faster rollout was associated with increased vaccine impact (Figure 3C); however, gains plateaued at rollout speeds above 2,500 doses per day, beyond which gains in impact were driven only by improvements in coverage (i.e. total dose numbers).

### Sud Kivu

In Sud Kivu, the potential impact of vaccination on the epidemic was primarily determined by whether SWs could be prioritised for vaccination. With two million doses of *MVA-BN* delivered as a single-dose strategy and prioritising SWs, we estimate that the epidemic could have been almost entirely prevented, with 91% (95% CrI 81% – 98%) of infections and 91% (95% CrI 79% – 99%) of deaths averted (Figure 4A-B). Substantial impact was estimated even with very low total dose numbers: with only 10,000 doses, giving a single dose of *MVA-BN* targeting SWs was estimated to avert 73% (95% CrI 45% – 95%) of infections, equivalent to 510 (95% CrI 320 – 710) infections per 100 doses (Figure S22B), and 74% (95% CrI 43% – 96%) of deaths (Figure 4A; Figure S21).

In contrast, strategies that did not prioritise SWs averted at most 47% (95% CrI 36% – 61%) of infections, equivalent to around 1.5 per 100 doses (95% CrI 1.0 – 2.3) (Figure S22B) – a level of population impact achievable using 200x fewer doses under the targeted strategy. Without prioritisation, 10,000 doses would have minimal impact - averting less than 1% of infections. Strategies prioritising children were the least effective, for example, with one million *MVA-BN* doses, only 4% (95% CrI 0% – 9%) of infections were averted, with impact only beginning to accrue once sufficient doses were available to extend vaccination to adults (i.e. more than one million).

Strategies involving *LC16m8* were less effective than single-dose *MVA-BN* in Sud Kivu because SWs, who are most affected by transmission in this setting, are ineligible for this vaccine, meaning higher overall population coverage is required achieve similar impact. Even with two million doses of *LC16m8,* only 39% (95% CrI 30% – 51%) of infections could be averted (Figure 4A).

Faster rollout improved epidemic impact across all vaccination scenarios (Figure 4C). When SWs could be prioritised, increasing rollout speed produced smaller additional gains because impact was already high; with 2 million doses, infections averted increased from 84% (95% CrI 72%–97%) at 250 doses per day to 94% (95% CrI 88%–99%) at 20,000 doses per day. In contrast, gains faster rollout were substantially larger in non-targeted strategies. With the same total dose supply and no prioritisation, increasing rollout speed increased infections averted from 2.5% (95% CrI 0%–7.6%) to 69% (95% CrI 58%–86%). Together, these results indicate that while direct targeting of SWs provides the greatest overall benefit, increasing rollout speed without prioritisation can substantially improve impact when targeting is not feasible or cannot be implemented rapidly.

### Bujumbura

As in Sud Kivu, strategies targeting SWs produced the greatest reductions in transmission in Bujumbura. However, achievable impact was lower overall compared to Sud Kivu because the modelled dose range was smaller (Figure 5A-B). In Bujumbura, we modelled a substantially smaller total dose supply (upper bound 50,000), reflecting the absence of mpox vaccine pledges in Burundi and using COVID-19 vaccine delivery volumes as a feasible proxy. Within this constrained supply range, prioritising SWs ahead of the general population was the only strategy considered that produced substantial impact. We estimate that dispensing 50,000 doses of single-dose *MVA-BN* could have averted 36% (95% CrI 26%–47%) of infections (Figure 5B). Similar reductions in transmission were achieved across the modelled range of rollout speeds (Figure 5C), with faster rollout providing diminishing gains in cases averted once a delivery capacity of ∼250 doses per day was reached.

Sensitivity analyses exploring higher-supply scenarios showed that once coverage exceeded 100,000 doses (∼10% of the population), further increases in vaccine supply produced only modest gains in infections averted (Figure S27). Even in these higher supply scenarios, overall impact remained lower than in Sud Kivu (generally <40% infections averted), primarily due to delayed outbreak detection, meaning that vaccination could only have commenced at a relatively later epidemic stage (Figures S25, S26).

## Discussion

We used mathematical modelling to characterise the transmission and severity dynamics of Clade Ia and Ib of mpox in concurrent Central African outbreaks, and evaluate the potential impact of different vaccination strategies, finding distinct regional patterns.

In Equateur (where Clade Ia predominates), observed case patterns were consistent with ongoing zoonotic spillover and limited secondary spread, in accordance with a previously published genomic analysis of Clade Ia (8). The level of spillover inferred by our model is informed by the age-specific contact matrix against the age structure of zoonotic spillover rates derived from historical data, along with differences in case ascertainment by age group. While these components are not independently identifiable (Figure S12), we present the full posterior range of parameters to reflect uncertainty in the relative contributions of zoonotic and person-to-person transmission. Our findings on transmissibility in enzootic regions diverge somewhat from previous estimates. In Equateur, we estimate a lower reproduction number (*R_t_* ∼ 0.5) and higher zoonotic spillover rate (∼22 cases per day) than previously estimated in Tshuapa, a neighbouring enzootic province of DRC (48). Further studies of transmission routes in enzootic settings are urgently needed to refine our understanding of underlying dynamics.

In Sud Kivu and Bujumbura, large Clade Ib outbreaks could be explained by sexual transmission, with subcritical spread (R_0_<1) outside of sexual networks. Our estimates of the proportion of sexual transmission among Clade Ib cases (72% and 76% in Sud Kivu and Bujumbura, respectively) is broadly consistent with an observational study of the first 100 mpox cases reported in Sud Kivu, 87% of whom (95% CI 79% – 93%) had symptoms initially appearing in the genital area (49). Compared with previous studies modelling the Sud Kivu outbreak, our model explains case patterns by higher transmissibility combined with age differences in case detection: *R_t_* ∼1.58 compared with *R_t_* = 1.2 − 1.3 estimated by Marziano *et al*. (50).

Our model explicitly accounts for under-ascertainment of cases, estimating the lowest reporting rates in Bujumbura (14%) and Sud Kivu (38%), and substantially higher detection in Equateur (75%). These levels of under-detection imply that observed case counts likely capture only a fraction of true transmission, limiting the ability to accurately estimate attack rates, population immunity, and epidemic stage using surveillance data alone. Mpox surveillance in our study settings is hampered by multiple challenges, including fragmented health systems, limited laboratory capacity, difficult-to-access populations (both geographically and socially), and symptom overlap with co-circulating pathogens such as measles.

Our hypothesis is that the pattern of case decline observed in the Clade Ib outbreaks is better explained by a larger outbreak than detected in surveillance data. Notably, a larger outbreak is consistent with previous estimates of transmissibility in Bujumbura as estimated by Jin *et al* (49). Nonetheless, these findings reinforce the urgent need for serological studies to assess the true attack rate and levels of infection-induced immunity in affected populations, and thereby to capture the full scope of transmission, particularly in newly affected areas.

Our estimates of severity also varied by setting, with the CFR consistently estimated to be lower in Sud Kivu (Clade Ib) than in Equateur (Clade Ia), consistent with observed crude CFR estimates (3). It has been previously hypothesised that regional differences in severity are due to different age profiles of cases, for example because Clade Ia cases are predominantly reported in children who are at higher risk of death from mpox (45). However, using our age-stratified model, we conclude that differences in overall severity are not fully explained by age patterns of cases. It remains unclear to what extent the lower CFRs in Sud Kivu reflect intrinsic differences between clades, differences in viral load owing to transmission route (e.g. consumption of bushmeat vs. sexual transmission), or unmeasured population-level factors such as prior immunity, comorbidities, or access to care.

A lack of publicly available data on the number of vaccines administered over time, where, and to whom, combined with the fact that only a fraction of the 5.3 million vaccines initially pledged had arrived in DRC by the start of 2025 (32), drove our decision to perform a retrospective, scenario-based analysis (51). We found that vaccination had the potential to substantially reduce case numbers and deaths. Across settings and deployment scenarios, we found despite a slightly lower single-dose effectiveness, giving one dose of *MVA-BN* to a larger number of individuals was consistently a more effective use of limited vaccine supplies than giving fewer people two doses. The regional epidemiological differences we inferred translated into differences in the most effective vaccination strategies in each setting. In Equateur, prioritising those aged 12 years and under for one dose of *LC16m8* ahead of the general population was consistently the most effective and efficient strategy to reduce infections and deaths, with this prioritisation being less important as dose supply increased. Our results are in line with previous modelling work by Savinkina *et al.* (52), who found that vaccinating 50% of under-15s with an 85%-efficacy vaccine in a setting with 70% zoonotic transmission averted around 40% of cases. In our most comparable scenario – vaccinating 50% of under-12s with a 74%-efficacy vaccine – we estimated a 28% reduction in cases, which scales to approximately 40% when adjusting for vaccine effectiveness and population size, suggesting strong consistency between studies despite differences in model structure.

The dominant determinant of impact in Clade Ib settings was whether vaccination could be targeted to SWs, with total dose supply and rollout speed playing secondary roles. In Sud Kivu, prioritising SWs was the most effective strategy, and we estimate that the epidemic could have been prevented in full by a targeted approach with as few as 5,000 doses. Disentangling the impact of vaccination in Bujumbura was more difficult due to both the lower realistic range of dose numbers and slower rollout speed.

Our conclusion that Clade Ib epidemics could be most efficiently and effectively brought under control by vaccinating SWs can only be realised if this key population can be identified. In the absence of a targeted strategy, orders of magnitude more vaccines were required to control the epidemics. Negative stigma attached to sex work in many settings makes prioritising key populations difficult in practice: across April – August 2024, around the time of the discovery of Clade Ib and PHEIC declaration, there was a sudden drop in women in Sud Kivu self-reporting as SWs, coinciding with a steep rise in those self-reporting as “unemployed” (53). Moreover, before the declaration of the PHEIC, there was found to be little awareness of mpox among SW within DRC (54). Where targeted vaccination to SWs is operationally difficult, we found that increasing rollout speed can partially compensate by reducing the delay in reaching high-transmission networks.

Our analysis has several limitations. Firstly, we modelled each outbreak at the provincial level; this approach treats each region as internally homogeneous, and does not capture spatial heterogeneity in transmission, access to care, or vaccine delivery. Localised clusters of transmission, particularly in urban centres or mining communities, have contributed disproportionately to case burden or vaccine demand but are not explicitly captured here. Secondly, in the absence of data on timing and intensity, we did not incorporate the potential effects of public health and social measures, such as case isolation, community engagement, or behavioural change, on transmission dynamics, thus assuming contact patterns within and between population strata (e.g. age groups, key populations) remained constant over time. Our model attributes all reductions in cases and deaths solely to the effects of population immunity, either via infection or vaccination. Thirdly, although we incorporated transmission within sexual networks through dedicated key population compartments, we faced identifiability challenges in jointly estimating the size of the SW population and transmission within the population. Similar epidemic trajectories could be reproduced by assuming a smaller population with higher transmissibility or a larger population with lower transmissibility. This limits the precision with which we can quantify the impact of targeting these groups for vaccination. However, the qualitative conclusion that early vaccination of high-transmission groups is critical to epidemic control remained robust across parameter combinations. Finally, while our model captures key outcomes such as infections and deaths, it does not account for the broader health system impacts of mpox outbreaks or the long-term consequences of severe disease.

Overall, our results highlight the potential for vaccination to control mpox, particularly in settings where transmission is concentrated within sexual networks. Our analysis evaluated high-coverage vaccination scenarios, including population-wide vaccination, to provide conceptual benchmarks for potential impact. In many of the settings considered here, mass vaccination is unlikely to be feasible or recommended due to logistical, supply, and programmatic constraints. In this context, we treat mass vaccination as a reference scenario, providing a baseline against which the efficiency and impact of targeted vaccination approaches can be evaluated under realistic supply limitations. Although our analysis has focussed on recent trends (2024 – 2025) in three regions, the findings have broader relevance to other areas affected by enzootic Clade Ia or emerging Clade Ib epidemics. Crucially, our comparison of Sud Kivu and Bujumbura illustrates the cost of delay: the greater estimated impact of vaccination in Sud Kivu reflects earlier detection and intervention, whereas delayed action in Bujumbura limited the potential to curb transmission. As the acute phase of the recent mpox emergency subsides, there is a critical opportunity to act on these lessons, investing in timely detection, improved awareness, targeted vaccination, and long-term strategies to prevent similarly serious situations emerging in the future.

## Acknowledgements

RMc, EK, AH, DOM, VMC, KC, CW, AG, NMF, and LW were funded by the Medical Research Council (MRC) Centre for Global Infectious Disease Analysis (MR/X020258/1), funded by the UK MRC and carried out in the frame of the Global Health EDCTP3 Joint Undertaking supported by the EU. TL acknowledges funding from the Wellcome Trust (grant number 312174/Z/24/Z). AP acknowledges funding from the Wellcome Trust (grant number 220900/Z/20/Z). NMF is funded by the National Institute for Health and Care Research (NIHR) Health Protection Research Unit in Health Analytics and Modelling, a partnership between the UK Health Security Agency, LSHTM and Imperial College, the MRC Centre for Global Infectious Disease Analysis (reference MR/X020258/1), funded by the UK Medical Research Council (MRC). This UK-funded award is carried out in the framework of the Global Health EDCTP3 Joint Undertaking. NMF also acknowledges funding from Community Jameel via the Jameel Institute for Disease and Emergency Analytics, and from Wellcome Trust via the Vaccine Impact Modelling Consortium (226727/Z/22/Z). LW acknowledges funding from the Wellcome Trust (grant number 218669/Z/19/Z).

The funders had no involvement in the study design, analysis, or writing of the paper.

The authors are grateful to all staff involved in one or more levels of mpox surveillance in DRC and Burundi, especially those offering surveillance at the primary health care level, involved in notifying and sampling of suspected cases. The authors are also grateful to colleagues at the World Health Organization for their helpful discussions on the analysis (Alba Vilajeliu; Judith van Holten; Olivier le Polain; Ana Hoxha; Martina McMenamin; Finlay Campbell; Lennox Kesington Ebbarnezh).

## Data availability

The model is available from https://github.com/mrc-ide/mpoxseir and the analysis from https://github.com/mrc-ide/mpox-clade1-vax-analysis.

## Author contributions

RMc, EK, and LW designed and coded the mathematical model. EK performed the model calibration and led on the technical implementation of the model. RMc, AH, VMC, and DOM performed vaccine scenario analyses. AH and VMC performed data visualisation. KC performed model validation. BA, JCB, TK, OK, LKE, PM, DN provided expert guidance on mpox epidemiology and recent outbreak trends. TL, RM, GM, KP, AP, and TR performed data extraction and formatting from the publicly available sources. AG, NF, and DN provided expert guidance on mathematical modelling. LW conceptualised and supervised the project. RMc wrote the first draft. All authors reviewed and approved the final manuscript.

