## Appendix for "Epidemiological characteristics and vaccination impact scenario modelling of concurrent Clade I mpox outbreaks in the Democratic Republic of the Congo and Burundi"

**Bibliography** 45

### S1 Methods

Below we provide a full description of the model structure, parameterisation, and fitting and parameter estimation process, and provide an overview of the vaccination scenarios considered.

#### S1.1 Model structure

We developed an age- and vaccine-stratified stochastic compartmental model of mpox transmission and disease progression. Figure S1 presents a schematic illustration of the model.
The population $N$ is stratified into 18 groups $i$ (16 age groups in 5-year bands and 2 key populations). The initial age groups $5-9$ and $10-14$ were redistributed into $5-11$ and $12-14$, respectively, to allow for 12 to be an age cutoff in some vaccine scenarios. The two key populations capturing sexual transmission are then drawn from relevant age groups to represent sex workers (SW; aged 12–49 years old) ($i = 17$) and people who buy sex (PBS; aged $20-49$ years old) ($i = 18$). There is no ageing in the model nor transitions in or out of key population strata.
Let $X_{ij}(t)$ denote the number of individuals from group $i$ and vaccine strata $j$ in disease state $X \in \{S, E_a, E_b, I_R, I_D, R, D\}$ at time $t$. We split the exposed compartment into two sub-compartments $E_a(t)$ and $E_b(t)$ to allow the incubation period to take an Erlang distributional form. The infected compartment is also split into those who will recover, $I_R(t)$, and those who will die, $I_D(t)$, to allow for different durations in the time from symptoms to recovery and from symptoms to death. Throughout:

$$E(t) = E_a(t) + E_b(t);$$
$$I(t) = I_R(t) + I_D(t);$$
$$X(t) = \sum_i \sum_j X_{ij}(t)$$

At each time step $t$, two transition steps occur. First, vaccination occurs, from which movement occurs across vaccine strata (for example, from $j = 2$ to $j = 3$) for eligible compartments; and second, disease progression occurs, from which movement occurs through $S, E, I, R, D$ compartments, within strata $i$ and $j$.

##### S1.1.1 State transitions: Vaccination

We include a class for previous smallpox vaccination ($j = 1$) as this has been shown to provide protection against mpox [1]. If initially allocated to this vaccination class (see Section S1.2.1), individuals remain here for the entire period considered by the model as we have assumed that limited vaccination resources would not be allocated to those with prior protection.
For the vaccination classes relating to the current vaccination period ($j = 2, 3, 4$), the population may only progress through the classes sequentially (individuals may only receive a first dose if they are unvaccinated, and individuals may only receive a second dose if they have received a first dose). We do not incorporate waning of protection given the relatively short-time period of the analysis in comparison to orthopoxvirus antibody duration [2].

**S1.1.1.1 Dose allocation** Let $V^{LC}(t)$, $V^{MVA1}(t)$, $V^{MVA2}(t)$ denote the available number of doses of: $LC16m8$, first doses of $MVA-BN$ and second doses of $MVA-BN$, on day $t$, respectively. Values for these parameters were varied across a range of plausible values, with details provided in Section S1.4).
Given $v^V(t)$, where $V \in \{LC, MVA1, MVA2\}$, vaccines are then allocated across a select number of groups each day if three criteria are met: eligibility, prioritisation and target. Individuals are eligible for a given vaccine depending on their age, and only if they are alive and non-infectious (e.g. those in susceptible, exposed, or recovered compartments). Due to the infeasibility of mass vaccination, we considered different prioritisation strategies in which high-risk groups are vaccinated ahead of the rest of the population, for example, young children or SWs. Further details are provided in Vaccination scenarios. Finally, using estimates of mpox vaccine hesitancy, we implement an upper limit of the percentage of people within each group who may accept a mpox vaccine if offered, given recent research from the region. Vaccines may only be allocated to this group until this limit is met (see Section S1.4.3).

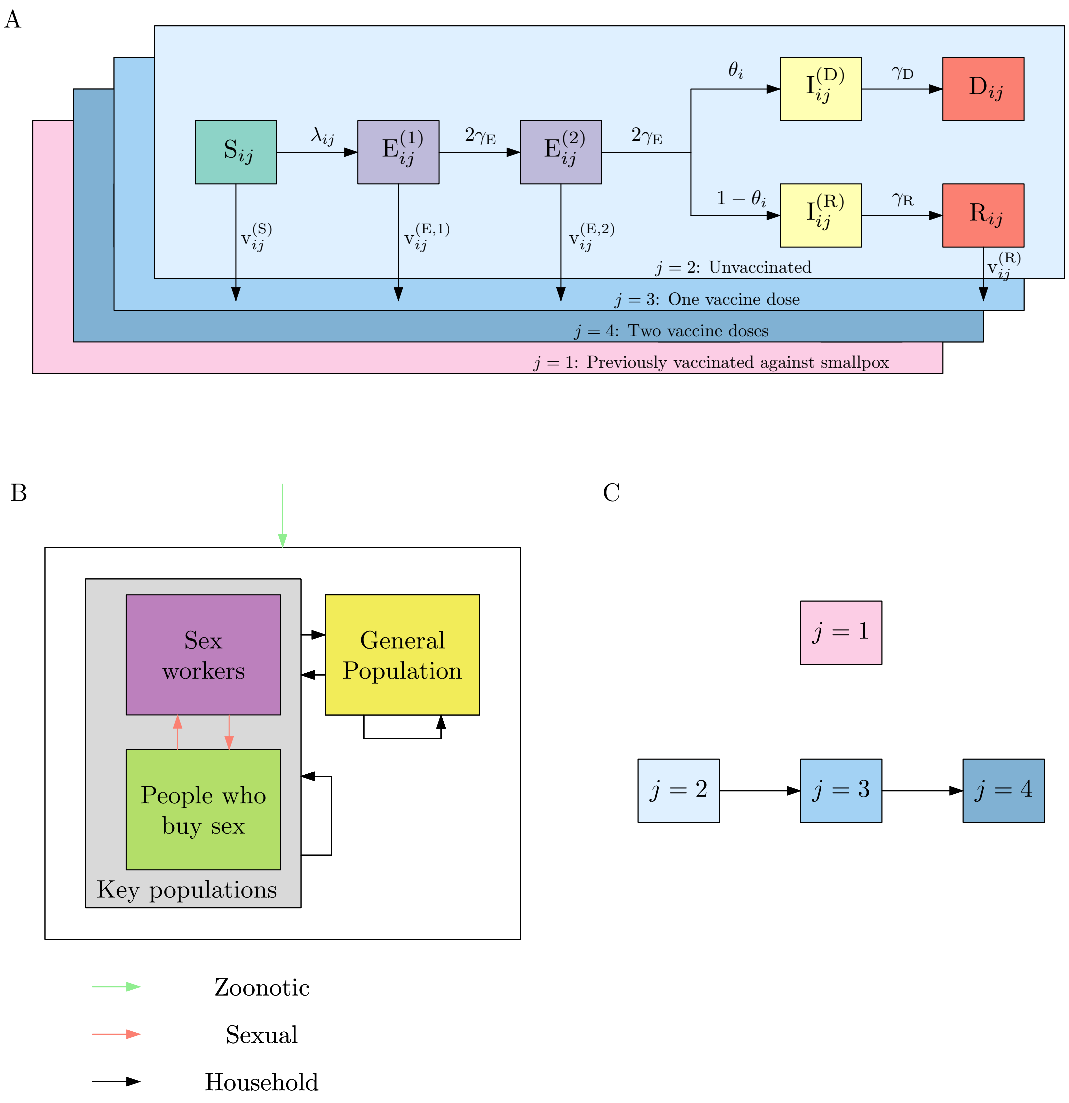

Figure S1: Overview of the mathematical model of mpox disease transmission, progression and vaccination presented in this manuscript. (A) The population moves through the different disease states susceptible ($S$), exposed ($E_a$,$E_b$), infected ($I_R, I_D$) and recovered ($R$) or dead ($D$). The disease states follow an identical flow regardless of vaccination stratum $j$ (although the likelihoods of becoming infected change). (B) The model captures key populations including sex workers and people who buy sex, drawn from the general population. The coloured arrows below indicate the three routes of transmission that have been modelled (sexual, non-sexual and zoonotic). (C) Vaccine stratum $j = 1$ accounts for individuals who have had previous smallpox vaccination, with no one able to move in or out of $j = 1$. The remaining strata correspond to being unvaccinated ($j = 2$), vaccinated with a single dose of either MVA-BN or LC16m8 ($j = 3$) or vaccinated with two doses of MVA-BN ($j = 4$). The population may only move through the vaccine strata sequentially and cannot move backwards (e.g. going from being vaccinated to unvaccinated).

Bringing this together, and considering eligibility, prioritisation, and target as binary indicators, we have:

$$G_i(t) = \begin{cases} 1, & \text{group } i \text{ is eligible, prioritised and below target coverage at time } t \\ 0, & \text{otherwise.} \end{cases}$$

Alongside restriction to eligible, prioritised demographic groups with unsaturated vaccine coverage (demographic groups $i$ such that $G_i(t) = 1$), vaccination is also restricted to one specific vaccine stratum $j$ at each timestep. Vaccination is in either a first dose cycle (targeting vaccine stratum $j = 2$) or a second dose cycle (targeting stratum $j = 3$). Similarly to the demographic group indicator variable $G_i(t)$, we define the vaccine stratum indicator variable:

$$H_j(t) = \begin{cases} 1, & \text{vaccine stratum } j \text{ is targeted for vaccination at time } t \\ 0, & \text{otherwise.} \end{cases}$$

At each time step, vaccines are allocated using a dual-step multinomial draw. First, for each vaccine type $V$, the $v^V(t)$ vaccines are allocated to each qualified group $i$ proportionally to the size of that group compared to other qualified groups:

$$v_i^V(t) \sim Multinomial\left( v^V(t),\ \frac{\sum_{X \in \{S, E_a, E_b, R\}} \left[X_{ij}(t) G_i(t)\right]}{\sum_i \sum_{X \in \{S, E_a, E_b, R\}} \left[X_{ij}(t) G_i(t)\right]} \right)$$

Second, within a given group $i$, and for each vaccine type $V$, vaccines are allocated across the relevant compartments $(S, E_a, E_b, R)$ proportionally to the number of individuals of the eligible vaccine stratum in each compartment under consideration.

$$v_{ij,X}^V(t) \sim Multinomial\left( v_i^V(t),\ H_j(t) \cdot \frac{X_{ij}(t)}{\sum_{\Xi \in \{S, E_a, E_b, R\}} \Xi_{ij}(t)} \right).$$

Changes to each state are then given as:

$$\delta_{ij,X}^V(t) = \begin{cases} 0, & j = 1 \\ -v_{ij,X}^V(t), & j = 2 \\ -v_{i,j-1,X}^V(t) + v_{ij,X}^V(t), & j = 3 \\ v_{i,j-1,X}^V(t), & j = 4 \end{cases}$$

for $V \in \{LC, MVA1, MVA2\}$ and $X \in \{S, E_a, E_b, R\}$ such that:

$$\delta_{ij,X}(t) = \sum_V \delta_{ij}^V(t).$$

###### S1.1.2 State transitions: Transmission and disease progression

Each state variable is a random variable with transitions between states sampled from the binomial distribution at each time step. Letting $n^{XX'}$ denote the number of people transitioning from state $X$ to state $X'$, we have:

$$\begin{aligned}
n_{ij}^{SE_a}(t) &\sim Binomial(S_{ij}(t) + \delta_{ij,S}(t), 1 - e^{-\lambda_{ij}(t)dt}) \\
n_{ij}^{E_a E_b}(t) &\sim Binomial(E_{a_{ij}}(t) + \delta_{ij,E_a}(t), 1 - e^{-2\gamma_E dt}) \\
n_{ij}^{E_b I}(t) &\sim Binomial(E_{b_{ij}}(t) + \delta_{ij,E_b}(t), 1 - e^{-2\gamma_E dt}) \\
n_{ij}^{E_b I_R}(t) &\sim Binomial(n_{ij}^{E_b I}(t), 1 - \theta_i) \\
n_{ij}^{E_b I_D}(t) &= n_{ij}^{E_b I} - n_{ij}^{E_b I_R} \\
n_{ij}^{I_R R}(t) &\sim Binomial(I_{R_{ij}}(t), 1 - e^{-\gamma_{I_R} dt}) \\
n_{ij}^{I_D D}(t) &\sim Binomial(I_{D_{ij}}(t), 1 - e^{-\gamma_{I_D} dt})
\end{aligned}$$

The force of infection incorporates infections coming from: zoonotic spillover (Z); non-sexual human-to-human (H); sexual human-to-human (S); and is defined by:

$$\lambda_{ij}(t) = (\beta^{\mathrm{H}} \sum_k m^{\mathrm{H}}_{ik} \frac{\sum_r I_{kr}(t)}{\sum_r N_{kr}(t)} + \mathbb{1}_{\{i \in \{\mathrm{SW},\mathrm{PBS}\}\}}(i)\beta^{\mathrm{S}} \sum_k m^{\mathrm{S}}_{ik} \frac{\sum_r I_{kr}(t)}{\sum_r N_{kr}(t)} + \mathbb{1}_{\{region=Equateur\}}\beta^{\mathrm{Z}}_i) \times (1 - ve_{ij})$$

where $\beta^{\mathrm{H}}$ captures the non-sexual human-to-human transmission rate; $\beta^{\mathrm{S}}$ captures the sexual human-to-human transmission rate; $\beta^{\mathrm{Z}}_i$ represents the (age-stratified) zoonotic spillover rate; $m^{\mathrm{H}}_{ik}$, $m^{\mathrm{S}}_{ik}$ represent the non-sexual ($H$) and sexual ($S$) rate of contact between groups ($i, k \in \{1, \ldots, 18\}$) which is assumed to be independent of vaccination status; and $ve_{ij}$ represents vaccine efficacy against infection for demographic group $i$ and vaccination status $j$ (hence in the unvaccinated group, $j = 2, ve_{i2} = 0$). $\mathbb{1}$ defines an indicator function which takes the value 1 when the condition in the subscript is met and 0 otherwise. Note that we assume that zoonotic spillover is only applicable to Equateur province (Clade Ia).
The length of stay in each disease state after becoming infected are represented by the variables $\gamma_X$ for $X \in \{E, I_R, I_D\}$, and the case fatality ratio (CFR) is represented by $\theta$. Due to only one observed death in Burundi, we do not model deaths in Bujumbura.
Bringing both the vaccination and disease progression steps together, the model can be described via the following equations:

$$\begin{aligned}
S_{ij}(t+1) &= S_{ij}(t) - n^{SE_a}_{ij}(t) + \delta_{ij,S}(t) \\
E_{a_{ij}}(t+1) &= E_{a_{ij}}(t) + n^{SE_a}_{ij}(t) - n^{E_aE_b}_{ij}(t) + \delta_{ij,E_a}(t) \\
E_{b_{ij}}(t+1) &= E_{b_{ij}}(t) + n^{E_aE_b}_{ij}(t) - n^{E_bI}_{ij}(t) + \delta_{ij,E_b}(t) \\
I_{R_{ij}}(t+1) &= I_{R_{ij}}(t) + n^{E_bI_R}_{ij}(t) - n^{I_RR}_{ij}(t) \\
I_{D_{ij}}(t+1) &= I_{D_{ij}}(t) + n^{E_bI_D}_{ij}(t) - n^{I_DD}_{ij}(t) \\
R_{ij}(t+1) &= R_{ij}(t) + n^{I_RR}_{ij}(t) + \delta_{ij,R}(t) \\
D_{ij}(t+1) &= D_{ij}(t) + n^{I_DD}_{ij}(t)
\end{aligned}$$

The model was written in *odin2* (v0.3.24) [3], fitted using *monty* (v0.3.31) [4] and is available in the package *mpoxseir* (v0.2.28) [5].

#### S1.2 Model fitting and parameter estimation

We first describe the non-fitted model parameters and inputs, before then outlining the fitting and parameter estimation process for the regions of Equateur, Sud Kivu, and Bujumbura.

##### S1.2.1 Parameterisation: non-fitted parameters

Table S1 provides an overview of the non-fitted model parameters, each of which are expanded upon in the following sections.

**S1.2.1.1 Demography** Age-stratified estimates of the total population of the Democratic Republic of the Congo (DRC) and Burundi were obtained from UN country-level estimates [6], with province-level population sizes sourced from UN estimates [7] for DRC and the National Ministry of Health for Burundi [8]. We assumed that the group structure observed nationally also held provincially, and age-stratified estimates of Equateur, Sud Kivu, and Bujumbura were taken by applying the national-level proportions to the estimated population of each province.
Population sizes for SW and PBS were calculated by multiplying the model parameters representing the proportion of the general population in these groups, $p_{\mathrm{SW}}$ and $p_{\mathrm{PBS}}$ respectively, by the total population size of each of the relevant age groups. After allocation to a key population, individuals were removed from the corresponding age class in the general population, to prevent double counting of the population.
In all three regions, SWs were assumed to be aged 12 – 49 years old [16]. We estimated the total SW population in each province by applying the relevant percentage to the female population (assumed to be 50% of the total population) in relevant age groups and removed this value from the corresponding

Table S1: Values, stratifications and sources of the fixed model parameters and inputs.

| **Parameter** | **Interpretation** | **Stratification(s)** | **Value(s)** | **Source(s)** |
|---|---|---|---|---|
| $N$ | Population size | Region; group $i$; vaccination strata $j$ | Table S2<br>Table S3 | [6, 7, 8, 9, 10] |
| $M^{\mathrm{H}}_{i_1 i_2}$ | 'General' contacts between groups $i_1$ and $i_2$ | NA | NA | [11] |
| $\frac{1}{\gamma_E}$ | Incubation period | No | 7 days | [12] |
| $\frac{1}{\gamma_{I_R}}$ | Average time from infection to recovery | No | 18 days | [13] |
| $\frac{1}{\gamma_{I_D}}$ | Average time from infection to death | No | 10 days | [13] |
| $\theta_{ij}$ | Case fatality ratio | Age; vaccination strata $j$ | Table S5 | [14] |
| $ve_{ij}$ | Vaccine efficacy against infection | Vaccine used | $LC16m8$:<br>95% (expert guidance)<br>95% 1 dose $MVA-BN$<br>95% 73.6%<br>95% 2 doses $MVA-BN$<br>95% 81.8% | [15] |

age groups. The parameter $p_{\mathrm{SW}}$ was calibrated in model fitting, as discussed in Section S1.2.2.4.
In all three regions, PBS were assumed to be those aged between 20–49 years old. To fix the value of $p_{\mathrm{PBS}}$, we used data from the Demographic and Health Surveys (DHS) Program that estimated that 11% of men aged 15–49 years old in the DRC have paid for sexual intercourse in the previous 12 months [10]. We assumed this was applicable to Equateur and Sud Kivu regions, and in the absence of setting-specific data, also assumed this applied to Bujumbura. This produced estimates of the number of PBS of $32,105$ in Equateur; $123,113$ in Sud Kivu and $21,870$ in Bujumbura.
To account for protection from previous smallpox vaccination, we assumed a percentage of each group aged 40 years and over in DRC were already vaccinated as estimated by Taube et al. [1]. We used the same figures for Burundi but assumed that only those aged 55 and over would have been previously vaccinated, after discussion with in-country experts. We used a binomial distribution to split the population in age groups aged 40 years or 55 years and over, depending on region, across the unvaccinated and vaccinated classes. We assumed key populations were drawn from the unvaccinated population.
The age- and vaccine-stratified population estimates nationally and per region are presented in Table S2 and Table S3.

**S1.2.1.2 Social contacts and mixing** Mixing patterns between population groups were represented using two contact matrices: (i) $M^{\mathrm{H}}$ which represents 'general' person-to-person contact e.g. within households, the community, and non-commercial sexual contacts, and (ii) $M^{\mathrm{S}}$ which represents commercial sexual contacts, i.e. between SW and PBS.
To parameterise $M^{\mathrm{H}}$ , we assumed that the populations of DRC and Burundi followed the same contact pattern as estimated by Melegaro et al. [11] for Manicaland Province in Zimbabwe, due to the absence of setting-specific data. To extend the age-stratified contact matrix to the key populations (SW and PBS) we first computed the overall number of contacts implied between each combination of age groups, $N_i\ m^{\mathrm{H}}_{ij}$ . For each age group containing key populations, we split the total number of general contacts according to the proportions of age group $i$ and $j$ that are part of key populations. We then summed the total contacts between each key population and each age group in the general population, as well as the number of 'general' contacts within and between each key population, ensuring that the total number of contacts between each pair of groups was balanced. Finally, we converted the matrix back into per-capita contact rates according to the population size of each group.
Due to the absence of information around contact rates and mpox transmissibility within commercial sexual networks, we varied the average number of PBS infected by a single infectious SW ($R_0^{(\mathrm{SW},\mathrm{PBS})}$) in model calibration. The parameterisation of $M^{\mathrm{S}}$ was then determined based on (i) the assumption

Table S2: Age- and historic-smallpox-vaccination-stratified estimates of the population of DRC overall in addition to Equateur and Sud Kivu. Sex workers and people who buy sex were drawn from the unvaccinated population in groups encompassing ages 12–49 and ages 20–49, respectively. As $p_{SW}$ is fitted in calibration, the exact population size varied per simulation.

| **Group** ($i$) | **DRC population** | **Smallpox vaccination (%)** | **Equateur** | | **Sud Kivu** | |
|---|---|---|---|---|---|---|
| | | | **Unvaccinated** | **Vaccinated** | **Unvaccinated** | **Vaccinated** |
| 0–4 | 15,827,439 | 0 | 302,547 | 0 | 1,160,178 | 0 |
| 5–11 | 18,270,871 | 0 | 349,255 | 0 | 1,339,285 | 0 |
| 12–14 | 6,916,506 | 0 | 132,212 | 0 | 506,991 | 0 |
| 15–19 | 9,472,510 | 0 | 181,071 | 0 | 694,351 | 0 |
| 20–24 | 7,729,713 | 0 | 147,756 | 0 | 566,601 | 0 |
| 25–29 | 6,439,523 | 0 | 123,094 | 0 | 472,028 | 0 |
| 30–34 | 5,340,933 | 0 | 102,094 | 0 | 391,499 | 0 |
| 35–39 | 4,421,401 | 0 | 84,517 | 0 | 324,096 | 0 |
| 40–44 | 3,629,088 | 46 | 37,461 | 31,910 | 143,650 | 122,368 |
| 45–49 | 2,967,441 | 71 | 16,500 | 40,396 | 63,272 | 154,906 |
| 50–54 | 2,421,737 | 71 | 13,425 | 32,867 | 51,480 | 126,037 |
| $55-59$ | 1,925,266 | 77 | 8,465 | 28,337 | 32,459 | 108,666 |
| $60-64$ | 1,486,411 | 79 | 5,967 | 22,446 | 22,881 | 86,075 |
| 65–69 | 1,099,430 | 79 | 4,413 | 16,603 | 16,924 | 63,666 |
| 70–74 | 789,010 | 79 | 3,167 | 11,915 | 12,146 | 45,690 |
| 75+ | 815,125 | 79 | 3,272 | 12,309 | 12,548 | 47,202 |
| Total | 89,561,404 | NA | 1,515,216 | 196,783 | 5,810,389 | 754,610 |

Table S3: Population size stratified by historic smallpox vaccinations status for Burundi overall in addition to Bujumbura. Sex workers were drawn from the unvaccinated population in groups encompassing ages 12–49 and people who buy sex were drawn from the unvaccinated population in groups encompassing ages 20–49. As $p_{SW}$ is fitted in calibration, the exact population size varied per simulation.

| **Group** ($i$) | **Burundi population** | **Smallpox vaccination (%)** | **Bujumbura** | |
|---|---|---|---|---|
| | | | **Unvaccinated** | **Vaccinated** |
| 0–4 | 2,053,840 | 0 | 189,186 | 0 |
| 5–11 | 2,421,951 | 0 | 223,094 | 0 |
| 12–14 | 905,016 | 0 | 83,364 | 0 |
| 15–19 | 1,216,932 | 0 | 112,096 | 0 |
| 20–24 | 1,042,770 | 0 | 96,053 | 0 |
| 25–29 | 986,811 | 0 | 90,899 | 0 |
| 30–34 | 875,169 | 0 | 80,615 | 0 |
| 35–39 | 664,378 | 0 | 61,198 | 0 |
| 40–44 | 446,705 | 0 | 41,148 | 0 |
| 45–49 | 300,927 | 0 | 27,719 | 0 |
| 50–54 | 259,788 | 0 | 23,930 | 0 |
| $55-59$ | 232,186 | 77 | 4,919 | 16,468 |
| $60-64$ | 201,291 | 79 | 3,894 | 14,648 |
| 65–69 | 138,340 | 79 | 2,676 | 10,067 |
| 70–74 | 70,370 | 79 | 1,361 | 5,121 |
| 75+ | 74,307 | 79 | 1,437 | 5,408 |
| Total | 11,890,781 | NA | 1,043,589 | 51,712 |

that the probability of transmission between SW and PBS is symmetric, conditional on one party being infected; and (ii) ensuring that the overall number of sexual contacts is balanced between the two groups.
The average number of PBS infected by a single infectious SW is given by:

$$R_0^{(\mathrm{SW,PBS})} = \beta^{\mathrm{S}} M^{\mathrm{S}}_{(\mathrm{SW,PBS})} \tau_{\mathrm{SW}}$$

where $\tau_i$ is the duration of infectiousness in group $i$. If partnerships balance, we know that:

$$M^{\mathrm{S}}_{(\mathrm{SW,PBS})} N_{\mathrm{SW}} = M^{S}_{(\mathrm{PBS,SW})} N_{\mathrm{PBS}}$$

Therefore, the transmission rate matrix is given by:

$$\beta^S M^S_{ik} = \begin{cases} \frac{R_0^{(\mathrm{SW,PBS})}}{\tau_{\mathrm{SW}}}, & i = \mathrm{SW}, k = \mathrm{PBS} \\ \frac{R_0^{(\mathrm{SW,PBS})}}{\tau_{\mathrm{SW}}} \frac{N_{\mathrm{SW}}}{N_{\mathrm{PBS}}}, & i = \mathrm{PBS}, k = \mathrm{SW} \\ 0, & \text{otherwise} \end{cases}$$

**S1.2.1.3 Basic reproduction number** We calculated the basic reproduction number $R_0$ for our model, defined as the average number of secondary infections generated by a single infectious individual in a wholly susceptible population, using next generation matrix (NGM) methods [17]. In addition to the overall $R_0$, we calculated transmission-route specific reproduction numbers, corresponding to transmission in the general population $R_0^{\mathrm{H}}$, and via commercial sexual networks $R_0^{\mathrm{S}}$. In each case, the basic reproduction number was defined as the dominant eigenvalue of the NGM. Each entry i,k in the NGM represents the average number of infections generated in group i by an infectious individual in group $k$.
To construct the NGM, we calculated the average duration of infectiousness for individuals in each population group $i$:

$$\tau_i^I = \frac{1-\theta_i}{\gamma_{I_R}} + \frac{\theta_i}{\gamma_{I_D}}$$

The NGM was then calculated for each transmission route $r \in \{H, S\}$:

$$NGM^r_{ik} = \beta^r M^r_{ik} \tau^I_k$$
$$i, k \in \{[0,5), ..., \{75+\}, \mathrm{SW}, \mathrm{PBS}\}$$

Giving an overall NGM of:

$$NGM_{ik} = NGM^{\mathrm{H}}_{ik} + NGM^{\mathrm{S}}_{ik}$$

We calibrated transmissibility in the general population by varying the basic reproduction number for this transmission route, $R_0^{\mathrm{H}}$, then calculating the corresponding value of $\beta^t extH$ , i.e.

$$\beta^{\mathrm{H}} = \frac{R_0^{\mathrm{H}}}{ev(M^{\mathrm{H}} \boldsymbol{\tau}^I)}$$

where $ev()$ represents eigenvalue.

**S1.2.1.4 Zoonotic transmission by age in Equateur** The risk of zoonotic spillover of mpox Clade Ia varies by both region and age, with young children in endemic provinces most likely to be infected after contact with animal reservoirs [18]. The force of infection in our model incorporates parameter $\beta_i^Z$ to describe the age-stratified zoonotic transmission rate in endemic regions, i.e. Equateur. This parameter can be represented as $\beta_i^{\mathrm{Z}} = \beta^{\mathrm{Z}}\ \psi_i$, where $\psi_i \in [0,1]$ represents the relative risk of zoonotic transmission in each age group compared to the maximum, and $\beta^{\mathrm{Z}} = max_i\{\beta_i^{\mathrm{Z}}\}$ is the maximum per-capita rate of zoonotic transmission across all age groups.
In Equateur, we calibrated the expected number of daily zoonotic introductions $\zeta$, which was related to the age stratified rates via the following equation:

$$\zeta = \sum_i \beta_i^{\mathrm{Z}} = \beta^{\mathrm{Z}} \sum_i \psi_i$$

Table S4: Calculations underpinning estimated age-stratified transmission coming from zoonosis.

| Age group $a$ | Observed zoonotic cases $C_a$ | Proportion of cases vaccinated $p_a^{vax}$ | Estimated vaccinated cases $C_a^{vax}$ | Estimated unvaccinated cases $C_a^U$ | Estimated total cases without vaccination $C'_a$ | DRC population 1983 (millions) $N_a$ | Cases per $100,000$ $\sigma_a$ | Relative risk $\psi(a)$ |
|---|---|---|---|---|---|---|---|---|
| Source | [18] | [13] | $p_a^{vax}C_a$ | $C_a - C_a^{vax}$ | $\frac{C_a^{vax}}{1-ve_{i1}} + C_a^U$ | [19] | $\frac{C'_a}{N_a}$ | $\frac{\sigma_a}{max_a\{\sigma_a\}}$ |
| $0-4$ | 130 | 0.7% | 0 | 130 | 130 | 5.3 | 2.5 | 1.000 |
| $5-11$ | 93 | 14.3% | 13 | 80 | 130 | 5.5 | 2.2 | 0.853 |
| $12-14$ | 13 | 85.0% | 3 | 10 | 21 | 2.1 | 1.1 | 0.450 |
| 15+ | 9 | 85.0% | 7 | 2 | 29 | 16.1 | 0.2 | 0.079 |
| Total | 245 | NA | 23 | 222 | 310 | 29.0 | NA | NA |

Details of the calibration and prior estimate can be found in the main text and Section S1.2.2.4.

We parameterised the relative levels of zoonotic transmission in each age group, $\psi_i$, using data from historic epidemiological investigations into Clade Ia outbreaks in central DRC during the 1980s [18], adjusted for population-level smallpox vaccine coverage at that time [13].

In their 1988 paper [18], Jezek and colleagues report total numbers of zoonotic cases in age group $a$, which we denote $C_a$, and separately in their 1987 paper [13], report the proportion of cases that are vaccinated in each age group, which we denote $p_a^{vax}$. We estimated the number of cases among vaccinated individuals for each age group, $C_a^{vax} = p_a^{vax}C_a$, subject the constraint $\sum_a C_a^{vax} = 23$ [13]. We then calculated the number of cases among unvaccinated individuals $C_a^U = C_a - C_a^V$. We inferred how many cases would have occurred in a completely unvaccinated population by calculating: $C'_a = \frac{C_a^{vax}}{1-vei1} + C_a^U$ where $ve_{i1} = 73.6\% \; \forall \; i$ is the efficacy of first-generation smallpox vaccines against mpox infection estimated by Berry et al. [15] in their meta-analysis. We adjusted for the population age-distribution to obtain a weighted relative risk of infection per capita in each age group: $\sigma_a = \frac{C'_a}{N_a}$, which we scaled relative to the group most at risk: $\psi_a = \frac{\sigma_a}{max_a\{\sigma_a\}}$. This is reported in Table S4.

**S1.2.1.5 Epidemiological delays** The mean incubation period, $\frac{1}{\gamma_E}$, was taken as reported by Besombes et al. [12] in their analysis of Clade I mpox patients in Cameroon. Data on the mean time to recovery, $\frac{1}{\gamma_{I_R}}$, and to death, $\frac{1}{\gamma_{I_D}}$, were taken as the mean of gamma distributions fitted to data extracted from Figures 1 and 2 in Jezek et al. [13]. The three corresponding values used in the analysis can be found in Table S1.

**S1.2.1.6 Severity** The relationship between the CFR ($\theta_i$) and age was quantified by Whittles et al. [14]. The CFR for age group 40–44 was assumed to be applicable to all older age groups, as there are not specific point estimates. For the SW and PBS populations, the CFR is assumed to be the weighted average of those for the relevant age groups (12–49 for SWs; 18–49 for PBS). These estimates can be found in Table S5. While there is some evidence to suggest that vaccination may protect against severe disease and death in breakthrough infections [14], the level of protection has not been robustly quantified, so we conservatively assumed no additional protective effect of vaccination beyond that conferred against infection.

We fitted the CFR in the youngest age group,$\theta_{[0,5)}$, in Equateur and Sud Kivu and used the estimates from Whittles et al. [14] to infer $\theta_i \; \forall \; i \geq 2$ based on this, assuming that the age structure of the CFR was unchanged.

**S1.2.1.7 Initial model conditions** In Sud Kivu and Bujumbura, the initial number of infectious individuals at the start of the epidemic is simulated according to a Poisson distribution, such that:

$$\sum E_{a,ij}(0) = \begin{cases} \sim Poisson(\mu) & i = \text{SW}; j = 2 \\ 0 & \text{otherwise} \end{cases}$$

We assume that all initially infectious individuals are unvaccinated sex workers. In Sud Kivu, $\mu$ is fitted to reflect the uncertainty surrounding the beginning of the epidemic in this region, with phylogenetic

Table S5: Estimated case fatality ratio (CFR) from Whittles et al. [14] used to inform the age-stratified CFR.

| Group ($i$) | CFR among unvaccinated (%) |
|---|---|
| 0–4 | 10.2 |
| 5–11 | 5.4 |
| 12–14 | 3.5 |
| 15–19 | 2.6 |
| 20–24 | 2.0 |
| 25–29 | 1.6 |
| 30–34 | 1.3 |
| 35–39 | 1.2 |
| 40–44 | 1.0 |
| 45–49 | 1.0 |
| 50–54 | 1.0 |
| 55 − 59 | 1.0 |
| 60 − 64 | 1.0 |
| 65–69 | 1.0 |
| 70–74 | 1.0 |
| 75+ | 1.0 |
| SW | 1.8 |
| PBS | 1.5 |

evidence showing circulation of Clade Ib ahead of the first reported cases [20, 21].
In Bujumbura, epidemiological evidence [22] shows the epidemic stemmed from Sud Kivu, and so rather than fitting this parameter we assumed an arbitrarily small number ($\mu = 10$) of cases based on the approximate date of introduction constrained by epidemiological evidence.
In Equateur, where mpox is enzootic, we assume that the initial number of infectious individuals is distributed according to the expected proportion of zoonotic importation in that group:

$$E_{a,ij}(0) = \begin{cases} \sim Poisson(\frac{\psi_i}{\sum_i \psi_i}) & j = 2 \\ 0 & \text{otherwise} \end{cases}$$

Despite being seeded only into the incubating compartment, infections quickly propagate through the population and after a few generations of transmission. Cases then follow patterns driven by the structure of the ongoing seeding (in endemic areas) and contact matrices.

**S1.2.1.8 Vaccine effectiveness** Estimates of vaccine effectiveness against infection were based on a meta-analysis linking antibody titres to vaccine-derived protection by Berry et al. [15]: 73.6% for historic smallpox vaccination or one dose of MVA-BN, rising to 81.8% after two doses. For the single dose of LC16m8, experts at WHO advised using vaccine efficacy of 95%, in line with data showing high seroconversion rates after vaccination [23] and in the absence of real-world effectiveness studies.
We assumed that it takes 14 days from receiving a vaccine dose to achieving the maximum levels of protection against infection as described above [24]. This was incorporated within the model by shifting forward the date of vaccination of by 14 days.

##### S1.2.2 Calibration data and observation model

To describe the epidemic in each region, we fitted our model to time series data on weekly cases and deaths, the CFR in each age group, and the proportion of cases reported among sex workers. Table S6 describes the datasets used to calibrate the model to the regional epidemics.
The data available for model calibration differed between the countries considered, as detailed in Appendix Table S6, we therefore used bespoke likelihoods for the DRC regions (Sud Kivu, Equateur) and Bujumbura.

###### S1.2.2.1 Observation process in DRC

Table S6: Overview of the data sources and definitions used in model fitting for each region. Data for DRC came from the Integrated Disease Surveillance and Response (IDSR), with the data for Burundi from the national surveillance system for mpox.

| | **Description** | **Age bands ($k$)** | **Time period and source** | **Location** |
|---|---|---|---|---|
| $Y_k^C(t)$ | Weekly cases by age | $[0,5),[5,15),15+$ | 7 Jan - 19 May 2024 | DRC |
| | | | 18 Aug 2024 - 5 Jan 2025 | Burundi |
| $Y_k^D(t)$ | Weekly deaths by age | $[0,5),[5,15),15+$ | 7 Jan - 19 May 2024 | DRC |
| $Y^C(t)$ | Total weekly cases | All | 26 May 2024 - 9 Feb 2025 | DRC Bujumbura |
| $Y^D(t)$ | Total weekly deaths | All | 26 May 2024 - 9 Feb 2025 | DRC |
| $Y^B(t)$ | Total weekly cases in country | All | 8 Aug 2024 - 5 Jan 2025 | Burundi |
| $Y_{SW}^C(t)$ | Total weekly cases in SW | All | 7 Jan - 31 Mar 2024 | DRC |
| $Y^{C_L}(t)$ | Total weekly cases in the line-list | All | 7 Jan - 31 Mar 2024 | DRC |
| $Y_k^{CFR}(t)$ | Cumulative CFR by age | $[0,5),[5,15),15+$ | 9 Feb 2025 | DRC |

**S1.2.2.1.1 Overview of mpox data and surveillance in DRC** The surveillance system for mpox in the DRC integrates two complementary data streams: aggregated national surveillance data and individual case investigation data. Suspected mpox cases are first identified by healthcare providers across 9,621 health areas, where basic epidemiological and clinical information is collected during the initial investigation and sampling.

For the aggregated surveillance data, health zones compile weekly line lists containing minimal epidemiological information such as age, gender, health area, and outcomes. These line lists are sent from the 519 Health Zone Central Offices to the 26 Provincial Health Divisions, which then produce weekly aggregated and quality-controlled data reports. These reports are forwarded to the Directorate of Epidemiological Surveillance (DSE), which compiles national-level aggregated data. This information is then transmitted to the National Institute of Public Health, where epidemiological situational reports are generated to inform public health decision-making.

Concurrently, the individual case investigation process documents detailed information about each suspected case at the time of sampling. Samples collected from suspected cases are shipped alongside individual case investigation forms to the National Reference Laboratories (INRB) located in Kinshasa and Goma and decentralized laboratories. The INRB compiles and maintains an individual-level laboratory results database that includes key variables such as age, gender, province, health zone, illness onset date, dates of sample collection and reception, and PCR results. Although the full content of the case investigation forms is not entered into a database, the lab database with minimal epidemiological data is pseudonymized and shared with both the Directorate of Epidemiological Surveillance (DSE) and the National Institute of Public Health.

Ultimately, aggregated surveillance data and individual-level laboratory results are consolidated and provided to the study team for comprehensive analysis. This integrated approach ensures both a broad overview of national mpox trends and the detailed epidemiological information needed for in-depth investigation of individual cases.

**S1.2.2.1.2 Surveillance data time series** In the DRC regions, where time series data on cases ($Y_k^C(t)$) and deaths ($Y_k^D(t)$) in age group $k$ and week $t$ were available up to time $T$ =26 May 2024, we represented the data as the observed realisations of underlying hidden Markov processes, defined as:

$$W_k^C(t) = \phi_k \sum_{i \in k} \sum_j n_{ij}^{SE_a}(t)$$
$$W_k^D(t) = \phi_k \sum_{i \in k} \sum_j n_{ij}^{I_D D}(t)$$

where $\phi_k, k \in \{[0,5), [5,15), 15+\}$ represent the case-ascertainment rates in each of the age tranches used in the surveillance system.
We related the modelled processes, $W_k^l(t)$, $l \in \{C, D\}$, to the data, $Y^l(t)$, via the reporting distributions:

$$Y_k^l(t) \sim NegBinom(W_k^l(t), \kappa_l)$$

so that $\mathbb{E}[Y_k^l(t)] = W_k^l(t)$ and $var[Y_k^l(t)] = W_k^l(t) + \frac{(W_k^l(t))^2}{\kappa^l}$.
We allowed for overdispersion in the observation process, (i.e. for the variance to be greater than the mean) via the parameters $\alpha^l = \frac{1}{\kappa^l} : l \in \{C, D\}$. We varied the overdispersion parameters in calibration, as detailed in the main text.
Where age-stratified time series data were unavailable, i.e. after $T =$26 May 2024 (see Table S6) we used aggregated time series of the total numbers of cases and deaths $Y^l(t)$, which were compared to modelled estimates in the same way $W^l(t) = \sum_k W_k^l(t)$.
The contribution to the overall likelihood function from weekly cases and deaths was therefore:

$$\mathbb{L}^C = (\Pi_{t \geq T} f_{Y^C}(Y^C(t)|W^C(t), \kappa^C))(\Pi_k \Pi_{t<T} f_{Y^C}(Y_k^C(t)|W_k^C(t), \kappa^C))$$
$$\mathbb{L}^D = (\Pi_{t \geq T} f_{Y^D}(Y^D(t)|W^D(t), \kappa^D))(\Pi_k \Pi_{t<T} f_{Y^D}(Y_k^D(t)|W_k^D(t), \kappa^D))$$

**S1.2.2.1.3 Cumulative CFR by age** To ensure we replicated the observed severity by age later in the DRC epidemics, when age stratified time series data were not available, we compared the observed CFR in each age category in the surveillance data, $Y_k^{CFR}(t) \in [0,1], k \in \{[0,5), [5,15), \{15+\}\}$, to our model predictions:

$$Y_k^{CFR}(t) \sim Beta(\sum_s^t X_k^D(s), \sum_s^t X_K^C(s) - \sum_s^t X_k^D(s))$$

so that $\mathbb{E}[Y_k^{CFR}(t)] = \frac{\sum_s^t X_k^D(s)}{\sum_s^t X_k^C(s)}$.
The contribution to the overall likelihood function from the observed CFR is:

$$\mathbb{L}^{CFR} = \Pi_k f_{Y^{CFR}}(Y_k^{CFR}(t)|X_k^C(s), X_k^D(s); s = 1, ..., t).$$

**S1.2.2.1.4 Line-list data on cases** We compared the number of cases from the line-list whose occupations were recorded as SW in week $t$, $Y_S W^C(t)$, with that predicted by our model, allowing for the total number of cases recorded in the line-list that week, $Y^{C_L}(t)$.

$$Y_{SW}^C(t) \sim Binom(Y^{C_L}(t), \frac{W_{SW}^C(t)}{W^C(t)}$$

The contribution to the likelihood of the line-list data was:

$$\mathbb{L}^{SW} = \Pi_t f_{Y_{SW}^C}(Y_{SW}^C(t)|Y^C(t), W_{SW}^C(t), W^C(t)).$$

**S1.2.2.1.5 Overall likelihood** In the DRC epidemics, where deaths were recorded, the overall likelihood was therefore:

$$\mathbb{L} = \mathbb{L}^C \mathbb{L}^D \mathbb{L}^{CFR} \mathbb{L}^{SW}.$$

**S1.2.2.2 Observation process in Burundi** In Bujumbura, no data on the proportion of cases in SW was available, and there was minimal mortality, so the likelihood was based on case numbers alone:

$$\mathbb{L} = \mathbb{L}^C$$

While age-stratified cases numbers were available for Burundi as a whole (denoted $Y_k^B(t)$), only age-aggregated time series data on cases were available for the Bujumbura regions specifically (denoted $Y^C(t)$, as above).
Similarly to the DRC regions, we related the model-predicted weekly cases $W^C(t)$ to the data via the reporting distribution:

$$Y^C(t) \sim NegBinomial(W^C(t), \kappa_l).$$

Since mpox cases in Bujumbura represent the majority of the national burden (67%) we assumed that the proportion of cases in each age group over time observed nationally is mirrored within Bujumbura. We used nested Beta-Binomial distributions to represent this age distribution such that:

$$\begin{aligned} Y^B_{[0,15)}(t) &\sim BetaBinom(Y^B(t), \frac{W^C_{[0,15)}(t)}{W^C(t)}, \rho) \\ Y^B_{[0,5)}(t) &\sim BetaBinom(Y^B_{[0,15)}(t), \frac{W^C_{[0,5)}(t)}{W^C_{[0,15)}(t)}, \rho). \end{aligned}$$

We allowed for overdispersion in the observation process via the parameter $\rho$, which we varied in calibration.
Under this formulation (and equivalently for $Y^B_{[0,5)}(t)$):

$$\begin{aligned} \mathbb{E}[Y_{[0,15)}(t)] &= Y^B(t)\frac{W^C_{[0,15)}(t)}{W^C(t)} \\ var[Y_{[0,15)}(t)] &= \mathbb{E}[Y_{[0,15)}(t)](1 - \frac{W^C_{[0,15)}(t)}{W^C(t)})(1 + (Y^B(t) - 1)\rho). \end{aligned}$$

**S1.2.2.3 Likelihood estimation** An analytical expression for the likelihood of the observed data given the underlying model and its parameters was not tractable, so we used particle filtering methods to obtain an unbiased estimators, from which we could efficiently sample [25].

**S1.2.2.4 Parameters varied in calibration and prior distributions** Up to nine model parameters per region were fitted during calibration. We set prior distributions for the fitted parameters using information from the published literature where available, and weakly informative priors restricting the parameters to plausible bounds otherwise as set out in Table S7 and described below.
Flat, uninformative priors were placed on: $R_0^{\mathrm{H}}$ and $R_0^{\mathrm{SW,PBS}}$ (Equateur, Sud Kivu); $\theta_{[0,5)}$ (Equateur, Sud Kivu); $\phi_{[5,15)}$ and $\phi_{\{15+\}}$ (Equateur, Sud Kivu, Bujumbura); $\zeta$ (Equateur); $\mu$ (Sud Kivu).
Using the posterior draws for Sud Kivu, more informative priors were constructed for $R_0^{\mathrm{H}}$ and $R_0^{\mathrm{SW,PBS}}$ in Bujumbura. For $R_0^{\mathrm{H}}$ , the mean and standard deviation of the posterior draws of this parameter in Sud Kivu were taken to parameterise a Gamma distribution using method of moments. The prior on $R_0^{\mathrm{SW,PBS}}$ was arbitrarily narrowed based on the Sud Kivu results but remained uninformative and was minimally impactful on the results as all values fell comfortably below the upper limit.
Parameters related to overdispersion ($\alpha^C, \alpha^D, \rho$) were given Beta priors skewed towards 0, e.g. assuming a low level of overdispersion.
We constructed priors for $p_{\mathrm{SW}}$ in each region using the Beta distribution. Although $p_{\mathrm{SW}}$ was allowed to vary between 0 and 1, we subsequently multiplied the parameter by 0.5 to account for this proportion applying to the female population only (assumed to be roughly half of the total population).
Estimates from Laga et al. [26] suggest approximately 0.7% of women aged 15–49 years old in the DRC are SW which we apply to Equateur province, while reports suggest this is closer to 3% in Sud Kivu [27]. A report of key populations [9] suggested a lower and upper bound on the number of SWs in Bujumbura Mairie as $2,670$ and $4,543$, respectively. We took the midpoint of these two values,

Table S7: Prior distribution for fitted parameters per region.

| Fitted parameter | Description | Prior distribution | | |
|---|---|---|---|---|
| | | Equateur | Sud Kivu | Bujumbura |
| $p_{SW}$ | Proportion of the population engaging in sex work | $Beta(2, 283.71)$ | $Beta(2, 64.7)$ | $Beta(2, 109.11)$ |
| $R_0^{\mathrm{H}}$ | Basic reproduction number in general population | $U[0, 1000]$ | $U[0, 1000]$ | $Gamma($ $shape = 17.91,$ $scale = 0.02)$ |
| $R_0^{\mathrm{SW,PBS}}$ | Basic reproduction number from SW to PBS | $U[0, 1000]$ | $U[0, 1000]$ | $U[0, 10]$ |
| $\phi_{[5,15)]}$ | Level of case under-ascertainment in older children | $Beta(1, 1)$ | $Beta(1, 1)$ | $Beta(1, 1)$ |
| $\phi_{\{15+\}}$ | Level of case under-ascertainment in adults | $Beta(1, 1)$ | $Beta(1, 1)$ | $Beta(1, 1)$ |
| $\alpha^C$ | Overdispersion parameter for observation of cases | $Beta(1, 9)$ | $Beta(1, 9)$ | NA |
| $\alpha^D$ | Overdispersion parameter for observation of deaths | $Beta(1, 9)$ | $Beta(1, 9)$ | NA |
| $\rho$ | Overdispersion parameter for observation of cases in Bujumbura compared to Burundi as a whole | NA | NA | $Beta(1, 9)$ |
| $\theta_{[0,5)}$ | Case fatality ratio in the youngest age group | $Beta(1, 1)$ | $Beta(1, 1)$ | NA |
| $\zeta$ | Average weekly importations from the zoonotic reservoir in endemic regions | $U[0, 1000]$ | NA | NA |
| $\mu$ | Poisson mean of number of infections seeded in SWs in Sud Kivu | NA | $U[0, 1000]$ | NA |

3,852, as a central estimate corresponding to an estimate of 0.9% of women aged 15–49 years, which we assumed applied to the whole Bujumbura region. The Beta prior was parameterised to be centred on these values in each region.

**S1.2.2.5 Implementation of pMCMC** To sample from the posterior distribution of the model parameters and obtain fitted model trajectories compatible with the observed data, we used particle Markov chain Monte Carlo (pMCMC) methods. We implemented a particle marginal Metropolis–Hastings algorithm with a bootstrap particle filter and 400 particles using the R packages *dust2* (v0.3.23) [28] and *monty* (v0.3.31) [4]. The number of particles was chosen to balance the variance of the likelihood estimate with computational efficiency, making use of parallel processing across available CPU cores.

To avoid numerical issues when the model predicts zero expected counts where non-zero observations are recorded, leading to particles with zero weight and likelihood estimates of zero, we added a small amount of noise to modelled counts within the particle filter. Specifically, we perturbed model-predicted counts using exponentially distributed noise with a mean of $10^{-6}$, ensuring that each particle retained a small but non-zero weight at every observation time.

At each iteration, the sampler proposes an update to the joint distribution of parameters using multivariate Gaussian proposals centred on the current values. The proposal distribution's covariance matrix was tuned to encourage efficient mixing of the Markov chain. Reflecting boundaries were imposed to ensure that proposed parameter values remained within biologically and mathematically plausible ranges while preserving the symmetry of the proposal kernel.

#### S1.3 Estimation of effective reproduction number

We estimated the effective reproduction number, $R_t$, over the course of each epidemic, i.e. the average number of infections caused by an infectious individual at time $t$, accounting for the depletion of susceptibles over the course of the epidemic as individuals become immune following infection. Similarly to the calculation of $R_0$, the calculation of $R_t$ was based on the dominant eigenvalue of the NGM adjusted for the fraction of the population in each group $i$ that remained susceptible. Results are presented in Figure S19.

#### S1.4 Vaccination scenarios

##### S1.4.1 Rollout scenarios

In line with in-country considerations, we simulated scenarios dispensing: (1) Lc16m8 only; (2) a mixed strategy where both Lc16m8 and single-dose MVA-BN are deployed in parallel; and MVA-BN only, delivered under either (3) a single-dose, or (4) a two-dose regimen, where the latter strategy vaccinates half as many people, but offers higher individual-level protection.
LC16m8 vaccines are not given to SWs due to risks related to this vaccine for immunocompromised individuals. In scenarios using both vaccines, LC16m8 was given to those under 12, and MVA-BN to older age groups and key populations. Smallpox-vaccinated individuals were assumed ineligible for mpox vaccination.

##### S1.4.2 Prioritisation strategies

Prioritisation of risk groups was implemented in the model based on the work of Hogan et al. [29]. We ran scenarios in which all of the population were given the vaccine regardless of age or key population status, referred to as "no prioritisation".
Given the relationship between age and death conditional on mpox Clade Ia infection [14], we modelled prioritising those aged $0-11$ for vaccination until target levels were met, before vaccination then commenced in the general population. This is referred to as "prioritise $< 12$ years old". We modelled an analagous set of scenarios for prioritising SWs, which is referred to as "prioritise sex workers".

##### S1.4.3 Vaccine uptake and target coverage

Studies of vaccine hesitancy across the region suggest that not everyone who is offered a mpox vaccine will accept it [30, 31]. We used estimates of reported intention to take a vaccine if offered to inform the maximum number of individuals within a group $i$ that we assume could be vaccinated. In a given prioritisation strategy, these estimates serve as the target vaccination coverage required within each group $i$ before the campaign can move to the next stage.
A study by Petrichko et al. [30] conducted across late 2023 – early 2024 estimated mpox vaccine acceptance of 47.8% and 55.8% among adults in Equateur and Sud Kivu, respectively. A separate study by Du et al. [31] considering mpox vaccine hesitancy in six African countries considered acceptance in children separately to adults. Restricting the estimates in this analysis to Uganda and Kenya, the two countries most socioeconomically and demographically similar to DRC, the acceptance rate in children was 95% that of the acceptance rate in adults. An analogous analysis suggested individuals reporting risky sexual behaviour were 15% more likely to accept vaccination compared to the general adult population. Therefore, we adjusted estimated acceptance in adults in each province for children and key populations according to these rates. Values specific to Burundi were unavailable, thus in the absence of setting-specific evidence, were assumed equivalent to DRC at the national level using the same group adjustments for children and key populations.
Table S8 presents vaccine acceptance per region as incorporated in the model.

Table S8: Vaccine acceptance by region and group considered in the model. Children are assumed to be those aged under 18 years old. Due to the absence of setting-specific data, values for Bujumbura are assumed to be that of DRC nationally. These estimates are taken as vaccination targets within the relevant groups according to the prioritisation strategy under consideration.

| **Group** | **Acceptance relative to adult population** | **Source** | **Vaccine acceptance (%)** | | | |
|---|---|---|---|---|---|---|
| | | | **Equateur** | **Sud Kivu** | **Bujumbura** | **Source** |
| Adults | NA | | 47.8 | 55.8 | 61.0 | [30] |
| Children | 0.95 | [31] | 45.4 | 53.0 | 58.0 | [30, 31] |
| SW/PBS | 1.15 | [31] | 55.0 | 64.2 | 70.2 | [30, 31] |

###### S1.4.4 Dose supply

Fragmented and sparse reporting hindered our ability to obtain the true number of MVA-BN and LC16m8 vaccines purchased by, pledged to, or received by DRC. In addition, because Burundi has been responding to the outbreak with non-pharmaceutical interventions, instead we focussed on the potential impact, in terms of cases and deaths averted, under different plausible values for dose numbers in each region.

Reports suggested that DRC were expecting 3 million doses of LC16m8, and the wider region expected approximately 2.3 million doses of MVA-BN [32, 33]. In November 2024, the WHO reported that 85% of the recently donated doses were allocated to DRC due to their high case load [34]. Applying this percentage to the number of expected MVA-BN doses in the region would suggest an upper bound of approximately 2 million doses of this vaccine allocated to DRC. This suggests a ratio of 2 MVA-BN for every 3 LC16m8 vaccines, which is what we implemented in scenarios in which both vaccine types were modelled.

In DRC, we treated the 2 million MVA-BN vaccines pledged to the country as an upper bound on the number of doses that could be delivered in Sud Kivu (population $\sim$ 5.8 million). We then derived a population-scaled upper bound for Equateur (population $\sim$ 1.6m) of $520,000$ doses. In Burundi, in the absence of mpox vaccine pledges, we used the number of vaccines delivered during the COVID-19 pandemic as a proxy (upper bound $50,000$ doses). For each region, we explored progressively lower supply scenarios by reducing dose availability down to 0.5% of these upper bounds, corresponding to lower bounds of $10,000$ doses in Sud Kivu; $2,600$ doses in Equateur and 500 doses in Bujumbura. Below, we also conducted an additional analysis with up to $800,000$ available doses in Bujumbura to assess trends at higher vaccine coverages.

###### S1.4.5 Capacity to deliver vaccinations

The maximum number of vaccinations that can be delivered in region, $r$, on a given day is held constant throughout the vaccination period. We modelled seven different rollout speeds reflecting the number of doses that could be given per day, with a lower bound of 250 and an upper bound of 20,000 for both Equateur and Sud Kivu. Based on expert in-country guidance, we reduced the rollout speeds used in DRC by a factor of 10 for Bujumbura.

For low-dose scenarios, daily vaccination capacity was reduced to ensure a minimum rollout of one week; in some high-dose scenarios daily capacity constraints meant that not all pledged vaccines could be administered within the two-year time frame.

###### S1.4.6 Dosing strategy

The MVA-BN vaccine requires two doses to reach maximum effectiveness. In practise, a two-dose regime entails fully vaccinating only half of the number of people as there are available MVA-BN doses. Manufacturer guidance recommends a minimum of 28 days between first and second doses [35]. Here, we adopt a strict 28-day delay between doses for all scenarios in which two doses are administered, assuming that first doses are given for 28 days, followed by these individuals receiving second doses for the next 28 days, continuing in this pattern until all first and second vaccine doses have been given out, ensuring that everyone who receives a first dose then also receives a second dose. As WHO recommends giving one dose to more of the population at a lower effectiveness rather than giving fewer people two doses at maximum effectiveness [36], we modelled both scenarios to quantify

Table S9: The modelled values for the number of doses and daily vaccination delivery capacity (doses per day) for each region.

| Region | Total doses | Doses per day |
|---|---|---|
| Equateur | $\{2,600; 13,000; 26,000;$ $130,000; 260,000; 520,000\}$ | $\{250; 500; 1,000; 2,500;$ $5,000; 10,000; 20,000\}$ |
| Sud Kivu | $\{10,000; 50,000; 100,000;$ $500,000; 1,000,000; 2,000,000\}$ | $\{250; 500; 1,000; 2,500;$ $5,000; 10,000; 20,000\}$ |
| Bujumbura | $\{500; 2,500; 5,000; 25,000;$ $50,000; 100,000; 200,000;$ $400,000, 600,000, 800,000\}$ | $\{25; 50; 100; 250;$ $500; 1,000; 2,000\}$ |

the benefits of a single-dose rather than a two-dose MVA-BN regime.

##### S1.4.7 Evaluation of vaccines strategies

Combining the different rollout scenarios, prioritisation strategies, dose supply and rollout speed (see Table 1 in main text), we ran 378 different scenarios per region. To compare the impact of these strategies, we compared the outcomes to a counterfactual scenario where no vaccinations were given. We use the following metrics throughout the Results sections:

- The **expected number of infections or deaths** was estimated from the no vaccination baseline.
- For each scenario, **averted infections or deaths** at each time step were derived by taking the difference between modelled infections or deaths under the scenario in question and the expected number of infections or deaths.
- Dividing the cumulative infections or deaths averted under a given scenario by the corresponding values under the baseline produced the **percentage of infections or deaths averted** for each scenario.
- We divided the infections or deaths averted by the total doses administered per scenario to obtain **infections or deaths averted per dose**.

The doses administered are the total number of doses given out in the simulated vaccination scenario and will be less than or equal to the total doses available (outlined in Table 1), which are outlined in Table S9. As shown in the supplementary data files, sometimes only a proportion of available vaccinations will be administered if the daily rollout speed is too low to give out all available doses within the two-year time frame.

### S2 Supplementary results

#### S2.1 Model fits

Below, we present the models fitted to the data in each region (Equateur: Figure S2 - Figure S4; Sud Kivu: Figure S5 - Figure S7; Bujumbura: Figure S8 - Figure S9).

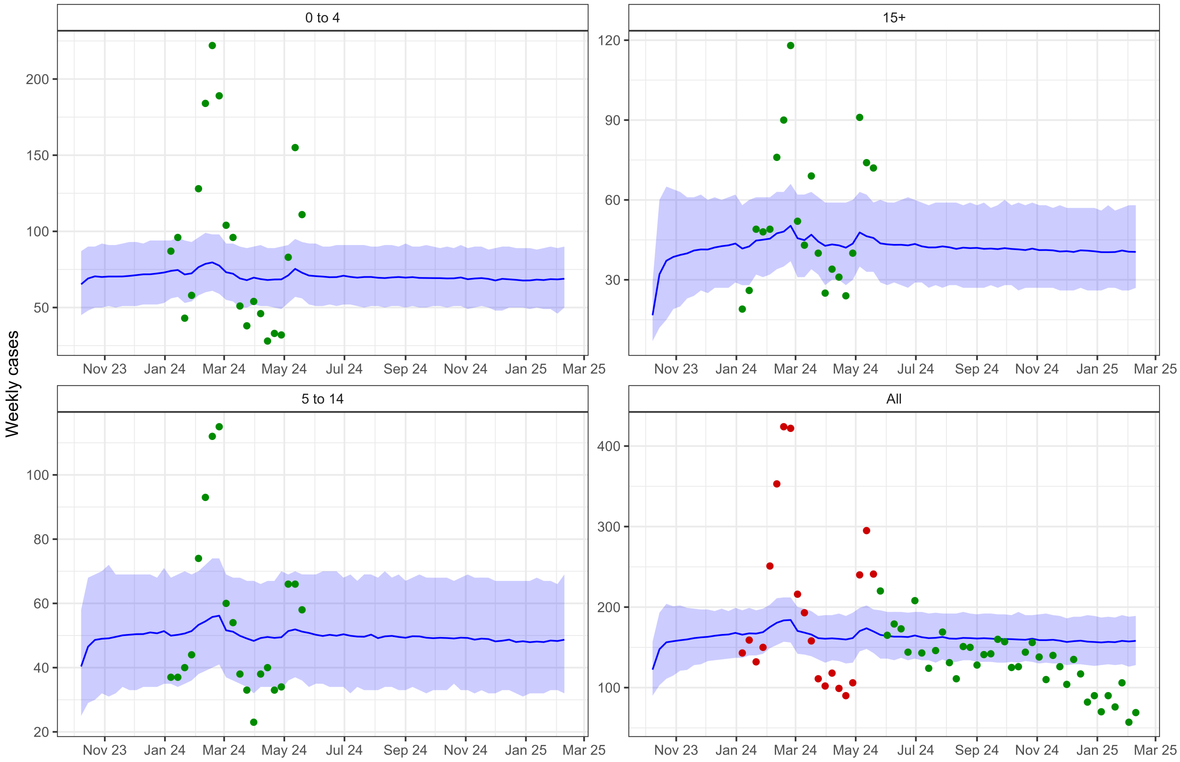

Figure S2: Weekly cases in Equateur stratified by age. Blue line is mean of modelled fit with blue shaded areas representing 95% credible intervals. Green points represent data that we fitted to and red points represent age-aggregated data (that we do not explicitly fit to) at time points where we have cases data by age.

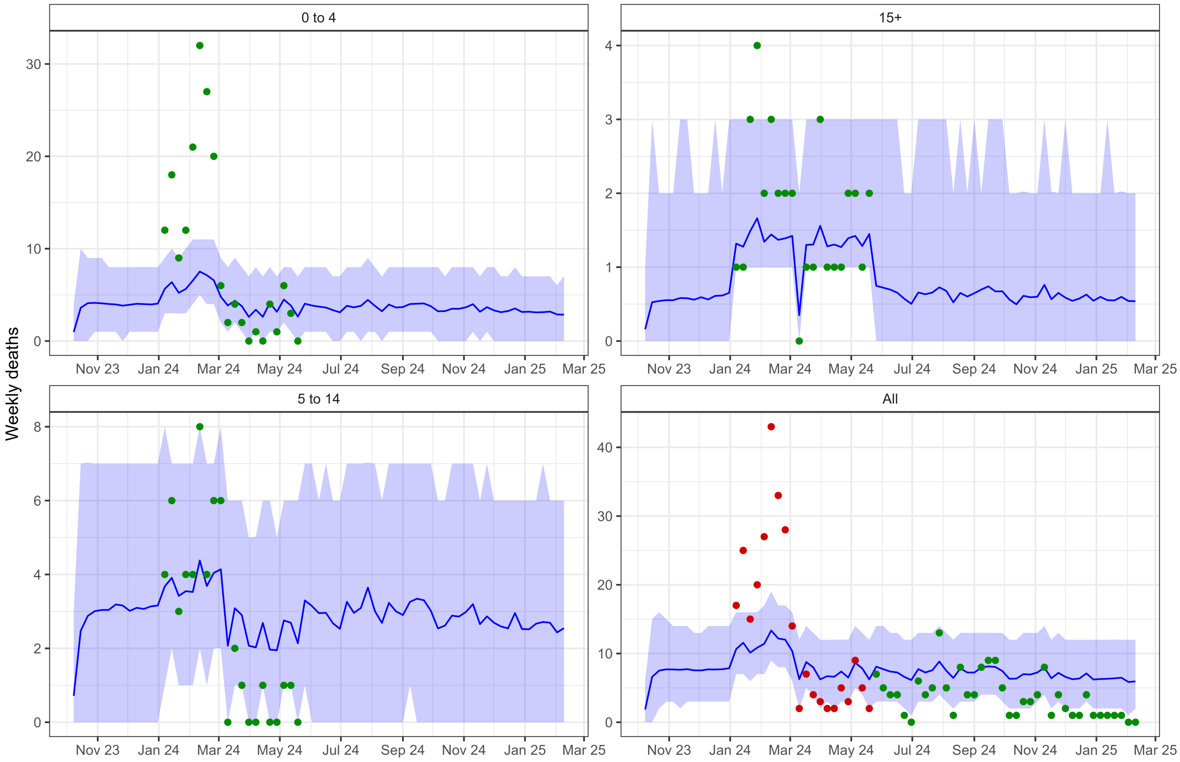

Figure S3: Weekly deaths in Equateur stratified by age. Blue line is mean of modelled fit with blue shaded areas representing 95% credible intervals. Green points represent data that we fitted to and red points represent age-aggregated data (that we do not explicitly fit to) at time points where we have cases data by age.

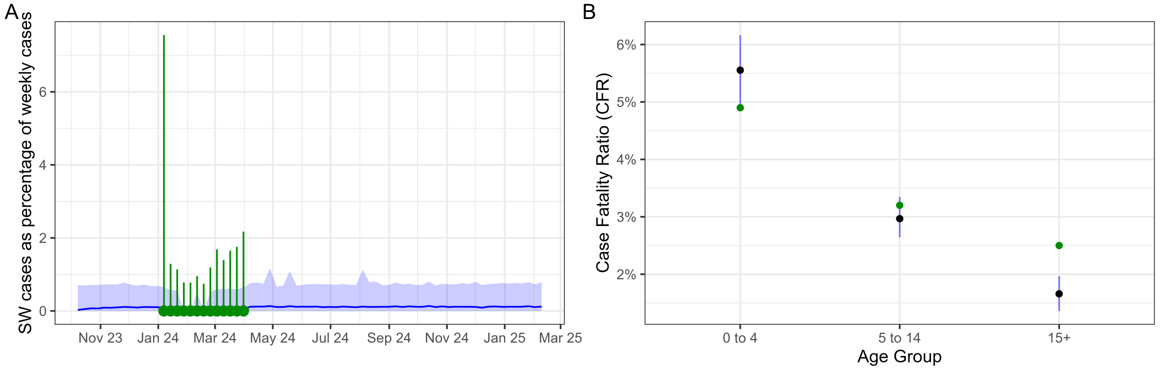

Figure S4: (A) Weekly SW cases in Equateur as percentage of all cases. Blue line is mean of modelled fit with blue shaded areas representing 95% credible intervals. Green points and bars represent mean and 95% binomial confidence intervals of the data that we fitted to. (B) Case fatality ratio in Equateur by age. Blue points are mean of modelled fits with blue bars representing 95% credible intervals. Green points are the data we fitted to.

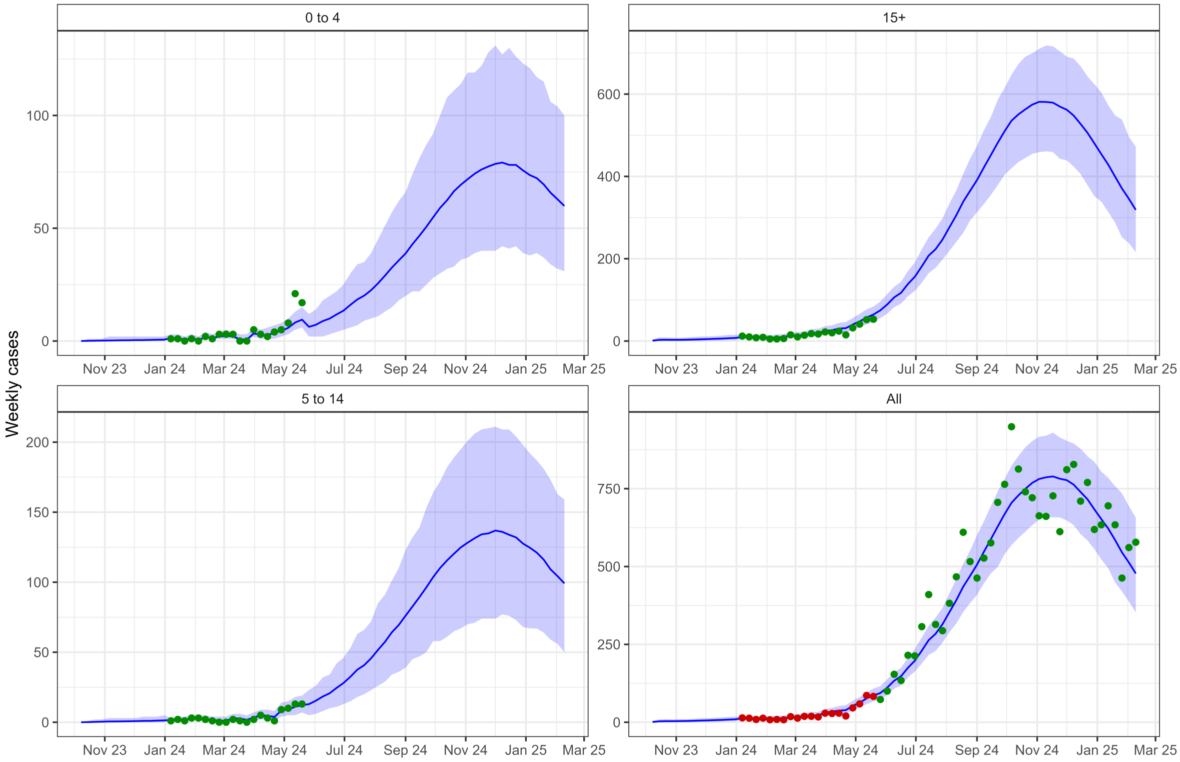

Figure S5: Weekly cases in Sud Kivu stratified by age. Blue line is mean of modelled fit with blue shaded areas representing 95% credible intervals. Green points represent data that we fitted to and red points represent age-aggregated data (that we do not explicitly fit to) at time points where we have cases data by age.

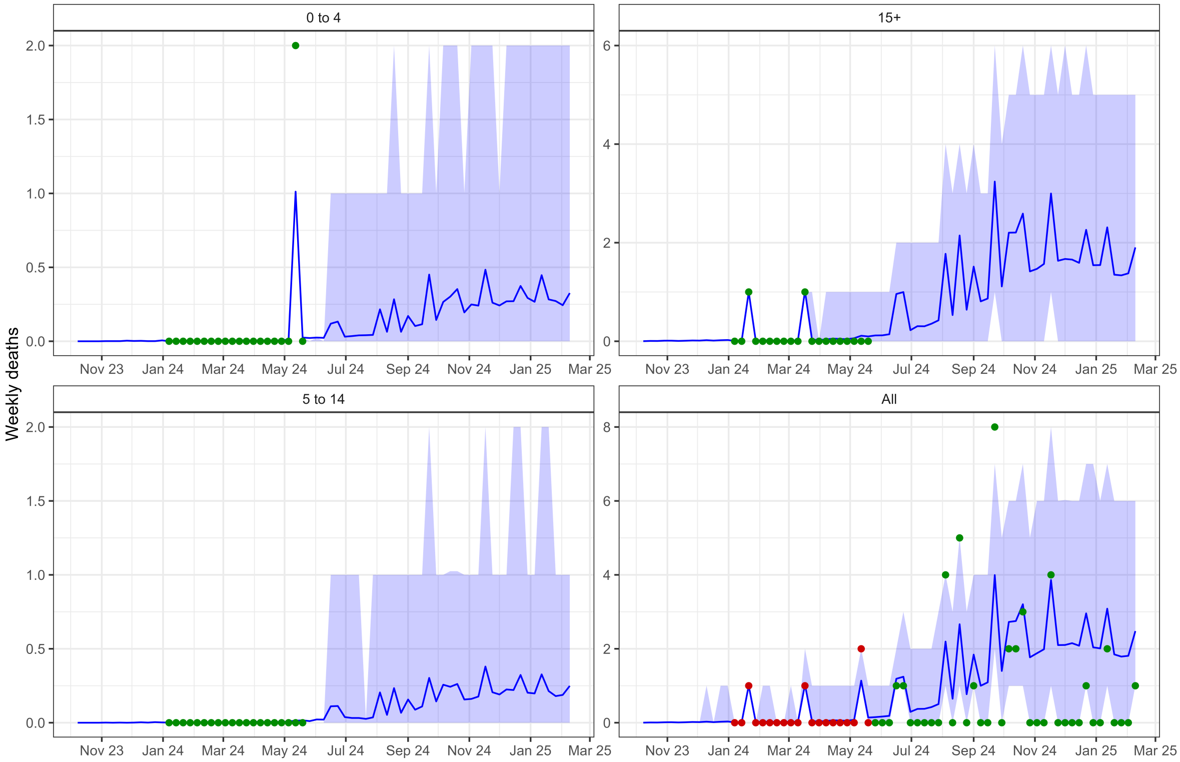

Figure S6: Weekly deaths in Sud Kivu stratified by age. Blue line is mean of modelled fit with blue shaded areas representing 95% credible intervals. Green points represent data that we fitted to and red points represent age-aggregated data (that we do not explicitly fit to) at time points where we have cases data by age.

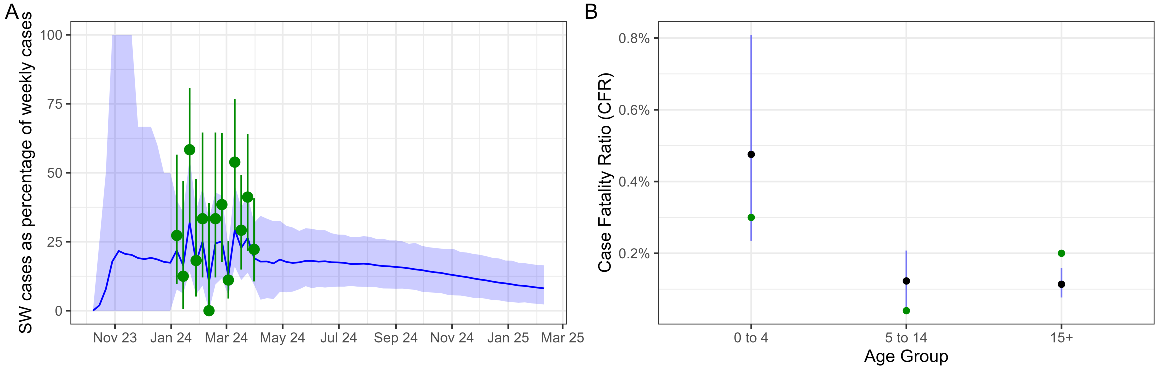

Figure S7: (A) Weekly SW cases in Sud Kivu as percentage of all cases. Blue line is mean of modelled fit with blue shaded areas representing 95% credible intervals. Green points and bars represent mean and 95% binomial confidence intervals of the data that we fitted to. (B) Case fatality ratio in Sud Kivu by age. Blue points are mean of modelled fits with blue bars representing 95% credible intervals. Green points are the data we fitted to.

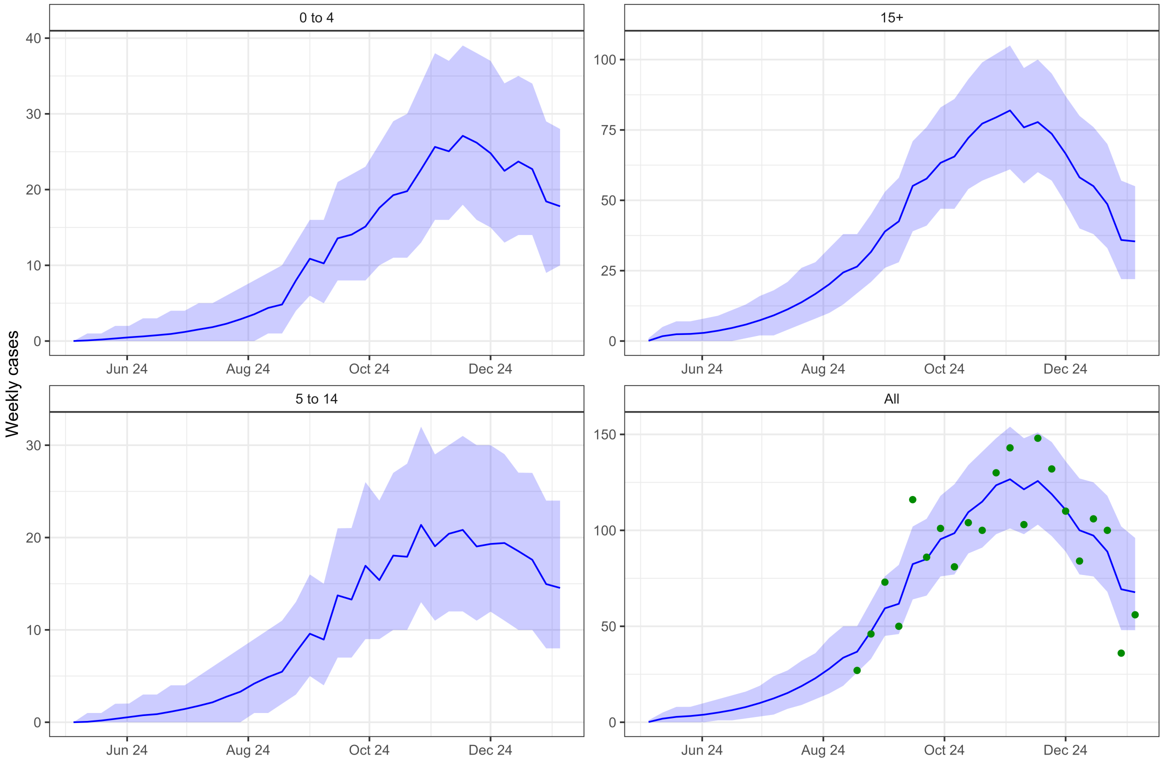

Figure S8: Weekly cases in Bujumbura stratified by age. Blue line is mean of modelled fit with blue shaded areas representing 95% credible intervals. Green points represent data that we fitted to.

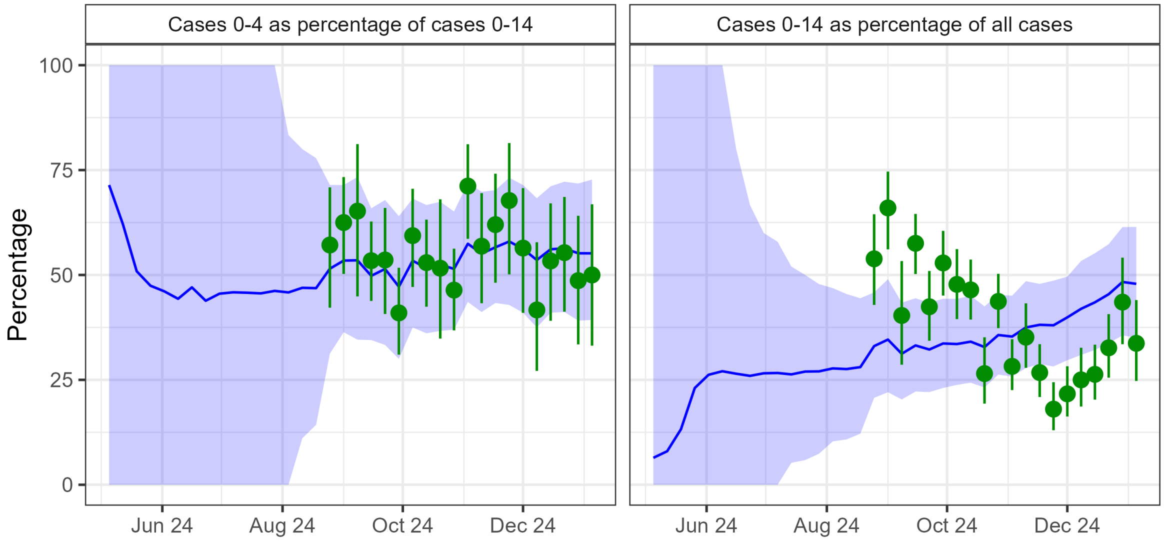

Figure S9: Weekly $0-4$ cases and $0-14$ cases in Bujumbura as percentage of $0-14$ and all cases, respectively. Blue line is mean of modelled fit with blue shaded areas representing 95% credible intervals. Green points and bars represent mean and 95% binomial confidence intervals of the data that we fitted to.

#### S2.2 MCMC diagnostics

Below, we present trace plots, rank plots and pairs plots for fitted parameters in each region (Equateur: Figure S10 - Figure S12; Sud Kivu: Figure S13 - Figure S15; Bujumbura: Figure S16 - Figure S18).

Table S10 presents $\hat{R}$ and the effective sample size ($ESS$) for fitted parameters per region.

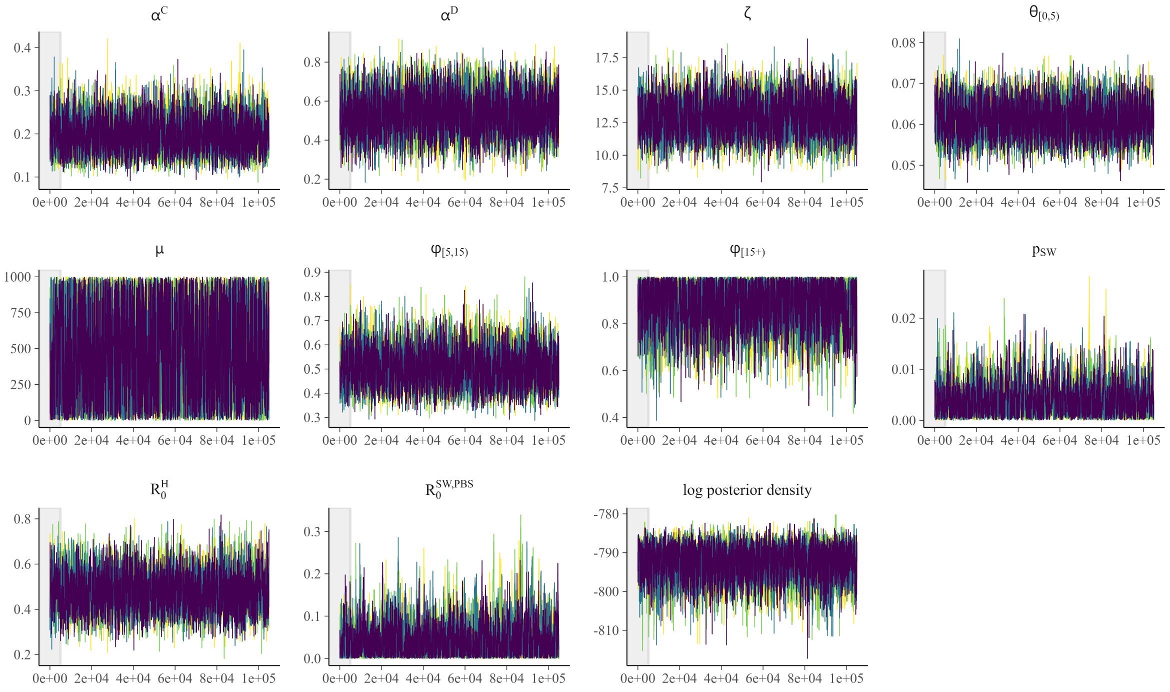

Figure S10: Trace plots of fitted parameters in Equateur. Grey-shaded area indicates burn-in period.

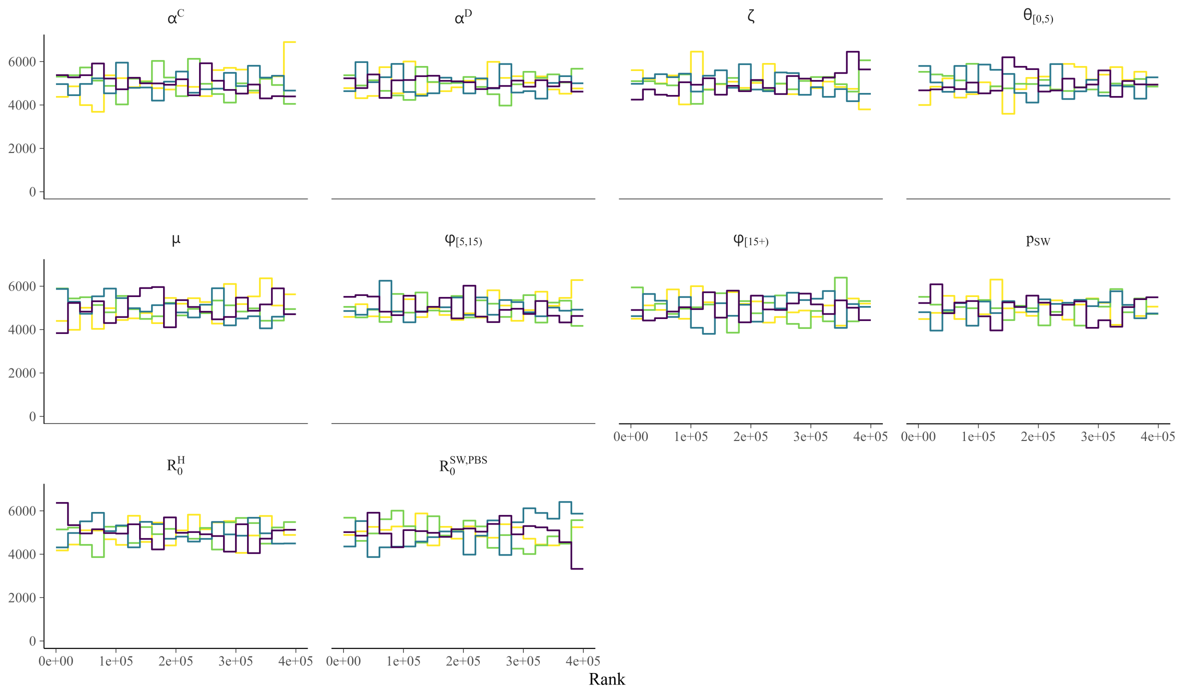

Figure S11: Rank plots of fitted parameters in Equateur.

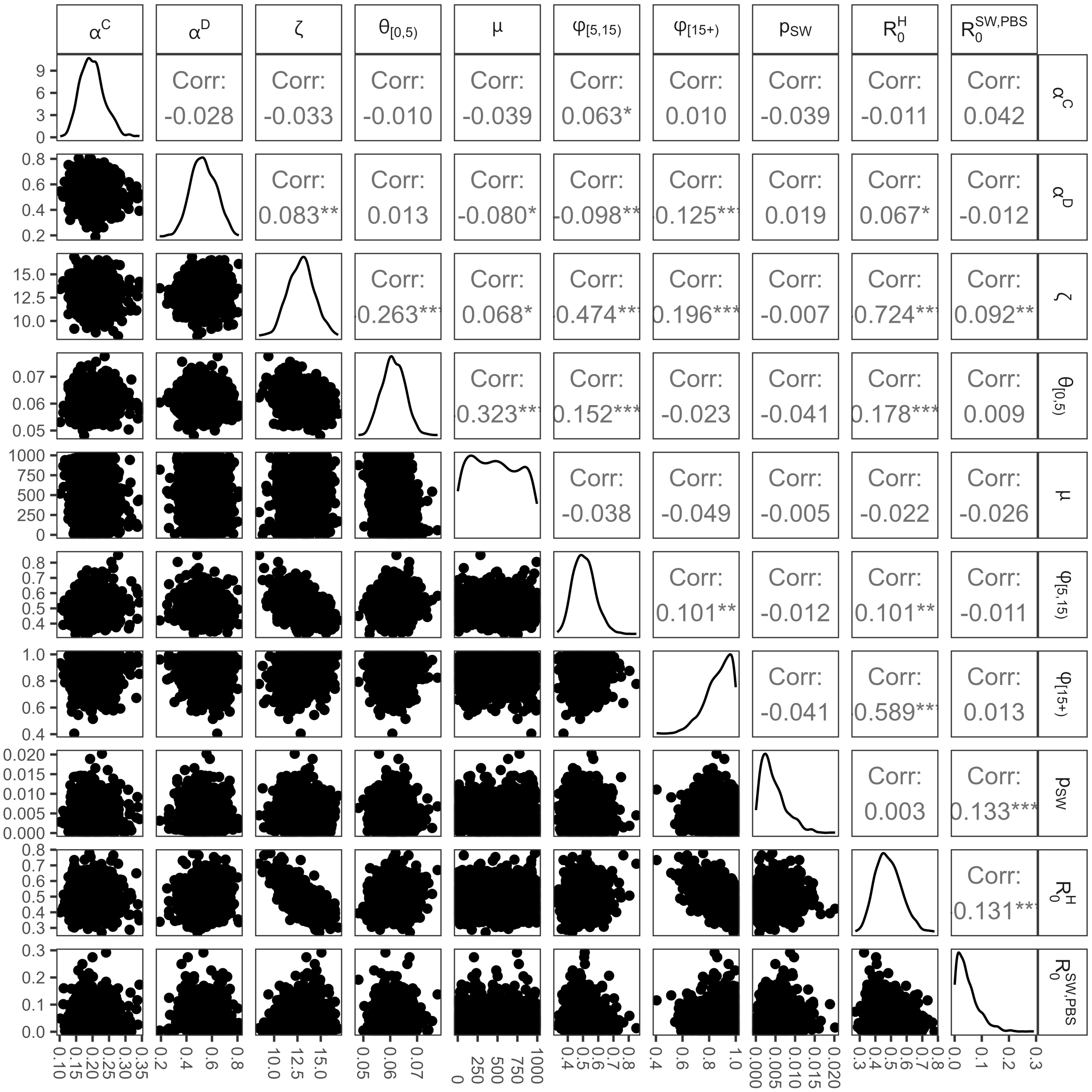

Figure S12: Pairs plot of fitted parameters in Equateur. For Pearson's product moment correlation coefficient on upper diagonal - $***$ indicates $p - value < 0.001$, $**$ indicates $0.001 \leq p - value < 0.01$, $*$ indicates $0.01 \leq p - value < 0.05$, . indicates $0.05 \leq p - value < 0.1$ and otherwise $p - value \geq 0.1$.

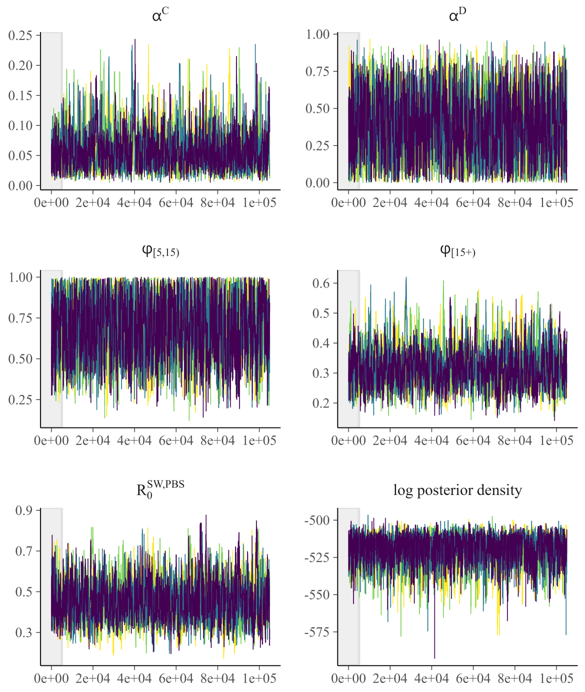

Figure S13: Trace plots of fitted parameters in Sud Kivu. Grey-shaded area indicates burn-in period.

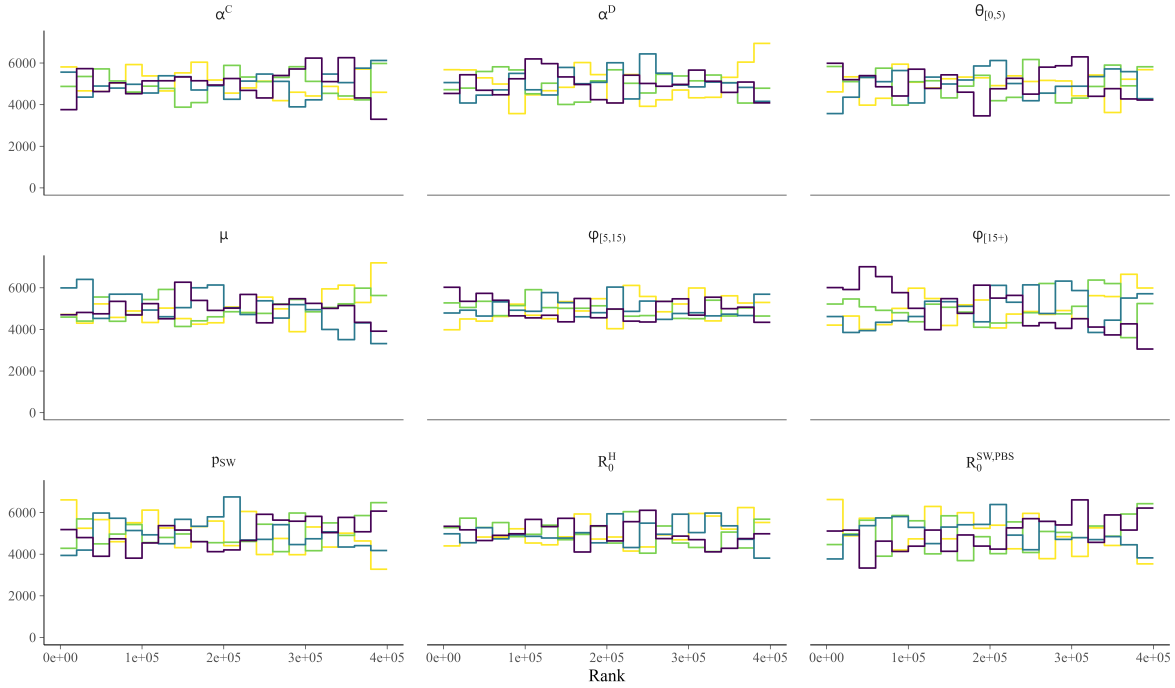

Figure S14: Rank plots of fitted parameters in Sud Kivu.

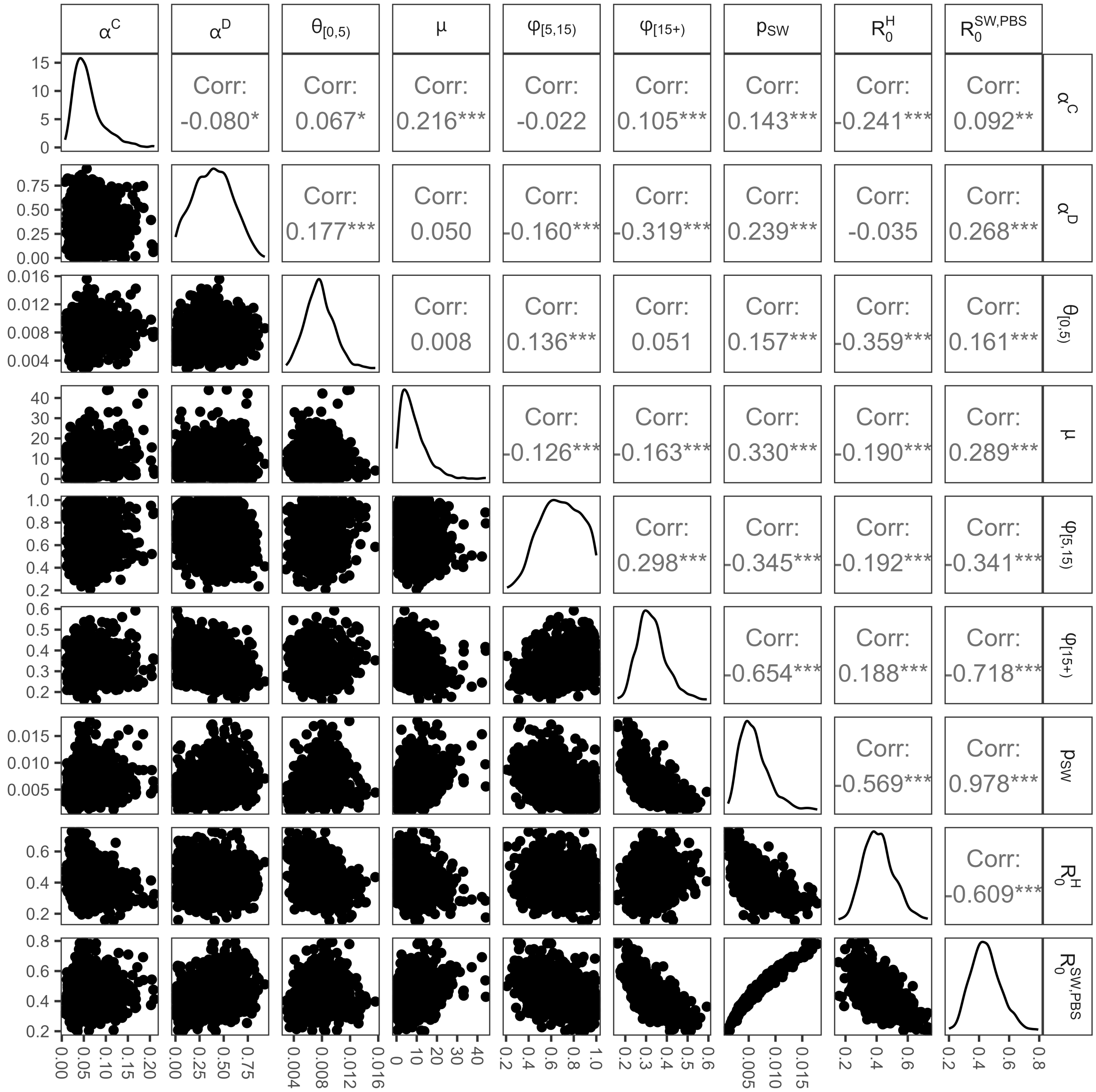

Figure S15: Pairs plot of fitted parameters in Sud Kivu. For Pearson's product moment correlation coefficient on upper diagonal - $***$ indicates $p-value < 0.001$, $**$ indicates $0.001 \leq p-value < 0.01$, $*$ indicates $0.01 \leq p-value < 0.05$, . indicates $0.05 \leq p-value < 0.1$ and otherwise $p-value \geq 0.1$.

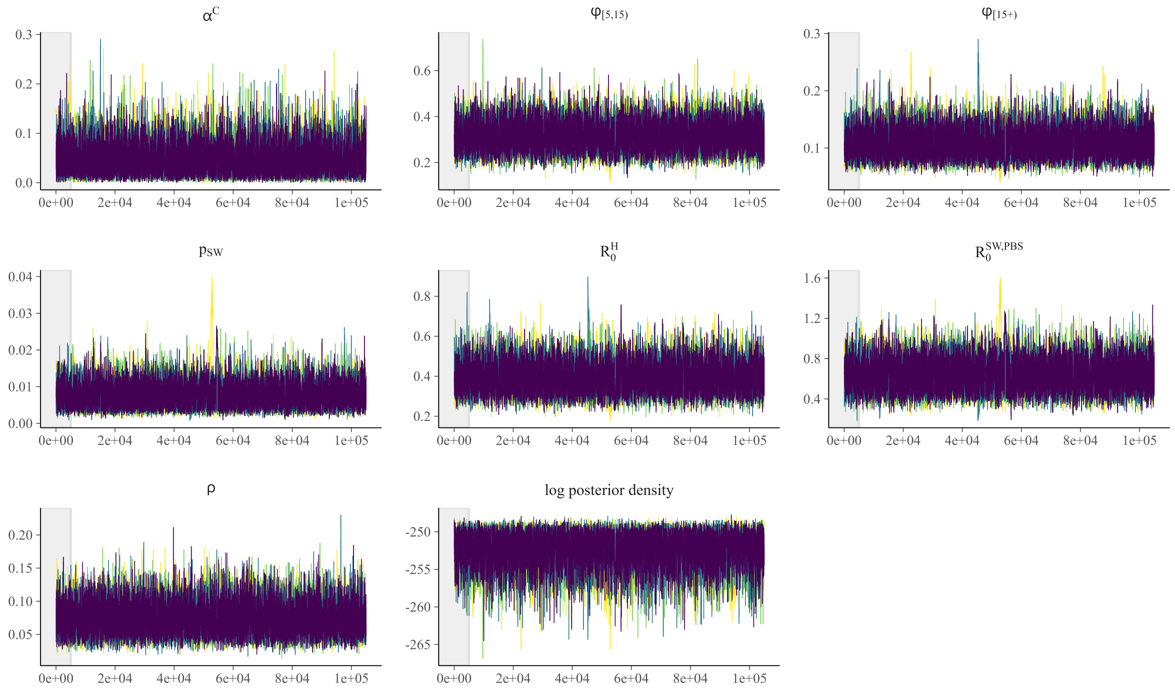

Figure S16: Trace plots of fitted parameters in Bujumbura. Grey-shaded area indicates burn-in period.

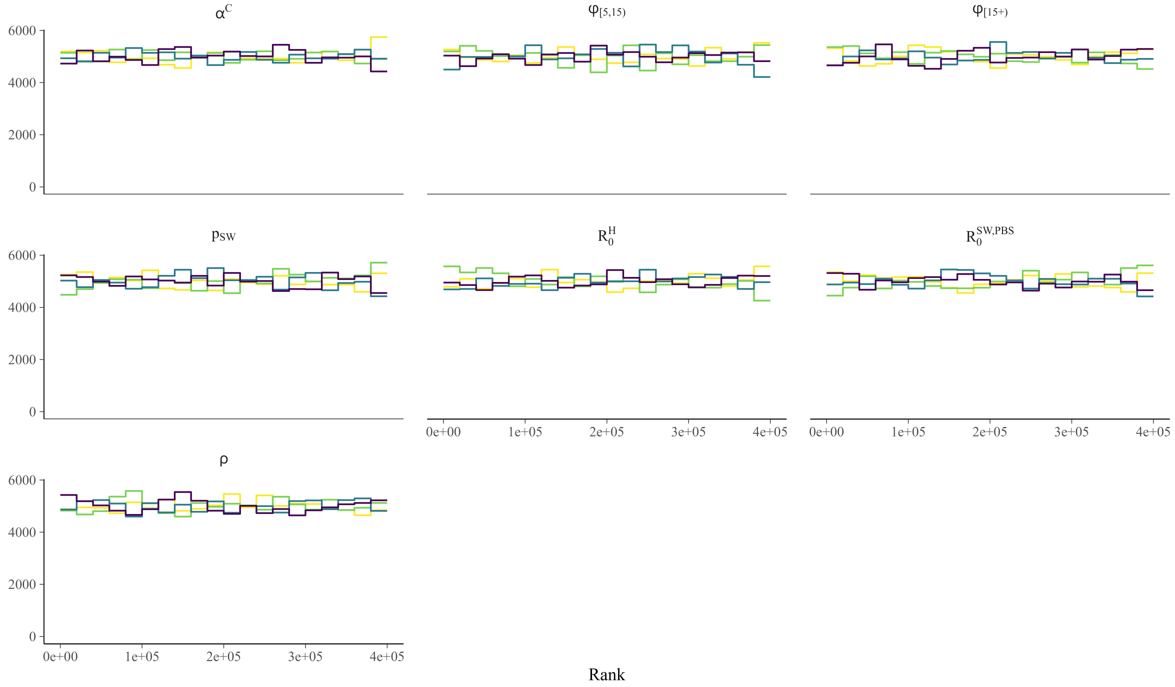

Figure S17: Rank plots of fitted parameters in Bujumbura.

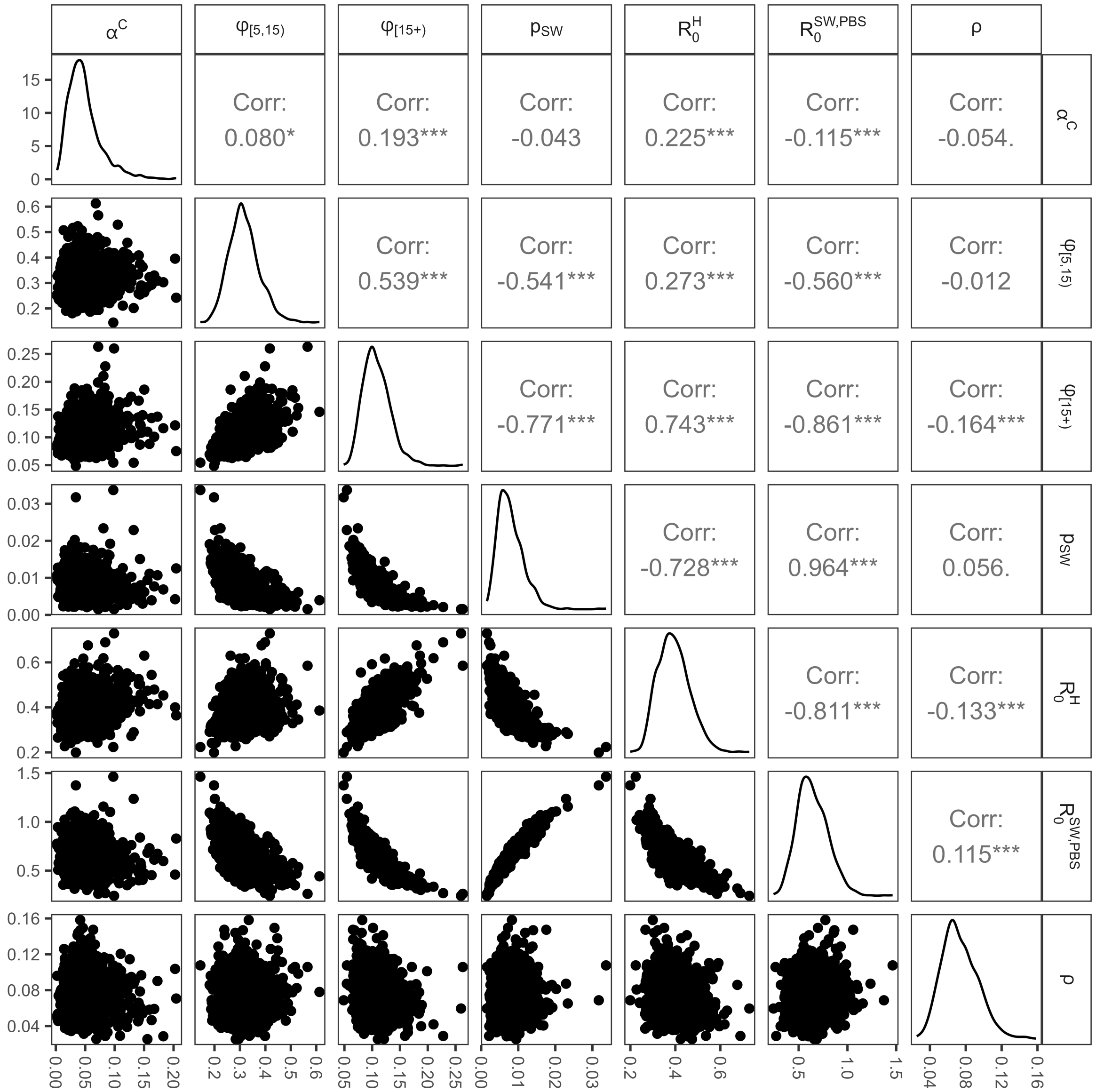

Figure S18: Pairs plot of fitted parameters in Bujumbura. For Pearson's product moment correlation coefficient on upper diagonal - $***$ indicates $p - value < 0.001$, $**$ indicates $0.001 \leq p - value < 0.01$, $*$ indicates $0.01 \leq p - value < 0.05$, . indicates $0.05 \leq p - value < 0.1$ and otherwise $p - value \geq 0.1$.

Table S10: Diagnostic per parameter and region. ($ESS$ = effective sample size).

| **Parameter** | $\hat{R}$ | $ESS_{bulk}$ | $ESS_{tail}$ |
|---|---|---|---|
| **Equateur** | | | |
| $\alpha_C$ | 1.002 | $2,758$ | $3,445$ |
| $\alpha_D$ | 1.001 | $2,831$ | $4,058$ |
| $\zeta$ | 1.002 | $2,926$ | $4,714$ |
| $\theta_{[0,5)}$ | 1.001 | $2,499$ | $4,114$ |
| $\phi_{[5,15)}$ | 1.002 | $2,839$ | $3,753$ |
| $\phi_{\{15+\}}$ | 1.001 | $2,492$ | $3,053$ |
| $p_{SW}$ | 1.002 | $1,824$ | $1,426$ |
| $R_0^{\mathrm{H}}$ | 1.001 | $2,550$ | $3,386$ |
| $R_0^{\mathrm{SW,PBS}}$ | 1.002 | $2,139$ | $1,866$ |
| **Sud Kivu** | | | |
| $\alpha_C$ | 1.001 | $1,197$ | $1,190$ |
| $\alpha_D$ | 1.003 | $1,580$ | $2,772$ |
| $\theta_{[0,5)}$ | 1.002 | $1,841$ | $2,847$ |
| $\mu$ | 1.009 | 584 | 728 |
| $\phi_{[5,15)}$ | 1.004 | $1,357$ | $2,310$ |
| $\phi_{\{15+\}}$ | 1.002 | $1,352$ | $2,100$ |
| $p_{SW}$ | 1.003 | 828 | 949 |
| $R_0^{\mathrm{H}}$ | 1.005 | 906 | $1,220$ |
| $R_0^{\mathrm{SW,PBS}}$ | 1.004 | 819 | $1,069$ |
| **Bujumbura** | | | |
| $\alpha_C$ | 1.000 | $8,083$ | $7,729$ |
| $\phi_{[5,15)}$ | 1.001 | $5,865$ | $5,893$ |
| $\phi_{\{15+\}}$ | 1.001 | $4,502$ | $4,209$ |
| $p_{SW}$ | 1.001 | $3,730$ | $2,304$ |
| $R_0^{\mathrm{H}}$ | 1.001 | $5,108$ | $5,432$ |
| $R_0^{\mathrm{SW,PBS}}$ | 1.001 | $3,902$ | $2,731$ |
| $\rho$ | 1.001 | $10,206$ | $11,130$ |

#### S2.3 Parameter estimation

Table S11 presents the mean and 95% credible intervals for estimated parameters related to case ascertainment, SW population size and transmissibility for the three regions. Table S12 presents the mean and 95% credible intervals for estimated parameters related to overdispersion and seeding. Table S13 presents the estimated daily number of cases in each age band attributable to zoonotic spillover in Equateur. Table S14 presents age- and key-population-stratified estimates of the CFR for Equateur and Sud Kivu. Deaths were not modelled in Bujumbura due to there only being 1 reported death from mpox to date (July 2025). Figure S19 presents $R(t)$ as estimated in each region.

Table S11: Estimates (mean with 95% credible intervals) of fitted parameters related to case ascertainment, sex worker (SW) population size and transmissibility for Equateur, Sud Kivu and Bujumbura. Note that while the proportion of the population that are SW looks consistent across regions, Figure 2 (main text) shows the full range of the distribution beyond this crude summarisation.

| | **Equateur** | **Sud Kivu** | **Bujumbura** |
|---|---|---|---|
| **Case ascertainment** | | | |
| $5-14$ ($\phi_{[5,15)}$) | 51% (39% – 66%) | 69% (35% – 98%) | 32% (21% – 45%) |
| 15+ ($\phi_{\{15+\}}$) | 87% (64% – 99%) | 33% (22% – 48%) | 11% (7% – 16%) |
| Overall | 75% (66% – 83%) | 38% (27% – 53%) | 14%(9%-20%) |
| **SW population (% of women aged $12-49$)** | | | |
| $p_{SW}$ | 0.44% (0.05% – 1.19%) | 0.6% (0.22% – 1.26%) | 0.79% (0.29% – 1.53%) |
| **Transmissibility** | | | |
| Overall ($R_0$) | 0.51 (0.36 – 0.76) | 1.58 (1.47 – 1.69) | 2.01 (1.77 – 2.24) |
| Household ($R_0^{\mathrm{H}}$) | 0.48 (0.34 – 0.66) | 0.41 (0.25 – 0.63) | 0.39 (0.27 – 0.54) |
| Sexual ($R_0^{\mathrm{SW,PBS}}$) | 0.05 (0 – 0.16) | 0.45 (0.28 – 0.66) | 0.64 (0.37 – 0.97) |
| R0 sexual networks ($R_0^{\mathrm{S}}$) | 0.23 (0.01 – 0.73) | 1.57 (1.46 – 1.68) | 2.00 (1.76 – 2.23) |
| **Contribution to transmission** | | | |
| General | 31% (20% – 44%) | 28% 16% – 41% | 24% (16% – 33%) |
| Zoonotic | 69% (56% – 0.80%) | NA | NA |
| Sexual | 0% (0% – 0%) | 72% (58% – 84%) | 76% (67% – 84%) |

Table S12: Estimates (mean with 95% credible intervals) of fitted parameters related to overdispersion across regions and seeding.

| **Parameter** | **Equateur** | **Sud Kivu** | **Bujumbura** |
|---|---|---|---|
| Overdispersion of cases ($\alpha_C$) | 0.2 (0.14 – 0.28) | 0.06 (0.02 – 0.15) | 0.05 (0.01 – 0.12) |
| Overdispersion of deaths ($\alpha_D$) | 0.53 (0.35 – 0.72) | 0.39 (0.03 – 0.77) | NA |
| Poisson mean of number of infections seeded in SWs in Sud Kivu ($\mu$) | NA | 8.3 (0.7 – 23.9) | NA |
| Overdispersion of cases in Bujumbura compared to Burundi as a whole ($\rho$) | NA | NA | 0.07 (0.04 – 0.12) |

Table S13: Estimated zoonotic cases per day in each age group (Equateur).

| **Group** ($i$) | **Zoonotic spillover** ($\beta^{\mathbf{Z}}\psi_i$) |
|---|---|
| 0 – 4 | 9.13 (7.19 – 11.23) |
| 5 – 11 | 9.03 (7.11 – 11.11) |
| 12 – 14 | 1.74 (1.37 – 2.14) |
| 15 – 19 | 0.47 (0.37 – 0.58) |
| 20 – 24 | 0.31 (0.24 – 0.38) |
| 25 – 29 | 0.26 (0.20 – 0.31) |
| 30 – 34 | 0.21 (0.17 – 0.26) |
| 35 – 39 | 0.18 (0.14 – 0.22) |
| 40 – 44 | 0.14 (0.11 – 0.18) |
| 45 – 49 | 0.12 (0.09 – 0.15) |
| 50 – 54 | 0.10 (0.08 – 0.13) |
| 55 – 59 | 0.08 (0.06 – 0.10) |
| 60 – 64 | 0.06 (0.05 – 0.08) |
| 65 – 69 | 0.05 (0.04 – 0.06) |
| 70 – 74 | 0.03 (0.03 – 0.04) |
| 75+ | 0.03 (0.03 – 0.04) |
| SW | 0.00 (0.00 – 0.01) |
| PBS | 0.07 (0.06 – 0.09) |

Table S14: Estimated age- and key-population-stratified case fatality ratio (CFR) in Equateur and Sud Kivu.

| **Group** ($i$) | **Prior** (%) | **Equateur** (%) | **Sud Kivu** (%) |
|---|---|---|---|
| 0–4 | 10.2 | 6.1 (5.3 – 7.0) | 0.8 (0.4 – 1.2) |
| 5–11 | 5.4 | 3.2 (2.8 – 3.7) | 0.4 (0.2 – 0.7) |
| 12–14 | 3.5 | 2.1 (1.8 – 2.4) | 0.3 (0.2 – 0.4) |
| 15–19 | 2.6 | 1.6 (1.4 – 1.8) | 0.2 (0.1 – 0.3) |
| 20–24 | 2.0 | 1.2 (1.0 – 1.4) | 0.2 (0.1 – 0.2) |
| 25–29 | 1.6 | 1.0 (0.8 – 1.1) | 0.1 (0.1 – 0.2) |
| 30–34 | 1.3 | 0.8 (0.7 – 0.9) | 0.1 (0.1 – 0.2) |
| 35–39 | 1.2 | 0.7 (0.6 – 0.8) | 0.1 (0.1 – 0.1) |
| 40–44 | 1.0 | 0.6 (0.5 – 0.7) | 0.1 (0.0 – 0.1) |
| 45–49 | 1.0 | 0.6 (0.5 – 0.7) | 0.1 (0.0 – 0.1) |
| 50–54 | 1.0 | 0.6 (0.5 – 0.7) | 0.1 (0.0 – 0.1) |
| 55 – 59 | 1.0 | 0.6 (0.5 – 0.7) | 0.1 (0.0 – 0.1) |
| 60 – 64 | 1.0 | 0.6 (0.5 – 0.7) | 0.1 (0.0 – 0.1) |
| 65–69 | 1.0 | 0.6 (0.5 – 0.7) | 0.1 (0.0 – 0.1) |
| 70–74 | 1.0 | 0.6 (0.5 – 0.7) | 0.1 (0.0 – 0.1) |
| 75+ | 1.0 | 0.6 (0.5 – 0.7) | 0.1 (0.0 – 0.1) |
| SW | 1.8 | 1.1 (0.9 – 1.2) | 0.1 (0.1 – 0.2) |
| PBS | 1.4 | 0.8 (0.7 – 0.9) | 0.1 (0.1 – 0.2) |

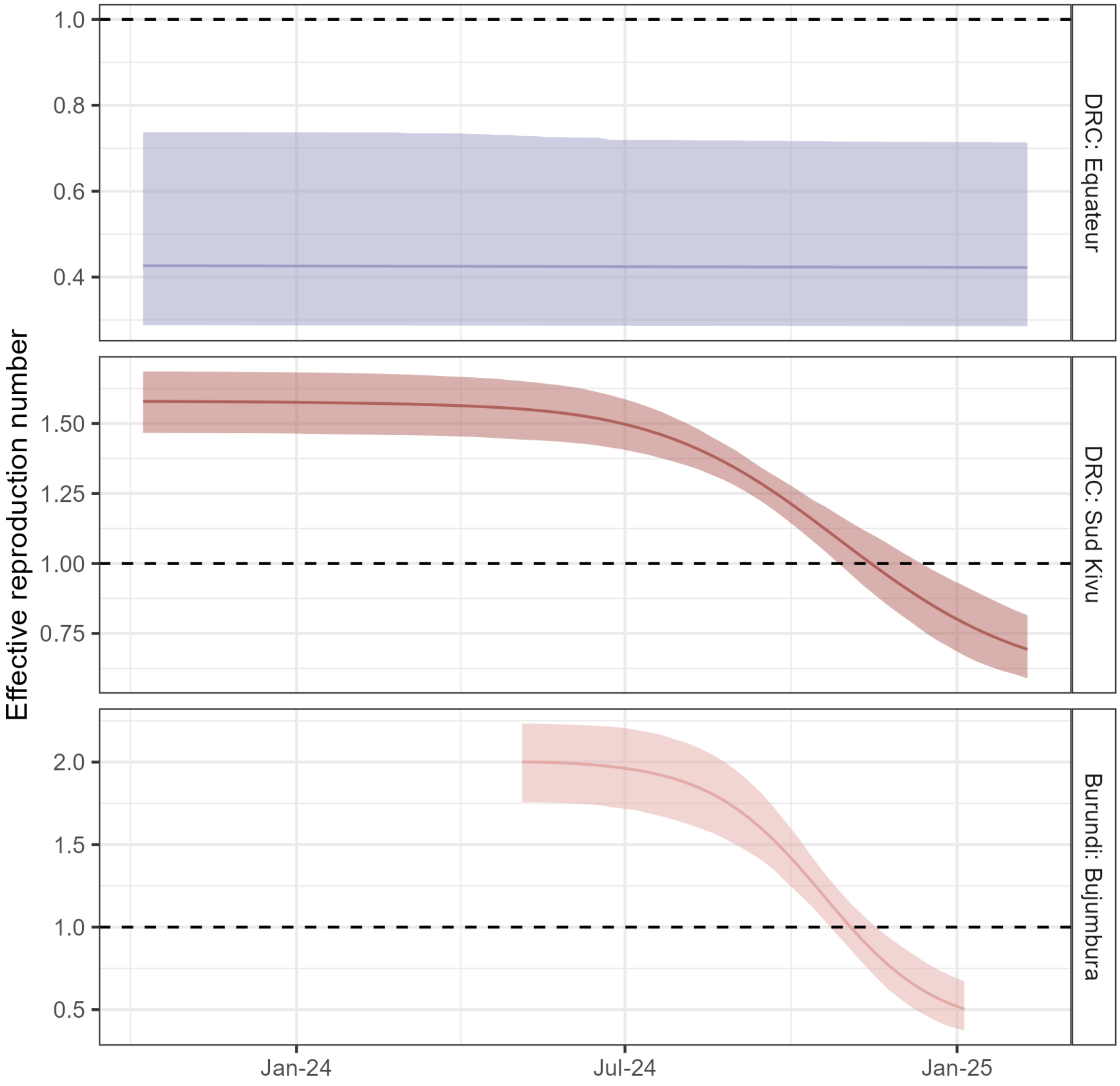

Figure S19: Modelled effective reproduction number ($R(t)$) in each of the three regions (Equateur – top, purple; Sud Kivu – middle, red; Bujumbura – bottom, pink). Dashed black line shows $R(t) = 1$, at which point cases are neither increasing nor decreasing.

#### S2.4 Deaths averted

Figure S20 and Figure S21 present analogous plots to Figure 3 and Figure 4 in the main text for Equateur and Sud Kivu, respectively, but focussing on deaths rather than infections.

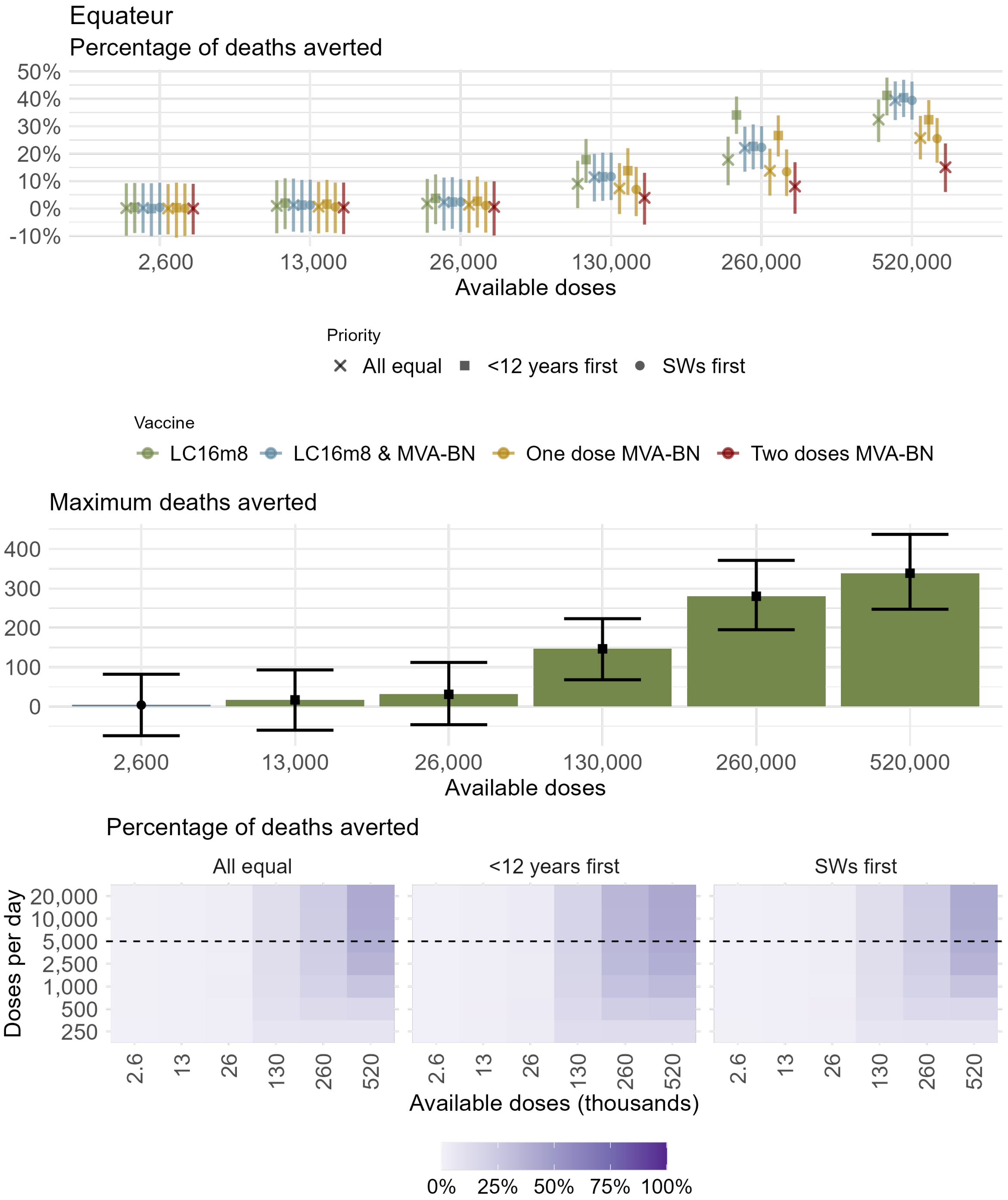

Figure S20: Vaccination scenarios in Equateur. Deaths that could potentially have been averted over the period January 2024 – January 2026 compared with the baseline scenario under the nine modelled vaccination strategies (Table 1 main text) and 6 different dosing levels. (A) Percentage of deaths averted; (B) Maximum number of deaths averted given the best performing vaccine and prioritisation scenario under each dosing level and for the central rollout speed; Colours indicate the four vaccine types while shapes indicate the three prioritisation strategies. Points and bars are posterior means and 95% credible intervals, respectively. (C) Posterior mean percentage of deaths averted for each rollout speed, total available doses and prioritisation strategy. For the corresponding dosing level and rollout speed each grid point represents the maximum number of deaths averted given the best performing vaccine and prioritisation scenario. Horizontal dashed line indicates the central rollout speed presented in A-B.

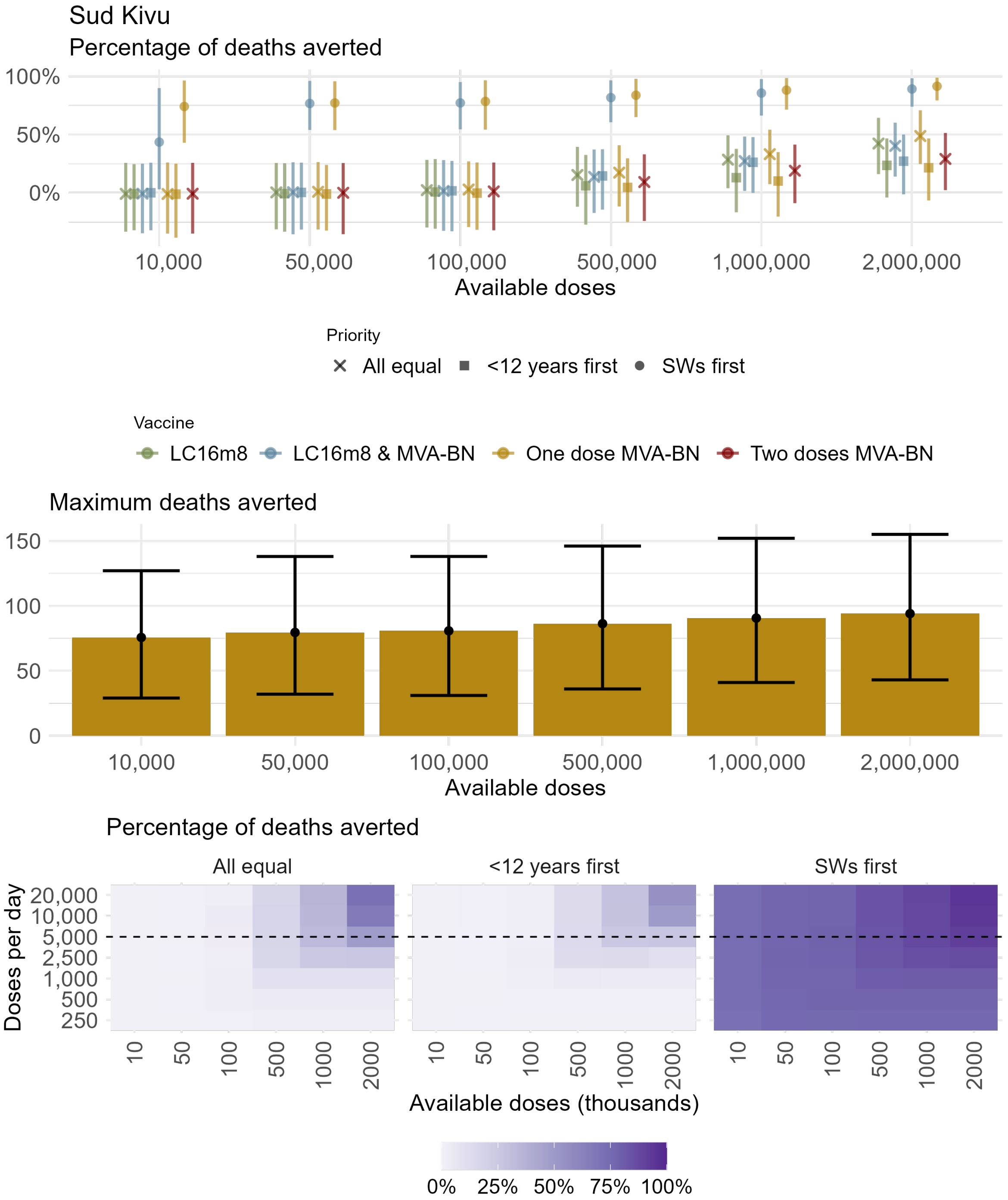

Figure S21: Vaccination scenarios in Sud Kivu. Deaths that could potentially have been averted over the period January 2024 – January 2026 compared with the baseline scenario under the nine modelled vaccination strategies (Table 1 main text) and 7 different dosing levels. (A) Percentage of deaths averted; (B) Maximum number of deaths averted given the best performing vaccine and prioritisation scenario under each dosing level and for the central rollout speed; Colours indicate the four vaccine types while shapes indicate the three prioritisation strategies. Points and bars are posterior means and 95% credible intervals, respectively. (C) Posterior mean percentage of deaths averted for each rollout speed, total available doses and prioritisation strategy. For the corresponding dosing level and rollout speed each grid point represents the maximum number of deaths averted given the best performing vaccine and prioritisation scenario. Horizontal dashed line indicates the central rollout speed presented in A-B.

#### S2.5 Infections and deaths averted per dose

Figure S22 and Figure S23 present the infections and deaths, respectively, averted per vaccine dose in each of the nine vaccine strategies under all dosing levels and for the central rollout speed value.

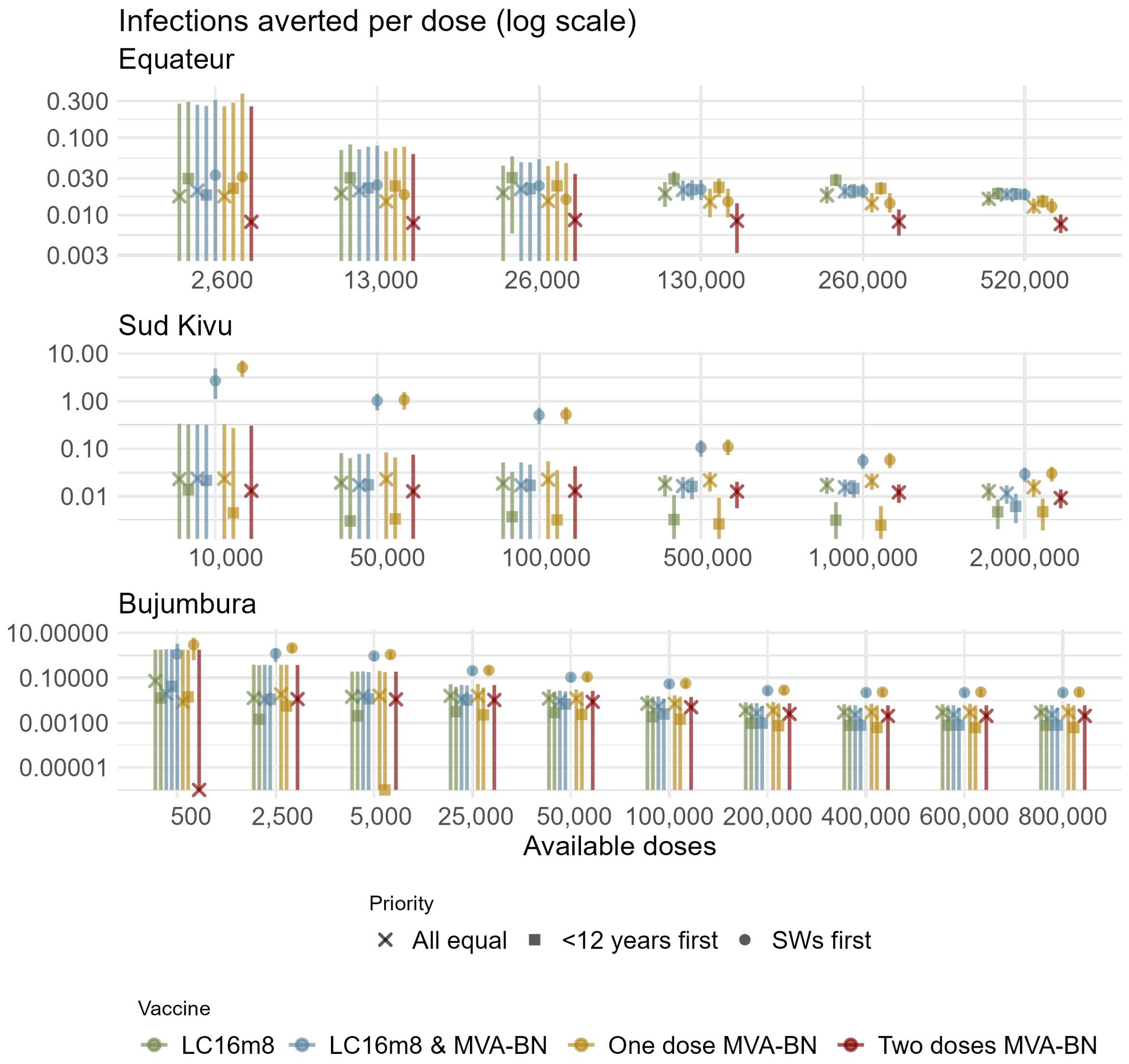

Figure S22: Vaccination scenarios of infections averted per dose (log scale) that could potentially have been averted over the period January 2024 – January 2026 compared with the baseline scenario under the nine modelled vaccination strategies (Table 1 main text) and all dosing levels considered in our analysis. (A) Sud Kivu under a central rollout speed of 5,000; (B) Equateur under a central rollout speed of 5,000; (C) Bujumbura under a central rollout speed of 500. Points and bars are posterior means and 95% credible intervals, respectively. Colours represent the four vaccine type scenarios while shapes indicate the three prioritisation scenarios.

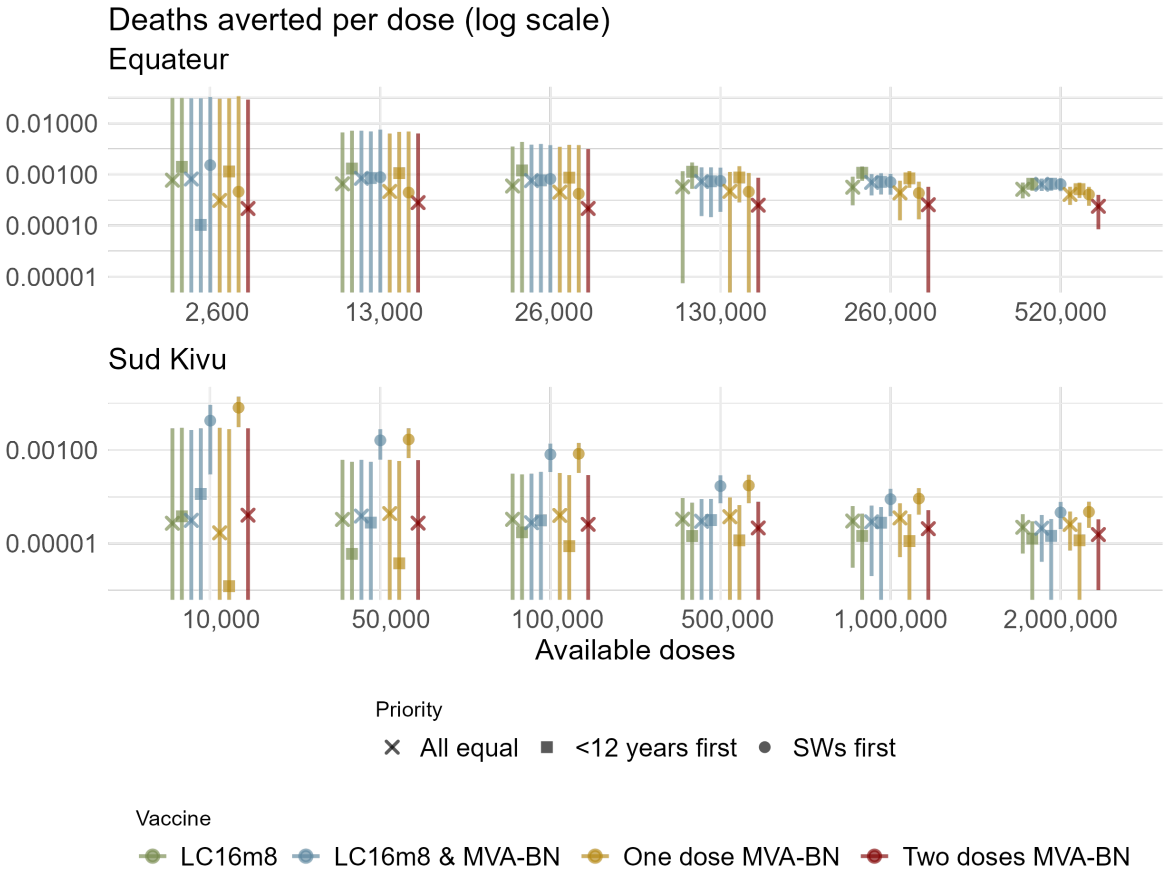

Figure S23: Vaccination scenarios of infections averted per dose (log scale) that could potentially have been averted over the period January 2024 – January 2026 compared with the baseline scenario under the nine modelled vaccination strategies (Table 1 main text) and all dosing levels for a central rollout speed of 5,000. (A) Sud Kivu; (B) Equateur. Points and bars are posterior means and 95% credible intervals, respectively. Colours represent the four vaccine type scenarios while shapes indicate the three prioritisation scenarios.

#### S2.6 Epidemic trajectories under vaccination

Figure S24 and Figure S26 present the trajectories of infections (all) and deaths (Equateur and Sud Kivu only) underpinning the estimates of infections and deaths averted.

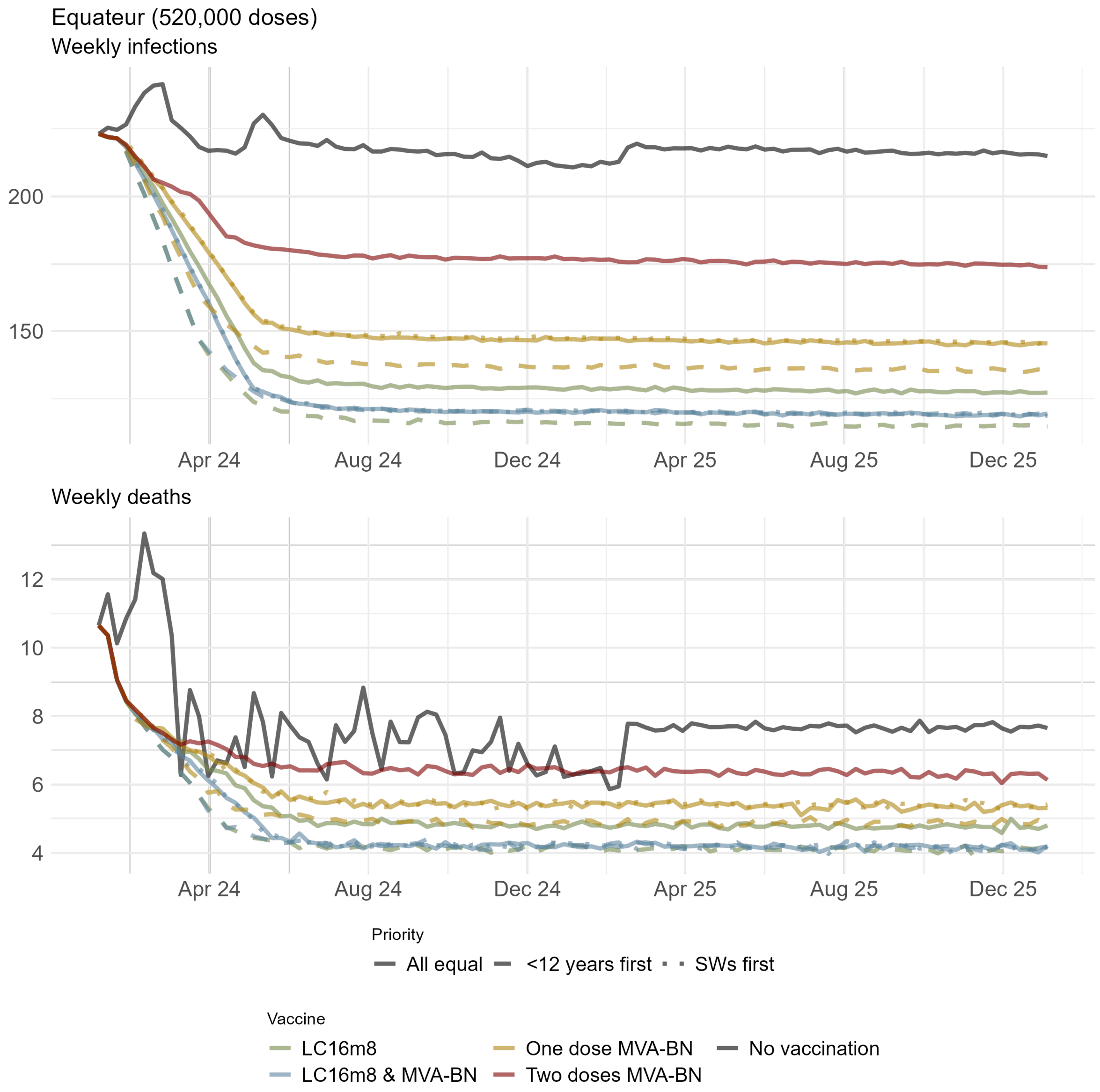

Figure S24: Epidemic trajectories of (A) infections and (B) deaths in Equateur under vaccination. Black solid line indicates a scenario where no vaccination has been implemented and forms the baseline for estimating infections and deaths averted. Colours and line types indicate the different vaccine types and prioritisation scenarios modelled, respectively.

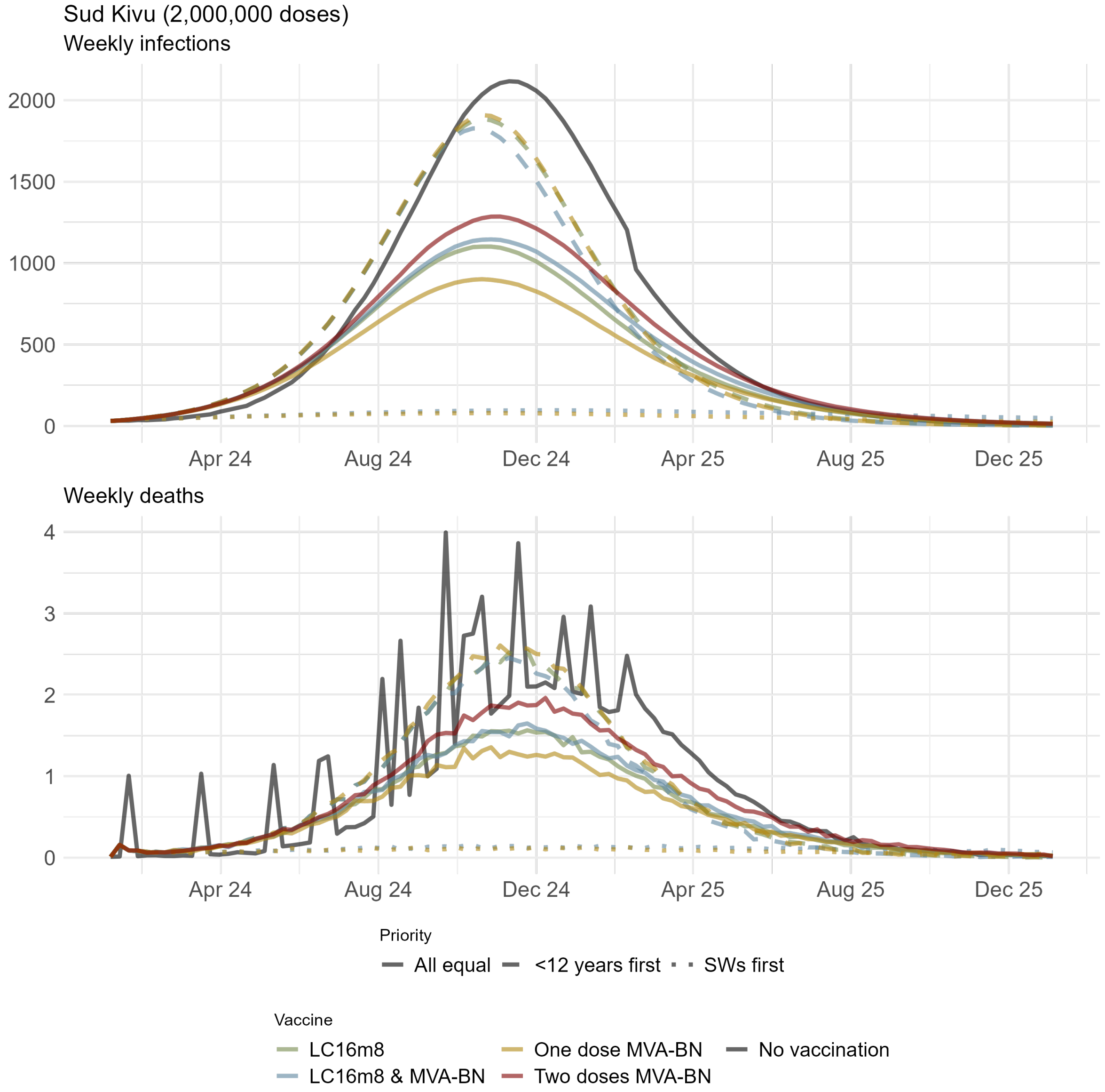

Figure S25: Epidemic trajectories of (A) infections and (B) deaths in Sud Kivu under vaccination. Black solid line indicates a scenario where no vaccination has been implemented and forms the baseline for estimating infections and deaths averted. Colours and line types indicate the different vaccine types and prioritisation scenarios modelled, respectively.

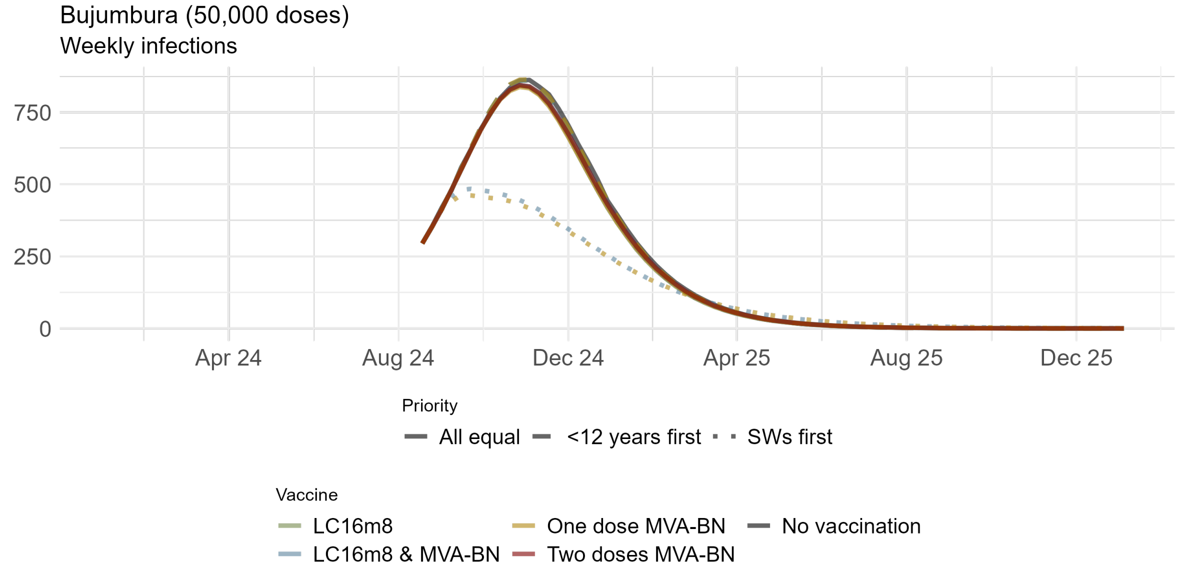

Figure S26: Epidemic trajectories of infections in Bujumbura under vaccination. Black solid line indicates a scenario where no vaccination has been implemented and forms the baseline for estimating infections and deaths averted. Colours and line types indicate the different vaccine types and prioritisation scenarios modelled, respectively.

##### S2.7 Analysis of additional doses in Bujumbura

Figure S27 extends the analysis presented in Figure 5 (main text) to consider the impact of more mpox vaccine doses than COVID-19 vaccine doses being administered in Bujumbura.

Even under 800,000 single doses of MVA-BN given to SWs ahead of the general population, the percentage of infections averted remains at 37% (95% CrI 28% – 48%), as vaccination commences late in the epidemic curve.

In line with the results for Sud Kivu in the main text, giving one dose of MVA-BN consistently averts more infections than administering two doses of MVA-BN, for 800,000 doses 12% (95% CrI 6%–22%) and 8% (95% CrI 2%–15%) of infections are averted respectively.

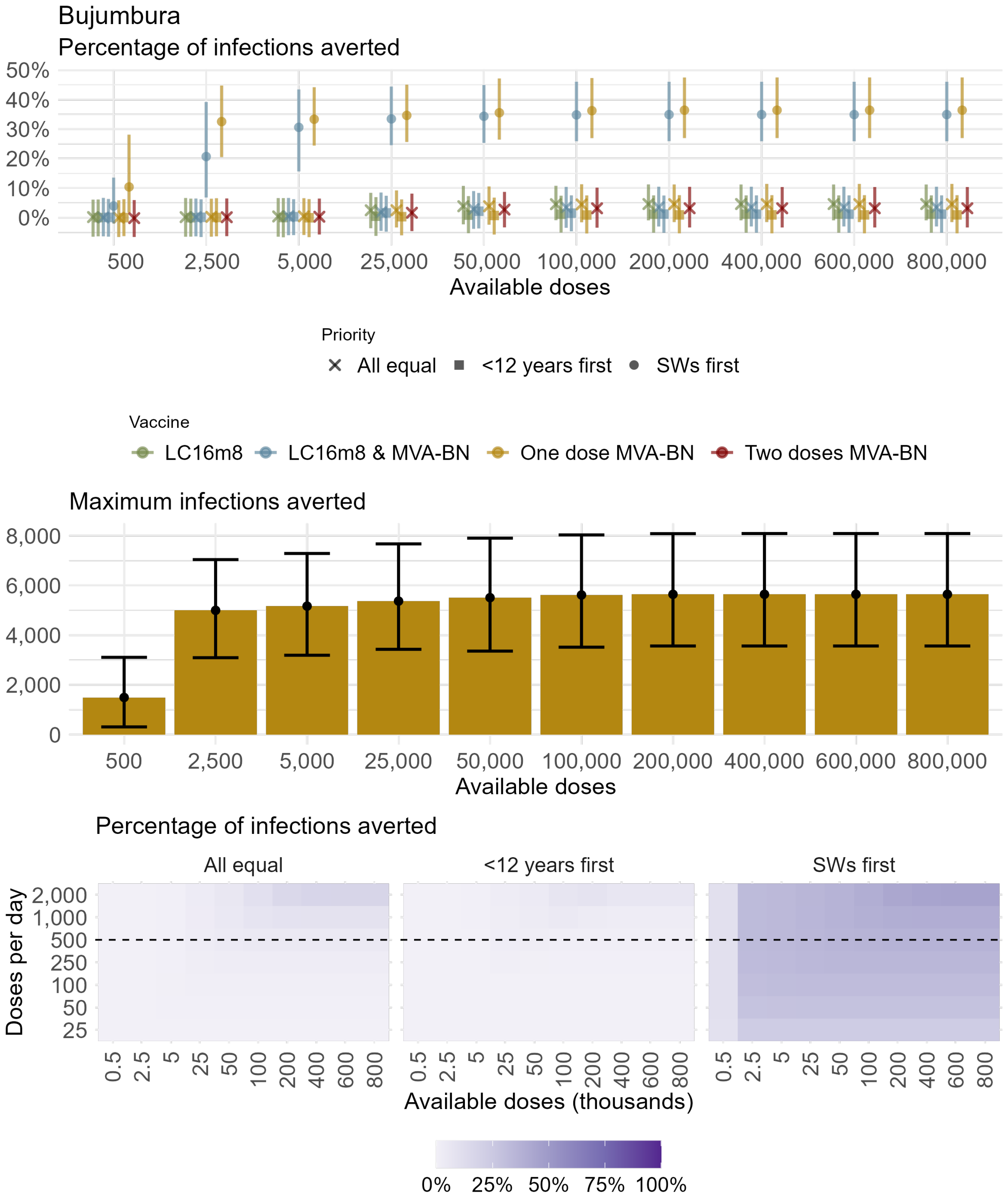

Figure S27: Vaccination scenarios in Bujumbura with dose numbers extended to 800, 000. Infections that could potentially have been averted over the period January 2024 – January 2026 compared with the baseline scenario under the nine modelled vaccination strategies (Table 1 main text) and 7 different dosing levels. (A) Percentage of infections averted; (B) Maximum number of infections averted given the best performing vaccine and prioritisation scenario under each dosing level and for the central rollout speed; Colours indicate the four vaccine types while shapes indicate the three prioritisation strategies. Points and bars are posterior means and 95% credible intervals, respectively. (C) Posterior mean percentage of infections averted for each rollout speed, total available doses and prioritisation strategy. For the corresponding dosing level and rollout speed each grid point represents the maximum number of infections averted given the best performing vaccine and prioritisation scenario. Horizontal dashed line indicates the central rollout speed presented in A-B.
